# The Relationship between Polyunsaturated Fatty Acids and Inflammation: Evidence from cohort and Mendelian randomization analyses

**DOI:** 10.1101/2023.11.27.23299099

**Authors:** Daisy CP Crick, Sarah L Halligan, George Davey Smith, Golam M Khandaker, Hannah J Jones

## Abstract

**Background:** Omega-3 (n-3) and omega-6 (n-6) polyunsaturated fatty acids (PUFAs) are thought to have anti- and pro-inflammatory roles respectively and influence the risk of various chronic diseases. However, it is unclear whether these associations are causal.

**Methods:** We examined the associations of dietary polyunsaturated FAs with biomarkers of systemic inflammation: C-reactive protein (CRP), Glycoprotein Acetyls (GlycA) and Interleukin 6 (IL-6) in two cohort datasets (Avon Longitudinal Study of Parents and Children (ALSPAC; N=2802) and UK Biobank (N=12,401)) using multivariable analyses. We investigated causality using two-sample Mendelian Randomization (MR). In addition to the inverse-variance weighted (IVW) method, we used sensitivity analyses to strengthen causal inference. We conducted multivariable MR (MVMR) to investigate causal effects of n-3 and n-6 on inflammation, accounting for the low-density lipoprotein (LDL)-cholesterol, triglycerides, mono-unsaturated FAs, and saturated FAs.

**Results:** Cohort analyses show a positive association between the n-6:n-3 ratio and each biomarker. Total n-3 and n-6 PUFAs were associated with higher GlycA levels ([mean-difference=0.33; 95% CI=0.29, 0.36, and 0.52; 95% CI=0.48, 0.55] respectively). The MR results suggest that total n-3 FAs cause higher circulating CRP (IVW=0.09; 95% CI=0.03, 0.16) and GlycA levels (0.12; 95% CI=0.04, 0.21). The positive association between n-3 FAs and GlycA remained in the MVMR analysis, after accounting for LDL-cholesterol, triglycerides, mono-unsaturated FAs and saturated FAs.

**Conclusions:** We find no convincing evidence of a simple pro- and anti-inflammatory dichotomy regarding the function of n-6 and n-3 PUFAs. Further research is needed to better understand mechanisms underlying the effects of PUFAs on specific immune biomarkers.

**Key messages:** Omega-3 (n-3) and omega-6 (n-6) polyunsaturated fatty acids (PUFAs) are thought to have anti- and pro-inflammatory roles respectively, but it is unclear whether these associations are causal.

Contrary to current understanding, n-3 FAs are not associated with lower inflammatory marker levels (C-Reactive protein (CRP) or Glycoprotein Acetyls (GlycA)). We report both n-3 and n-6 PUFAs are associated with higher GlycA and CRP levels.

Our findings argue against the presence of a simple pro- and anti-inflammatory dichotomy regarding the function of n-6 and n-3 PUFAs, respectively, and suggests that n-3 supplementation alone may not reduce systemic inflammation.

---

Polyunsaturated fatty acids (PUFAs), are proposed to influence systemic inflammation^(1)^. There are two families of PUFAs which are essential for many metabolic processes: Omega-3 (n-3)^(2, 3)^; and Omega-6 (n-6)^(3)^. Some PUFAs like docosahexaenoic acid (DHA; an n-3 PUFA (22:6-n3)) can come from both exogenous sources (such as meat, egg products and seafood) and endogenous sources (synthesised from their metabolomic precursors through desaturation and elongation reactions^(4)^). However, linolic acid (LA: an n-6 PUFA (18:2-n6)) can only be acquired through dietary consumption^(5)^.

Evidence suggests that n-3 PUFAs have anti-inflammatory effects and reduce the severity/occurrence of inflammation-related conditions^(6, 7)^, given that cytokines and acute-phase proteins are implicated in the pathophysiology of many non-communicable diseases (NCDs)^(8–16)^. A meta-analysis of 14 clinical trials (n=135,291) suggested that n-3 supplementation reduced the risk of adverse cardiovascular events/death^(17)^. However, an RCT investigating dietary intake and mortality in 3114 men with angina, found that risk of cardiac death was higher among individuals advised to eat oily fish/take fish oil, compared to a control group^(18)^. Additionally, studies have reported that the consumption of n-6 PUFAs had no effect on concentrations of inflammatory biomarkers, despite reports that they were pro-inflammatory^(19–22)^. However, these studies do not directly assess FAs impact on inflammation, and the contradictory results raise questions of whether PUFAs *causally* affect inflammation or whether changes to inflammatory biomarker levels are the result of residual confounding.

Both n-3 and n-6 PUFAs are metabolised by the same enzymes and compete for desaturation and elongation. Therefore, n-6 PUFAs act as competitive inhibitors of n-3 PUFAs and thus reduce the amount of end-product n-3 PUFAs that can be synthesised^(23)^. Therefore, the plasma n-6:n-3 ratio may also impact the concentration of inflammatory makers and subsequent health^(24–27)^. Research to understand the importance of the n-6:n-3 ratio on levels of inflammation and its impact on the occurrence of NCDs is of public health concern.

We investigated the effect of specific n-3 and n-6 PUFAs, which play a role in key metabolic processes on inflammatory markers using (1)Traditional observational epidemiological analyses and (2) Mendelian randomization (MR)^(28)^.

Our exposures were DHA and LA, positioned at opposite ends of the biosynthesis pathways (Supplementary Figure S1). To further interrogate the PUFA-inflammation relationship, we included total n-3 PUFAs, total n-6 PUFAs and the ratio of total n-6:total n-3 PUFAs. Outcomes included C-Reactive Protein (CRP) and Interleukin-6 (IL-6), commonly used biomarkers of systemic inflammation and Glycoprotein Acetyls (GlycA). GlycA is derived from multiple proteins involved in the acute-phase response and consequently, is thought to be a more stable marker of inflammation compared to single proteins like CRP ^(29–31)^. Our aim was to assess how PUFAs effect inflammation using specific and broad measures.

## 2. Methods

### 2.1. Observational Epidemiological Analyses

The primary analysis used the Avon Longitudinal Study of Parents and Children (ALSPAC) birth cohort which recruited a total of 14,541 pregnant women residing in South-West England^(32–34)^. For eligibility criteria see the study flowchart (Supplementary Figure S2). Participants were included if they had exposure (DHA, LA, total n-3 and total n-6) and outcome (CRP, IL-6 and GlycA levels) data available at 24y.

We used maternal self-reported highest educational qualification and highest occupation of either parent (measures of social economic position (SEP)), maternal and paternal smoking pattern during pregnancy (measures of pregnancy health), participants’ sex, smoking status, alcohol-drinking status and triglyceride, low density lipoprotein-cholesterol (LDL-cholesterol), saturated fatty acid (SFA) and mono-unsaturated fatty acids (MUFA) levels at 24y as covariates.

For additional information on ALSPAC, blood sampling and data collection/processing see Supplementary materials.

Linear regression analyses were used to examine the cross-sectional associations of PUFA levels with GlycA, CRP and IL-6 levels at age 24y. We ran an unadjusted model, a model adjusted for sex, age, maternal and paternal smoking patterns during pregnancy and SEP (model 2), a model including all covariates from model 2 and triglycerides and LDL-cholesterol (model 3) and finally a model which included all covariates from model 3 and SFAs and MUFAs (model 4). Due to high correlations between PUFAs, lipids, MUFAs and SFAs, results from model 2 were used as our main findings. Results from model 3 and 4 should be interpreted with multicollinearity in mind (variance inflation factors presented in Supplementary Figure S3). Details about data cleaning are presented in the Supplementary materials. Analyses were run using the whole sample and sex stratified. We used multiple imputation (MI) (described in detail in the Supplementary materials) to impute missing exposure, outcome, and covariate data in the eligible sample (N=2802).

We replicated the analysis using data from UK Biobank (N=12,401), a community-based, prospective study (https://www.ukbiobank.ac.uk)^(35)^. Despite, the larger sample size, UKB served as replication because it was used in the MR analysis. To strengthen confidence in results (triangulation) we used different methods in different cohorts^(36)^. Information about UKB and the replication analysis is presented in the eMethods.

### 2.2. Two-sample Mendelian Randomisation Analysis

We conducted two-sample MR using genome-wide significant single nucleotide polymorphisms (SNPs; P< 5·0 x10^-8^) as instrumental variables (IVs) from European GWAS of PUFAs^(37)^, GlycA^(38)^, CRP^(39)^ and IL-6^(40)^. SNPs were harmonized, aligning the genetic association for exposure and outcome on the effect allele using the effect allele frequency. See Supplementary materials for details on instrument selection and number of remaining instruments.

The inverse variance weighted (IVW)^(41)^ method was the primary analysis to calculate effect estimates. MR-Egger, weighted median, and weighted mode methods were used as sensitivity analyses. As these methods make different assumptions about instrument validity consistency across primary and sensitivity analyses increased confidence that findings were robust. Methods to investigate pleiotropy and heterogeneity are described in the Supplementary materials.

We replicated the MR analysis using different outcome datasets and results were consistent with the primary analysis. The GWAS used in this secondary analysis and results are discussed in the Supplementary materials.

### 2.3. MR analysis focusing on specific PUFA genes

The *FADS* gene cluster and *ELOVL2* gene encode key desaturase and elongase enzymes respectively and are involved in the n-3 and n-6 FA biosynthesis pathways. Therefore, we conducted a complementary and mechanistically informative MR analysis using only SNPs from within or close to (+/-500 kb) the *FADS* gene cluster (*FADS1*, *FADS2* and *FADS3*; chromosome 11:61,560,452–61,659,523) and *ELOVL2* gene (chromosome 6:10,980,992– 11,044,624) from the DHA GWAS and the LA GWAS. SNPs were selected and harmonised using the same method as in the primary analysis. Details are presented in the Supplementary materials.

### 2.4. Multivariable MR analysis

We conducted an MVMR analysis^(42)^ (MVMR-1) to estimate the independent direct causal effects of n-3 and n-6 PUFAs on the biomarkers of inflammation. Although SNPs within the FADs region should be reliable instruments of PUFAs due to their proximal relation to PUFA biosynthesis, results may be biased because the SNPs can also associate with non-fatty acid traits, such as LDL-cholesterol and triglycerides^(43)^. Therefore, we ran two further MVMR analyses: MVMR-2; as in MVMR-1 but also accounting for triglycerides and LDL-cholesterol and, MVMR-3; as in MVMR-2 but also accounting MUFAs and SFAs. Diagrams representing the intertwined metabolic pathway of LDL-cholesterol, triglycerides and FAs and their effects on inflammation are presented in Supplementary Figure S4.

MVMR analyses are further discussed in Supplementary materials.

We used MR-Lap to assess potential bias in effect estimates due to sample overlap, winner’s curse, or weak instruments^(44)^. Given that SNPs can influence the outcome in distinct ways via discrete biological mechanisms^(45)^ we explored heterogeneity using MR-Clust^(45)^. Greater detail of MR-Lap and MR-Clust is provided in Supplementary materials.

## 3. Results

Table 1 presents the median/range for PUFA levels and inflammatory biomarker levels in ALSPAC and UKB. Results are discussed in detail in the Supplementary materials. A summary figure of all results is presented in Figure 1.

**Figure 1:**
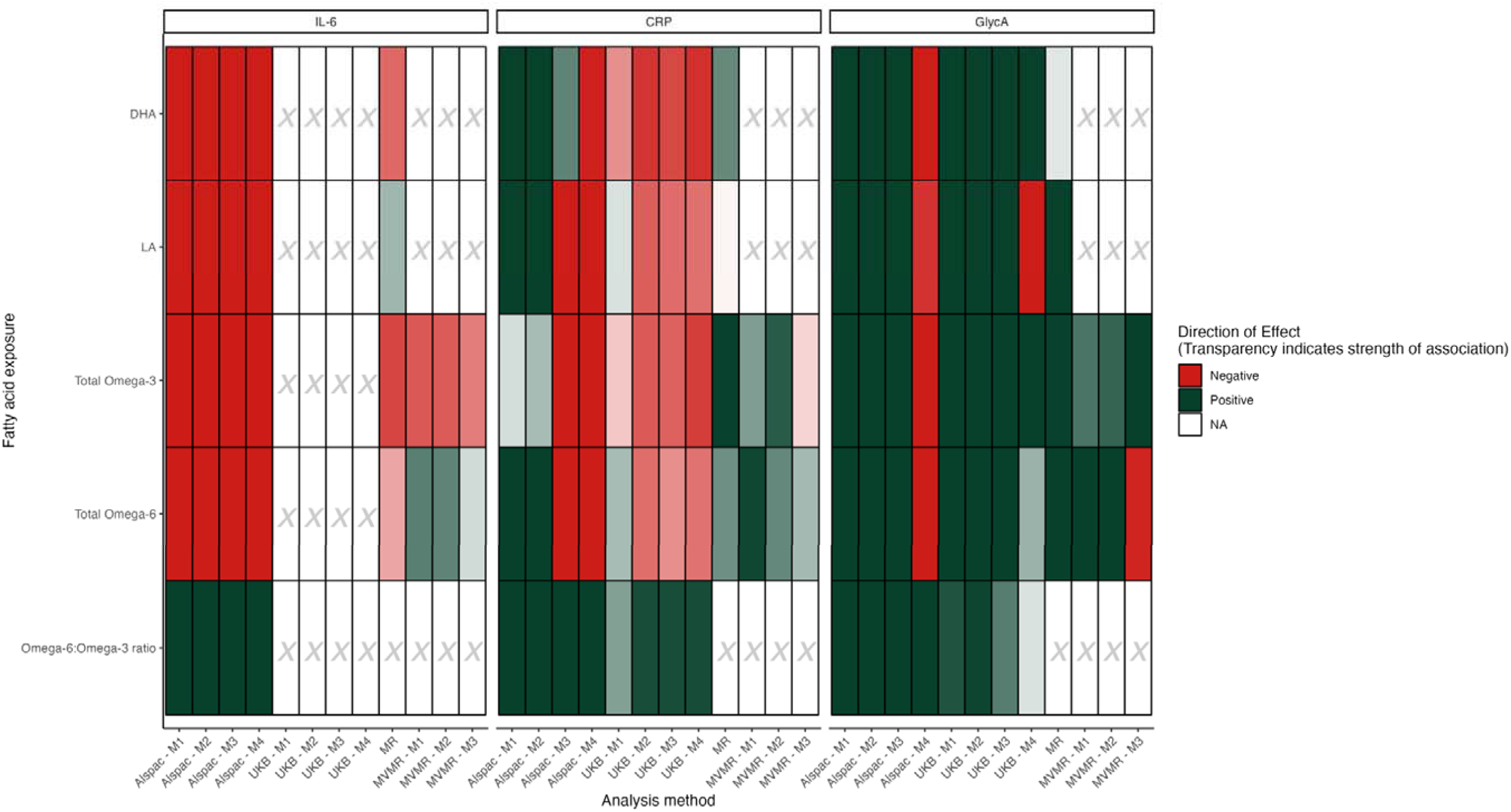
Summary figure of results from the cohort, Mendelian randomization (MR) and Multivariable MR (MVMR) analyses. Green represents when a positive association occurs, whereas red represents a negative association. Transparency indicates the strength of association, where bold colours are used for smaller p-values and muted colours are used for larger p-values. DHA, docosahexaenoic acid; LA, linoleic acid; IL-6, interleukin-6; CRP, C-reactive protein; GlycA, Glycoprotein acetyls; ALSPAC, Avon Longitudinal Study of Parents and Children; UKB, UK Biobank; MR, Mendelian randomization; MVMR, Multivariable Mendelian randomization For cohort analyses (ALSPAC and UKB): M1:no adjustment, M2: adjustment for sex, age, maternal and paternal smoking patterns during pregnancy and SEP, M3: adjustment for covariates from M2, triglycerides and Low-Density Lipoprotein-cholesterol, M4: covariates included in M3, saturated fatty acids (SFAs) and mono-unsaturated fatty acids (MUFAs). For MVMR analyses M1: inclusion of both total omega-3 and total omega-6 as exposures, M2: inclusion of exposures from M1, triglycerides and low-density lipoprotein cholesterol, M3: inclusion of exposures from M2, saturated fatty acids and mono-unsaturated fatty acids. ALT text: Heatmap summarizing the associations between various fatty acid exposures and three inflammation-related outcomes (interleukin-6 (IL-6), C-reactive protein (CRP), and Glycoprotein Acetyls (GlycA)) across a range of analysis methods. Rows represent fatty acid (FA) exposures: docosahexaenoic acid (DHA), linoleic acid (LA), Omega-3, Omega-6 and the ratio of Omega-6 to Omega-3. Columns are grouped by outcome (IL-6, CRP, and GlycA), with each group subdivided by analysis method used. Analyses include traditional regression models using both Avon Longitudinal Study of Parents and Children and UK Biobank data, MR and MVMR. Cells within the matrix are color-coded based on the direction and strength of association. The figure emphasizes the comparative consistency of findings across methods and outcomes. For example, DHA is consistently negatively associated with IL-6, while the majority of FAs are positively associated with GlycA. A transparency gradient gives a visual cue for the strength of association where darker cells represent more robust results.

**Table 1:** Median and interquartile range of exposure and outcome data.

|  | ALSPAC |  | UK Biobank* |  |
| --- | --- | --- | --- | --- |
|  | Median (IQR) | Range | Median (IQR) | Range |
| GlycA (mmol/L) | 1.22 (1.12-1.34) | 0.84-2.25 | 0.007 (-0.66-0.69) | -3.91- 4.00 |
| CRP (mg/l) | 0.87 (0.39-2.29) | 0.1-70.05 | 1.33 (0.66-2.73) | 0.08-77.32 |
| IL-6 (NPX log2) | 3.32 (2.96-3.85) | 1.82-9.86 | - | - |
| DHA | 0.11 (0.09-0.13) | 0.04-0.32 | 0.004 (-0.68-0.66) | -3.75-3.89 |
| LA | 2.31 (1.98-2.68) | 0.89-4.91 | 0.0002 (-0.67-0.67) | -3.59-4.20 |
| Total n-3 PUFAs | 0.29 (0.24-0.34) | 0.07-0.80 | 0.01 (-0.67-0.67) | -3.73-3.60 |
| Total n-6 PUFAs | 2.85 (2.48-3.28) | 1.23-5.82 | -0.001 (-0.66-0.67) | -3.66-4.06 |

*GlycA, Glycoprotein Acetyls; CRP, C-reactive protein; IL-6, Interleukin-6; DHA, Docosahexaenoic acid; LA, Linoleic acid; Total n-3 PUFAs, Total omega-3 poly-unsaturated fatty acids; Total n-6 PUFAs, Total omega-6 poly-unsaturated fatty acids.*
*\* UKB data are presented as inverse normal transformed with exception of CRP due to data availability.*

### 3.1. Results for primary analyses (model 2) using ALSPAC cohort data

Results for all models are presented in Figure and Table 2.

**Table 2:** Associations of PUFA and inflammatory biomarkers using imputed ALSPAC data (N = 2802)

|  | **Model 1 (Unadjusted)** |  | **Model 2\*** |  | **Model 3\*\*** |  | **Model 4\*\*\*** |  |
| --- | --- | --- | --- | --- | --- | --- | --- | --- |
|  | **Mean difference per SD increase in exposure (se)** | **95% CI** | **Mean difference per SD increase in exposure (se)** | **95% CI** | **Mean difference per SD increase in exposure (se)** | **95% CI** | **Mean difference per SD increase in exposure (se)** | **95% CI** |
| **Outcome** |  |  |  |  |  |  |  |  |
| **Exposure: DHA at age 24y** |  |  |  |  |  |  |  |  |
| IL-6 at 24y | -0.13 (0.02) | -0.16, -0.09 | -0.13 (0.02) | -0.17, -0.09 | -0.19 (0.02) | -0.23, -0.15 | -0.20 (0.03) | -0.25, -0.15 |
| CRP at 24y | 0.12 (0.02) | 0.08, 0.16 | 0.09 (0.02) | 0.05, 0.13 | 0.02 (0.02) | -0.02, 0.06 | -0.06 (0.03) | -0.11, -0.01 |
| GlycA at 24y | 0.28 (0.02) | 0.25, 0.32 | 0.31 (0.02) | 0.27, 0.35 | 0.12 (0.02) | 0.09, 0.15 | -0.09 (0.02) | -0.12, -0.05 |
| **Exposure: LA at age 24y** |  |  |  |  |  |  |  |  |
| IL-6 at 24y | -0.08 (0.02) | -0.12, -0.04 | -0.07 (0.02) | -0.11, -0.04 | -0.25 (0.02) | -0.30, -0.21 | -0.38 (0.04) | -0.45, -0.31 |
| CRP at 24y | 0.09 (0.02) | 0.05, 0.12 | 0.08 (0.02) | 0.04, 0.11 | -0.10 (0.02) | -0.14, -0.05 | -0.45 (0.04) | -0.52, -0.38 |
| GlycA at 24y | 0.47 (0.02) | 0.44, 0.51 | 0.48 (0.02) | 0.45, 0.51 | 0.26 (0.02) | 0.22, 0.29 | -0.04 (0.03) | -0.10, 0.01 |
| **Exposure: Total n-3 at age 24y** |  |  |  |  |  |  |  |  |
| IL-6 at 24y | -0.15 (0.02) | -0.19, -0.11 | -0.14 (0.02) | -0.18, -0.10 | -0.27 (0.02) | -0.31, -0.23 | -0.33 (0.03) | -0.38, -0.27 |
| CRP at 24y | 0.004 (0.02) | -0.03, 0.04 | 0.01 (0.02) | -0.03, 0.04 | -0.13 (0.02) | -0.17, -0.09 | -0.32 (0.03) | -0.37, -0.26 |
| GlycA at 24y | 0.31 (0.02) | 0.28, 0.35 | 0.32 (0.02) | 0.29, 0.36 | 0.07 (0.02) | 0.03, 0.10 | -0.19 (0.02) | -0.23, -0.16 |
| **Exposure: Total n-6 at age 24y** |  |  |  |  |  |  |  |  |
| IL-6 at 24y | -0.06 (0.02) | -0.09, -0.02 | -0.06 (0.02) | -0.09, -0.02 | -0.25 (0.03) | -0.30, -0.20 | -0.58 (0.05) | -0.68, -0.48 |
| CRP at 24y | 0.15 (0.02) | 0.11, 0.18 | 0.13 (0.02) | 0.09, 0.16 | -0.02 (0.03) | -0.07, 0.03 | -0.57 (0.05) | -0.67, -0.47 |
| GlycA at 24y | 0.51 (0.02) | 0.47, 0.54 | 0.51 (0.02) | 0.48, 0.55 | 0.31 (0.02) | 0.27, 0.35 | -0.13 (0.04) | -0.20, -0.06 |
| **Exposure: Total-6:total n-3 at age 24y** |  |  |  |  |  |  |  |  |
| IL-6 at 24y | 0.20 (0.02) | 0.16, 0.23 | 0.18 (0.02) | 0.15, 0.22 | 0.18 (0.02) | 0.15, 0.22 | 0.17 (0.02) | 0.13, 0.21 |
| CRP at 24y | 0.21 (0.02) | 0.17, 0.24 | 0.17 (0.02) | 0.14, 0.21 | 0.17 (0.02) | 0.13, 0.20 | 0.16 (0.02) | 0.12, 0.19 |
| GlycA at 24y | 0.18 (0.02) | 0.14, 0.22 | 0.16 (0.02) | 0.13, 0.20 | 0.15 (0.01) | 0.12, 0.17 | 0.14 (0.01) | 0.11, 0.16 |

ALSPAC, Avon longitudinal study of parents and children; SD, Standard deviation; se, Standard error; 95% CI, 95% confidence interval; DHA, Docosahexaenoic acid; LA, Linoleic acid; Total n-3, Total omega-3; Total n-6, Total omega-6; IL-6, interleukin-6; CRP, C-reactive protein; GlycA, Glycoprotein acetyls
\* Model 2: Estimates adjusted for household social class at birth, maternal highest education qualification at birth, maternal and paternal smoking status during pregnancy, offspring sex at birth, type of drinker at age 24 years, type of smoker at 24 years, and at age in months at 24 year clinic
\*\* Model 3: As Model 2 plus additional adjustment for low density lipoprotein-cholesterol (mmol/l) and triglycerides (mmol/l) at the 24 year clinic.
\*\*\* Model 4: As Model 3 plus additional adjustment for saturated fatty acids (mmol/l) and monounsaturated fatty acids (mmol/l) at the 24 year clinic.

DHA and LA were associated with higher CRP and GlycA levels, but lower IL-6 levels after adjusting for model 2 confounders.

Total n-3 and n-6 PUFAs were associated with higher GlycA levels, but lower IL-6 levels after adjusting for model 2 confounders. Total n-6 was associated with higher CRP levels and there was no strong evidence of association between total n-3 PUFAs and CRP after adjusting for model 2 confounders. The total n-6:n-3 ratio was associated with higher levels of all three biomarkers after adjusting for model 2 confounders.

In the sex-stratified analyses, the n-6:n-3 ratio was associated with higher levels of all three biomarkers in both sexes (Figure 3 and Supplementary Table S1). PUFAs were associated with higher GlycA in both sexes. The effects of all PUFAs on CRP and IL-6 levels attenuated to the null in males. In females, findings mirrored results from the whole cohort.

Results when including all listed covariates are presented in the Supplementary materials and (Figure 2). In all analyses, including triglycerides and LDL-cholesterol (model 3) and SFAs and MUFAs (model 4) shifted the effect size towards the negative. With the exception of the n-6:n-3 ratio, the model 4 shift of effect changed the direction of effect of all PUFA measures on CRP and GlycA levels (Figure 2). Given that MUFAs and PUFAs are highly correlated, these results should be interpreted with multicollinearity in mind.

**Figure 2:**
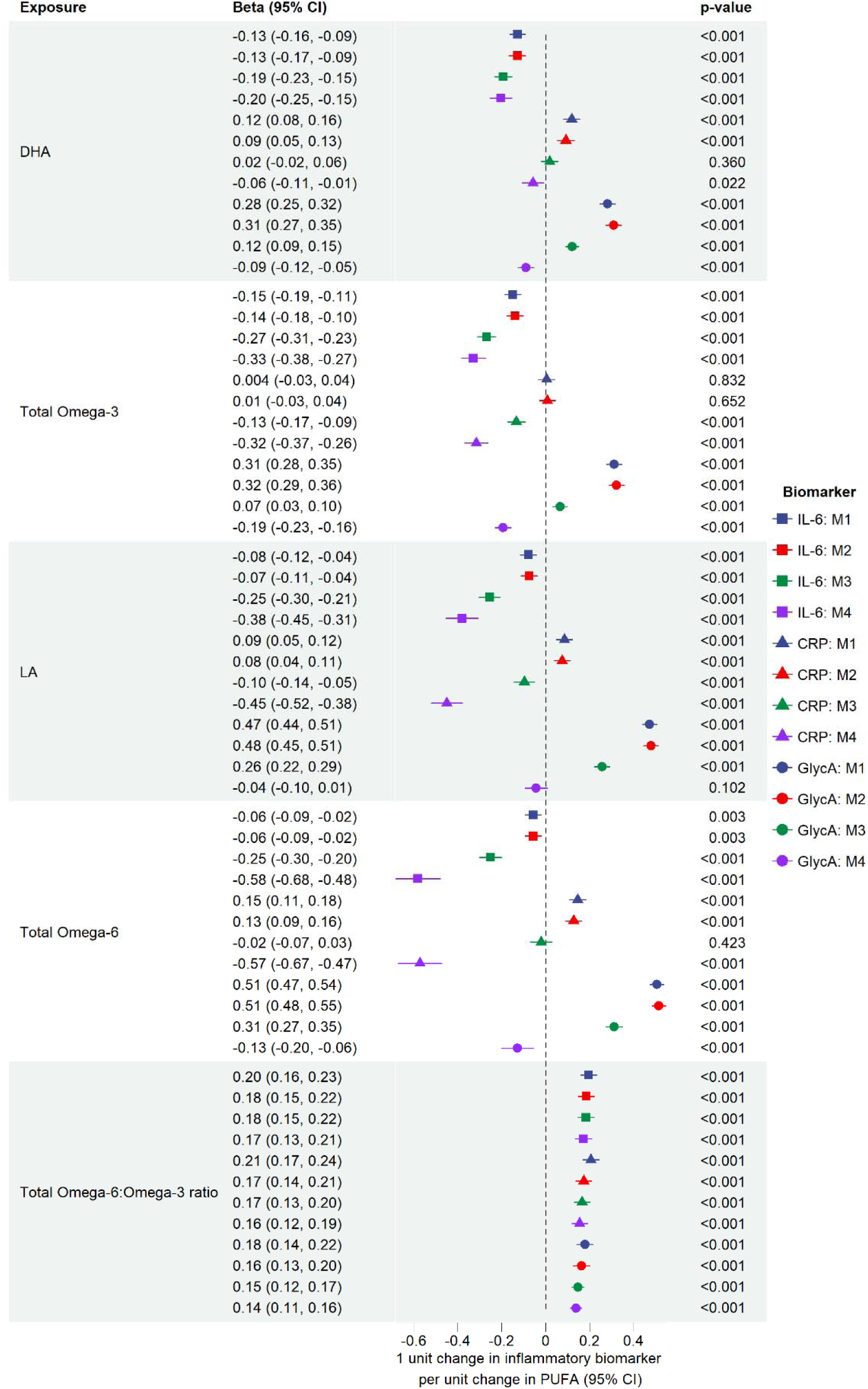
Association between fatty acids and inflammatory biomarkers using ALSPAC data at age 24 years. DHA, docosahexaenoic acid; LA, linoleic acid; IL-6, interleukin-6; CRP, C-reactive protein; GlycA, Glycoprotein acetyls; ALSPAC, Avon Longitudinal Study of Parents and Children; UKB, UK Biobank. M1:no adjustment, M2: adjustment for sex, age, maternal and paternal smoking patterns during pregnancy and SEP, M3: adjustment for covariates from M2, triglycerides and low-density lipoprotein cholesterol, M4: adjustment for covariates included in M3 and saturated fatty acids and mono-unsaturated fatty acids. ALT text: Graph depicting the association between fatty acid (FA) measures and biomarkers of inflammation (C-reactive protein, interleukin-6 and Glycoprotein Acetyls). The FA measures are: docosahexaenoic acid (DHA), linoleic acid (LA), Omega-3, Omega-6 and the ratio of Omega-6 to Omega-3. Each association is run over 4 models, where adjustment is made for additional covariates in each instance.

**Figure 3:**
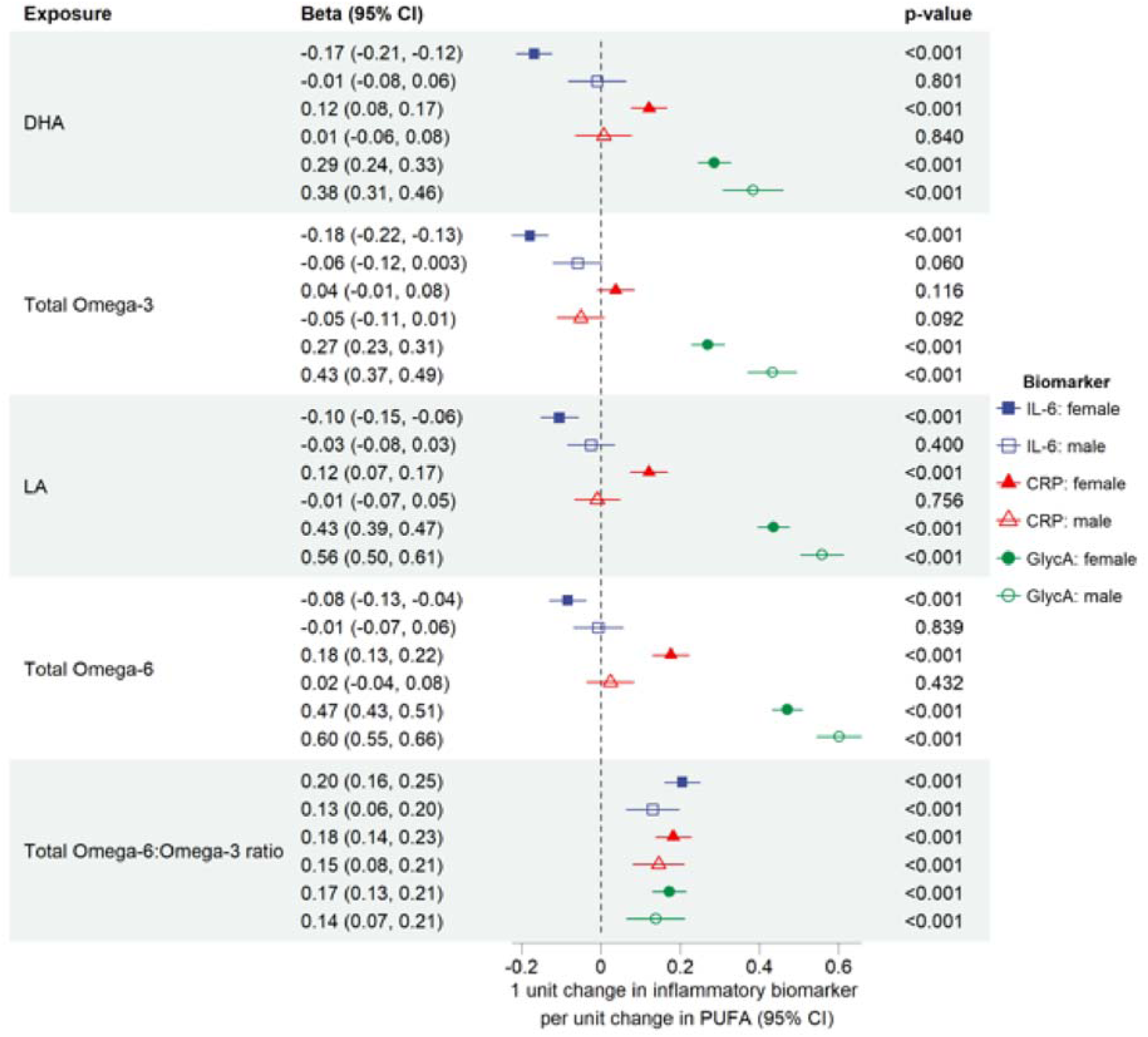
Association between fatty acids and biomarkers of inflammation using Avon Longitudinal Study of Parents and Children data at age 24 years stratified by sex at birth. DHA, docosahexaenoic acid; LA, linoleic acid; IL-6, interleukin-6; CRP, C-reactive protein; GlycA, Glycoprotein acetyls; ALSPAC, Avon Longitudinal Study of Parents and Children; PUFA, Polyunsaturated fatty acids. Analyses presented were adjusted for household social class, maternal highest education qualification, maternal and paternal smoking status during pregnancy, and participant age and smoking and drinking status when attending the ALSPAC 24-year clinic. ALT text: Graph depicting the association between fatty acid (FA) measures and biomarkers of inflammation (C-reactive protein, interleukin-6 and Glycoprotein Acetyls). The FA measures are: docosahexaenoic acid (DHA), linoleic acid (LA), Omega-3, Omega-6 and the ratio of Omega-6 to Omega-3. Each association is run for males and females separately.

Results when using UKB cohort data are presented in the Supplementary materials. Results for model 2 are consistent with findings using ALSPAC data, however when including the additional covariates (triglycerides, LDL-cholesterol, SFA and MUFA) findings diverged.

### 3.2. Results from MR Analyses

Tests investigating instrument validity indicated that instruments were unlikely to be subject to weak instrument bias (see Supplementary materials). MR results are presented in Supplementary Figure S5.

We observed no strong evidence of effect of DHA levels on CRP, GlycA or IL-6. These results were largely consistent across sensitivity analyses.

We observed no strong evidence of effect of LA levels on CRP or IL-6. However, estimates suggest that higher LA levels cause higher GlycA levels which is consistent with results from the cohort analyses. Other than in the analysis investigating the effect of LA levels on CRP, results were consistent across MR sensitivity methods.

Higher total n-3 PUFA levels were associated with higher GlycA levels (consistent with the cohort analyses). There was also evidence to suggest that higher total n-3 PUFA levels were associated with higher CRP and this was replicated in the secondary MR analysis. We found no effect of total n-3 on IL-6. Results attenuated to the null for sensitivity analyses of total n-3 PUFA levels on GlycA.

We observed no strong evidence of effect of total n-6 PUFA levels on CRP or IL-6. However, estimates did suggest that higher n-6 PUFA levels cause higher GlycA levels (consistent with cohort analyses). The MR effect estimates were fairly consistent with the main MR analysis.

Pleiotropy and heterogeneity tests are discussed in the Supplementary materials, along with the replication analysis. In short, there was no strong evidence of pleiotropy, but there was evidence of heterogeneity. Replication was fairly consistent with the main MR analysis.

### 3.4. Results from the MVMR analysis

MVMR analyses suggested that total n-6 PUFA levels independently increased both GlycA and CRP levels, but not IL-6 levels, after controlling for the effect of n-3 PUFAs. There was no direct independent effect of total n-3 PUFA levels on GlycA, CRP or IL-6 after controlling for the effect of n-6 PUFAs (Figure 4), suggesting pleiotropy may have biased the univariable MR. After further accounting for the effect of triglycerides and LDL-cholesterol, we found total n-6 PUFA levels independently increased levels of GlycA but not CRP levels. There was no independent effect of total n-3 PUFAs on any biomarker (Figure 5). A further description of the results, including the third MVMR, where triglycerides, LDL-cholesterol, SFAs and MUFAs (Figure 6) were also included, are presented in Supplementary materials along with results from MR-Lap.

**Figure 4:**
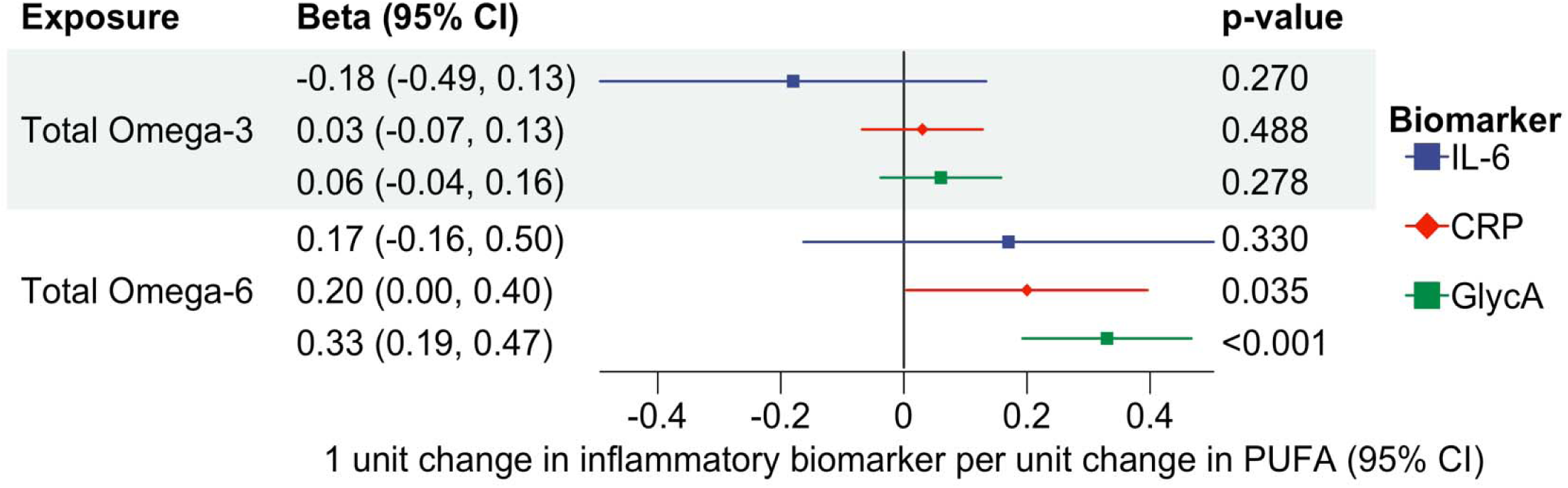
Multivariable Mendelian randomization analysis investigating the of direct effect of Omega-3 and Omega-6 on three biomarkers of inflammation (CRP, IL-6 and GlycA) independent of each other by accounting for their mutual relationships. IL-6, interleukin-6; CRP, C-reactive protein; GlycA, glycoprotein acetyls, PUFA, Polyunsaturated fatty acids. ALT text: Graph depicting the direct effect of total omega-3 and total omega-6 on biomarkers of inflammation (C-reactive protein, interleukin-6 and Glycoprotein Acetyls).

**Figure 5:**
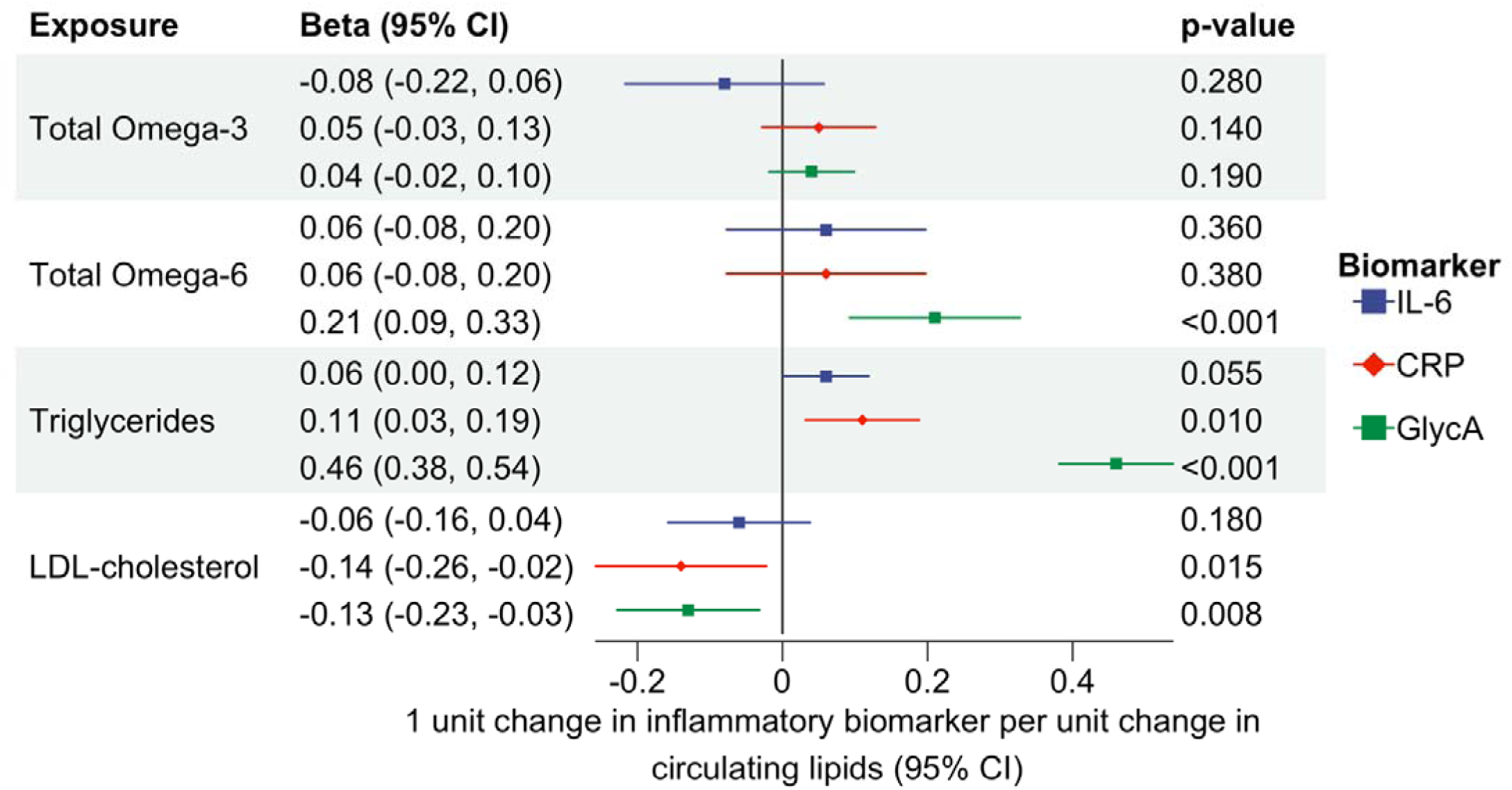
Multivariable Mendelian randomization analysis estimating the effects of omega-3, omega-6, triglycerides, and LDL-cholesterol on three biomarkers of inflammation (CRP, IL-6 and GlycA) independent of each other by accounting for their mutual relationships. IL-6, interleukin-6; CRP, C-reactive protein; GlycA, Glycoprotein acetyls; LDL-cholesterol, Low-density Lipoprotein-cholesterol; 95% CI, 95% confidence intervals. ALT text: Graph depicting the direct effect of total omega-3, total omega-6, triglycerides, and LDL-cholesterol on biomarkers of inflammation (C-reactive protein, interleukin-6 and Glycoprotein Acetyls).

**Figure 6:**
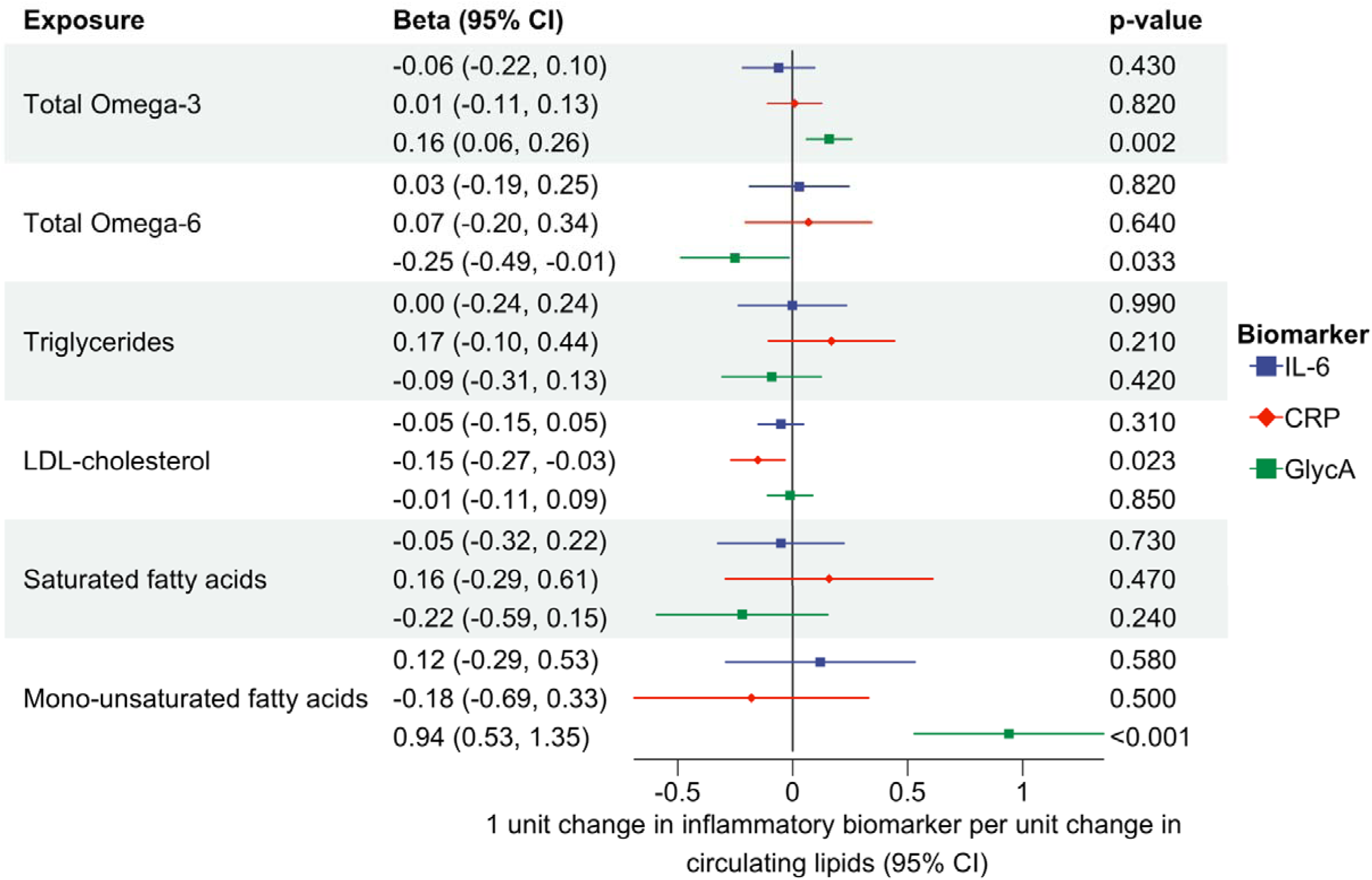
Multivariable Mendelian randomization analysis estimating the direct effects of omega-3, omega-6, triglycerides, LDL-cholesterol, saturated fatty acids and mono-unsaturated fatty acids on CRP, IL-6 and GlycA independent of each other by accounting for their mutual relationships. IL-6, interleukin-6; CRP, C-reactive protein; GlycA, Glycoprotein acetyls; LDL cholesterol, Low-density Lipoprotein-cholesterol, 95% CI, 95% confidence intervals. ALT text: Graph depicting the direct effect of total omega-3, total omega-6, triglycerides and LDL-cholesterol, saturated fatty acids, and mono-unsaturated fatty acids on biomarkers of inflammation (C-reactive protein, interleukin-6 and Glycoprotein Acetyls).

### 3.5 Results for MR analysis focusing on specific PUFA genes

MR analyses using SNPs from the *FADS* gene region suggested that DHA increased CRP levels, but that LA decreased CRP levels. *FADS* instrumented DHA and LA had no effect on GlycA or, where testable, IL-6. There was no evidence of a causal effect of any SNPs from the *ELOVL2* gene on GlycA, CRP, or IL-6. See Supplementary materials for further information.

### 3.5 Results from the MR-Clust analysis

Since SNPs can influence outcomes in distinct ways and associations between PUFAs and GlycA were consistent across cohort and MR-analyses, we used MR-Clust^(45)^ to identify SNP clusters driving heterogeneity. For LA, total n-3 and total n-6 PUFAs all clusters showed positive associations with GlycA, aligning with the main result (Supplementary Figures S6-S8). For DHA, two clusters were positively associated with GlycA, while one showed a negative association (Supplementary Figure S9). All three showed gene enrichment in lipid pathways/lipid-related GWAS. No cluster-related differentially expressed gene sets met statistical significance for tissue enrichment (Supplementary Tables S2-S6, Supplementary Figure S10). Detailed results presented in Supplementary materials.

## 4. Discussion

By combining cohort and genetic analyses, we provide insight into the relationships between PUFAs and systemic inflammation. Using multivariable linear regression, we found evidence that DHA, LA, total n-3 and total n-6 increased levels of GlycA (in ALSPAC and UKB). However, results between cohorts were less consistent for the effect of FAs on CRP. When using the ALSPAC cohort, we found a consistent positive association between the n-6:n-3 ratio and all three biomarkers, however this was not replicated in UKB.

Subsequent genetic analyses shed light into the complex nature of these associations. First, consistent with the cohort analyses, LA, total n-3 and total n-6 potentially causally increased levels of circulating GlycA which suggests a pro-inflammatory effect of these FAs. Further, as with GlycA, the MR results show total n-3 PUFAs causally increased circulating CRP levels. This contrasts the presumed anti-inflammatory effect of n-3 PUFAs. We also replicated this finding using secondary, independent GWAS data which increases confidence in this result.

Our MVMR analyses highlight the potential importance of total n-6 FAs above total n-3 FAs regarding their effect on systemic inflammation. We found, consistent with the cohort and univariable MR analysis, higher total n-6 FAs increased GlycA levels after accounting for the effect of n-3 FA levels, triglycerides and LDL-cholesterol. Therefore, as suggested by previous literature, n-6 FAs may increase levels of inflammation. However, total n-3 FAs had no effect on GlycA or CRP after accounting for total n-6 FAs, triglycerides and LDL-cholesterol which bring into question the use of n-3 supplementation as a method to reduce systemic inflammation. However, the biomarkers measure different aspects of inflammation (acute versus low-grade chronic inflammation) and the PUFAs affect them differentially. By only using three biomarkers of inflammation, we may have missed important effects PUFAs have on inflammation. Research using additionally biomarkers or inflammatory networks is needed to better understand the relationship between PUFA consumption and systemic inflammation.

Results from our MR analyses using genetic variants within/nearby the *FADS* gene cluster differed from the main analysis, indicating that the original results may be biased by pleiotropy, rather than reflecting the true effect of FA biosynthesis. Even when using genetic variants that fall within the *FADS* gene region, which is key for FA metabolism, the instruments may lack specificity because they are also enriched in genes involved in actions such as lipoprotein metabolism^(43)^. We aimed to mitigate this problem through the inclusion of lipoproteins in an MVMR, however they too may be impacted by horizontal pleiotropy.

A major strength of our study is that we investigated the association between PUFAs and levels of CRP, IL-6 and GlycA, using two different methods, replicated the methods using different cohorts and ran further sensitivity analyses to address potential biases that arise in epidemiological research.

Despite this, we recognize several limitations. Using cross-sectional analyses means we could not determine the temporal or causal relationship between the exposure and outcome. However, it does provide useful context and evidence for associations between PUFA levels and biomarkers of inflammation, including GlycA, which is novel.

DHA and LA appear at different ends of FA biosynthesis pathways (DHA is a product of n-3 desaturation/elongation reactions, while LA is a n-6 precursor), creating difficulties with direct comparisons. Additionally, the NMR platform did not measure all FAs, meaning key inflammatory metabolites may have been missed. Although our results suggest total n-6 increases inflammation, the limited NMR coverage and pleiotropic nature of FA genetic instruments prevent us from confidently identifying the specific classes of FAs driving these associations. Conditioning on total fatty acid levels could introduce collider bias and therefore we did not use FAs reported as a proportion of total FA. Subsequently, the relative abundance of each FA may be obscured, and it is harder to interpret the significance of individual FAs. Further, in some cases, reporting FAs individually may not provide enough context about the overall composition of the individual’s blood such as overall lipid profile. Despite this, reporting individual FAs was informative as this is the first study on their causal link to inflammation.

Another limitation is that inflammation is a latent trait inferred through various markers. Circulatory protein levels may not fully capture its biological complexity. Although we failed to observe anti-inflammatory properties, our findings do not rule out anti-inflammatory effects of n-3PUFAs and work is needed to understand this multifaceted relationship. However, given the focus of this work was to examine the relationship between PUFAs and systemic inflammation, CRP, IL-6 and GlycA are appropriate as they reflect broad inflammatory activity.

Lastly, although the *FADS* gene cluster encodes enzymes fundamental to PUFA biosynthesis suggesting that IVs within this region are more likely to satisfy MR assumptions, *FADS* variants have been shown to be highly pleiotropic^(46)^ and do not differentiate n-3 or n-6 effects. As such, our gene based analyses may capture pathways to inflammation through factors other than PUFAs, thus violating the exclusion-restriction assumption^(28)^.

Given the popularity of n-3 PUFAs supplements^(47–49)^, our results are of public health importance. Replication could shift messaging toward reducing total n-6 PUFA consumption and promoting a healthy, balanced n:6:n3 ratio to lower inflammation. However, we acknowledge that n-3 PUFAs have benefits beyond their posited anti-inflammatory effects, e.g., hypotriglyceridaemic properties^(50)^ and that some anti-inflammatory effects may not have been captured by our biomarkers. A better understanding of n-6 and n-3 interactions is needed before policy change.

## Supporting information

Supplement documentation

## Ethics

Ethical approval for the study was obtained from the ALSPAC Ethics and Law Committee and the Local Research Ethics Committee and can be found here: http://www.bristol.ac.uk/alspac/researchers/research-ethics/. Informed consent for the use of data collected via questionnaires and clinics was obtained from participants following the recommendations of the ALSPAC Ethics and Law Committee at the time. Consent for biological samples has been collected in accordance with the Human Tissue Act (2004).

UK Biobank has approval from the North West Multi-centre Research Ethics Committee (MREC) as a Research Tissue Bank (RTB).

## Data availability

Data needed to evaluate the conclusions presented in this paper are provided in the manuscript and/or the supplementary material. Additionally, ALSPAC data can be requested from the ALSPAC executive committee and reasonable requests from bona fide researchers. GWAS data is publicly available using the OpenGWAS website (https://gwas.mrcieu.ac.uk).Part of this research was conducted using data from UK Biobank project ID:81499 and project: 30418, a major biomedical database and can be provided by UKB (http://www.ukbiobank.ac.uk/).

Code for data management and statistical analysis has been made available in <u>daisycrick</u> <u>(github.com)</u>. Cohort analyses were performed using STATA version 17.0. All other analyses were performed in R Software version 4.1.0.

## Contributions

*Daisy C.P. Crick:* Conceptualization, Methodology, Investigation, Writing – original draft, Writing – review & editing. *Hannah Jones:* Writing – Conceptualization, Supervision, Methodology, review & editing. *Sarah Halligan:* Supervision, Writing – review & editing. *Golam M. Khandaker:* Conceptualization, Methodology, Supervision, Writing – review & editing. *George Davey Smith:* Methodology, Writing – review & editing. Correspondence to Daisy Crick

## Use of Artificial intelligence tools

None.

## Acknowledgements

This work was carried out using the computational facilities of the Advanced Computing Research Centre, University of Bristol - http://www.bristol.ac.uk/acrc/.

We are extremely grateful to all the families who took part in this study, the midwives for their help in recruiting them, and the whole ALSPAC team, which includes interviewers, computer and laboratory technicians, clerical workers, research scientists, volunteers, managers, receptionists, and nurses.

