## Supplement documentation for "The Relationship between Polyunsaturated Fatty Acids and Inflammation: Evidence from cohort and Mendelian randomization analyses"

**Investigating the Relationship between Polyunsaturated Fatty Acids and Inflammation using Cohort and Two-sample Mendelian Randomization Analyses - Supplementary Material**

**Table of Contents**

***1.***	***eMethods	3***

**1.1.**	**Mendelian randomization	3**

**1.2.**	**ALSPAC recruitment details	4**

**1.3.**	**UK Biobank recruitment details	5**

**1.4.**	**Blood sampling and data collection	5**

1.4.1.	ALSPAC	5

1.4.2.	UK Biobank	6

**1.5.**	**Covariates	6**

1.5.1.	ALSPAC	6

1.5.2.	UK Biobank	7

**1.6.**	**Data cleaning	7**

1.6.1.	ALSPAC	7

1.6.2.	UKB	8

**1.7.**	**Multiple Imputation	8**

**1.8.**	**Generating genetic instruments	8**

**1.9.**	**Instrument Selection for the primary univariable MR	9**

**1.10.**	**Methods to investigate pleiotropy and heterogeneity	9**

**1.11.**	**Multivariable mendelian randomization analysis	10**

**1.12.**	**MR-Lap analysis to investigate bias from sample overlap, weak instrument and winner’s curse	11**

**1.13.**	**Instrument selection for specific PUFA genes	11**

**1.14.**	**MR-Clust analysis to explore heterogeneity in MR instruments	13**

**1.15.**	**Functional annotation of loci identified within clusters of interest	13**

***2.***	***eResults	14***

**2.1.**	**Instrument validity	14**

**2.2.**	**Results from investigating the direct effects of total n-3 and total n-6 on inflammatory markers using multivariable Mendelian randomization	14**

2.2.1.	Adjusting for total n-3 and total n-6	14

2.2.2.	Adjusting for total n-3, total n-6, LDL and triglycerides	16

2.2.3.	Adjusting for triglycerides, LDL-cholesterol, MUFAs and SFAs	17

**2.3.**	**Results from investigating biases impacting MR using MR-Lap	17**

2.3.1.	Potential causal effect of DHA levels on inflammatory markers	18

2.3.2.	Potential causal effect of LA levels on inflammatory markers	18

2.3.3.	Potential causal effect of n-3 levels on inflammatory markers	19

2.3.4.	Potential causal effect of n-6 levels on inflammatory markers	19

**2.4.**	**Results from MR analysis focusing on specific PUFA genes	20**

**2.5.**	**Results from MR-Clust analysis to explore heterogeneity in MR instruments	20**

**2.6.**	**Functional annotations of DHA cluster SNPs using FUMAGWAS	21**

**2.7.**	**Secondary MR analysis and results	21**

***3.***	***eReferences	24***

### 1. eMethods

#### 1.1. Mendelian randomization

Mendelian Randomisation (MR)^[1, 2]^ is an instrumental variable analysis that uses the random assortment of genetic variants from parents to offspring to investigate the causal effect of an exposure on an outcome^[3]^. MR is bound by three assumptions (i) the relevance assumption, which describes how the genetic variants used to instrument the exposure of interest are statistically strongly associated with the exposure of interest and relevant to the population to which inference is being made; (ii) the independence assumption, which suggests that there is no confounding of the genetic instrument-outcome association; and (iii) the exclusion-restriction assumption, which suggests that the effect of the genetic instrument on the outcome is only via the exposure of interest. When all three assumptions are met MR can overcome issues of reverse causation and unmeasured confounding. As such, conventional multivariable regression analysis and MR have differing and unrelated sources of bias. Therefore, the methods can be used together to triangulate evidence and, when results are concordant, support causal inference ^[4]^.

#### 1.2. ALSPAC recruitment details

The Avon Longitudinal Study of Parents and Children (ALSPAC) birth cohort recruited pregnant women resident in Avon, UK with expected dates of delivery between 1st April 1991 and 31st December 1992 ^(5-7)^. There were 14,203 unique mothers initially enrolled in the study and 13,988 children were alive at 1 year of age. When the oldest children were approximately 7 years of age, the initial sample was bolstered with eligible cases who did not originally join the study. The total sample size for analyses using any data collected after the age of seven is therefore, 14,833 unique mothers and 15,447 pregnancies which resulted in 15,658 foetuses. Of these, 14,901 were alive at 1 year of age. The offspring, their mothers and the mother’s partners are regularly followed up.

Please note that the ALSPAC website contains details of all the data that is available through a fully searchable data dictionary and variable search tool http://www.bristol.ac.uk/alspac/researchers/our-data/. Ethical approval for the study was obtained from the ALSPAC Ethics and Law Committee and the Local Research Ethics Committee and can be found here: <http://www.bristol.ac.uk/alspac/researchers/research-ethics/>. Informed consent for the use of data collected via questionnaires and clinics was obtained from participants following the recommendations of the ALSPAC Ethics and Law Committee at the time. Consent for biological samples has been collected in accordance with the Human Tissue Act (2004). Study data were collected and managed using REDCap electronic data capture tools hosted at the University of Bristol. REDCap (Research Electronic Data Capture) is a secure, web-based software platform designed to support data capture for research studies^[5]^.

Details regarding sample processing and NMR analysis have been provided elsewhere^[6-8]^.

#### 1.3. UK Biobank recruitment details

UK Biobank (UKB) recruited ~500,000 participants and baseline assessments were completed between 2006-2010. Participants were aged 40-70 years at baseline, registered with a general practitioner and lived close to 22 assessment centres in England, Scotland, and Wales. Baseline assessments included demographics, lifestyle, and disease history, with linkages to electronic medical records. UK Biobank’s ethical approval was from the Northwest Multi-centre Research Ethics Committee.

Participants were included if they had exposure (docosahexaenoic acid [DHA], linoleic acid [LA], total omega-3 PUFAs (n-3) and total omega-6 PUFAs [n-6]) and outcome (C-Reactive Protein [CRP] and Glycoprotein Acetyls [GlycA] levels) data available at 24y. We excluded individuals of non-white European ancestry giving a total of 12,451 individuals (45.48% males). Information on IL-6 was not available.

#### 1.4. Blood sampling and data collection

##### 1.4.1. ALSPAC

Participants fasted overnight, or >6 hours if being seen in the afternoon, before attending the clinic for blood sampling at 24 years old. Blood samples were centrifuged immediately to isolate plasma and stored at -80 °C. There were no freeze-thaw cycles during storage. Total n-3, n-6, DHA and LA PUFA levels and plasma GlycA levels were measured using a high-throughput proton (1H) Nuclear Magnetic Resonance (NMR) metabolomics platform (Nightingale, UK). We also created a n-6:n-3 ratio (total n-6 PUFAs divided by total n-3 PUFAs). CRP was measured by automated particle-enhanced immunoturbidimetric assay (Roche UK, UK). IL-6 was measured by enzyme-linked immunosorbent assay (OLINK, UK).

GlycA levels ranged from 0.84 to 2.25 mmol/L, IL-6 levels ranged from 1.82 to 10.60 Normalized Protein eXpression (NPX) Log2 scale and CRP levels ranged from 0.15 to 70.05 mg/L.. CRP had detection limits of 0.15-80mg/L meaning measures outside this were removed. CRP was converted from mg/L to mmol/L so as to be in the same units as GlycA.

##### 1.4.2. UK Biobank

Blood samples were collected at the baseline clinic (mean age: 48y) and GlycA and PUFA measures was quantified using the same method as in ALSPAC. Serum CRP was measured by automated particle-enhanced immunoturbidimetric assay (Beckman Coulter (UK), Ltd) with detection limits of 0.08 - 80mg/L, however in accordance with the ALSPAC analysis, measures outside 0.15-80mg/L were removed. CRP was converted from mg/L to mmol/L so as to be in the same units as GlycA. Full details regarding CRP sample and data processing can be found elsewhere [9] and on the UKB website (<https://www.ukbiobank.ac.uk/enable-your-research/about-our-data/biomarker-data>).

#### 1.5. Covariates

##### 1.5.1. ALSPAC

Childhood socio-economic position (SEP) was indexed using maternal self-reported highest education qualification at 8-42 weeks gestation (dichotomised according to presence/absence of a degree level qualification); and the highest social occupation of either parent (dichotomised according to manual/non-manual occupation). Maternal and paternal smoking during pregnancy (categorised as no smoking: no reports of smoking in any trimester and, any smoking: reports of smoking in any trimester) was used as a proxy for general pregnancy health. Participants’ sex at birth was rded and age in months at the 24 year ALSPAC clinic, participants’ smoking status (categorised as current non-smoker or current smoker) and drinking status (categorised as none/infrequent or frequent) were reported at the assessment clinic. Blood samples collected at age 24y (described above) were used to quantify triglyceride levels, saturated fatty acid levels and mono-unsaturated fatty acid levels. These covariates were selected *a priori* based on known or plausible relationships with both systemic inflammation and polyunsaturated fatty acids (PUFAs).

##### 1.5.2. UK Biobank

We used own highest educational qualification as a measure of SEP which was dichotomised according to the presence/absence of a degree level qualification at baseline. To index maternal smoking during pregnancy, participants were asked at baseline ‘Did your mother smoke regularly around the time when you were born?’ (yes/no), where answers in the affirmative were taken to proxy maternal smoking during pregnancy. Participants’ sex and age (in years), smoking status and drinking status (categorised as never or ever) were self-reported at baseline. Blood samples collected at baseline (described above) were used to quantify triglyceride levels, saturated fatty acid levels and mono-unsaturated fatty acid levels.

#### 1.6. Data cleaning

##### 1.6.1. ALSPAC

CRP and IL-6 levels were not normally distributed and were log-transformed. To help with the interpretation of results, we z-transformed both exposure and outcome data, and so the effect estimates represent the increase in outcome in standard deviation (SD) per SD increase in exposure.

##### 1.6.2. UKB

Exposure data underwent an inverse normal transformation. As with the ALSPAC analysis, to help with interpretation of results, exposure and outcome data were z-transformed. CRP was not normally distributed and was log-transformed.

#### 1.7. Multiple Imputation

We performed multiple imputation in ALSPAC only, given that a) it was our primary cohort analysis sample and b) to bolster the limited sample size. We created 16 datasets with 160 iterations which aimed to balance computational efficiency while ensuring consistency of results^[9]^. The main and auxiliary variables used to inform the imputation are listed in Supplementary Table S7. Imputation was run in Stata version 17.0 (Stata Corp, College Station, TX) and estimates were then combined through Rubin's rules. We compared imputed vs non-imputed characteristics to check the imputation model.

#### 1.8. Generating genetic instruments

Genetic variant (single nucleotide polymorphism [SNP]) instrument sets were generated for each fatty acid phenotype of interest, namely circulating levels of DHA, LA, total n-3 PUFAs and total n-6 PUFAs, using summary level data from European population genome-wide association studies (GWAS) which used data from UK Biobank (UKB; n=114,999) ^[10]^. For the outcome variables, we used SNPs identified in summary-level data from European population GWAS of the inflammatory biomarkers GlycA^[11]^, CRP^[12]^ and IL-6^[13]^. The GlycA GWAS used data from UKB (n=114,999), the serum CRP GWAS used data from UKB and the Cohort for Heart and Aging Research in Genomic Epidemiology (CHARGE; n=575,531)^[14]^ and the IL-6 GWAS used data from a meta-analysis of 26 cohorts with a total of 52,654 individuals^[13]^ . Ethical approval was obtained in all original studies.

#### 1.9. Instrument Selection for the primary univariable MR

Using the TwosampleMR R package, fatty acid SNP instrument information (e.g. SNP alleles, effect allele frequency etc.) was harmonised with the outcome biomarker information meaning that the SNP-exposure and SNP-outcome effects corresponded to the same allele. During harmonization, palindromic SNPs with intermediate allele frequencies of above 0.48 and below 0.52 were considered strand ambiguous and removed and SNPs that were unavailable in the outcome datasets were replaced by proxy SNPs at R^2^>0.80. Further, SNPs that had a minor allele frequency of 0.01 or less were excluded and we ensured that SNPs were independent through clumping the harmonized SNPs for linkage disequilibrium (LD, r2=0.001). Not all SNPs were available in every GWAS which lead to a different number of available SNPs for each analysis. Steiger filtering was then applied, meaning SNPs were removed if they explained more variance in the outcome than in the exposure. The number of SNPs removed due to being palindromic, in linkage disequilibrium (LD) or as a result of Steiger filtering for each analysis is reported in Supplementary Table S8.

There were 46-58 SNPs available to investigate the effect of PUFAs on GlycA levels, 17-39 SNPs available to investigate the effect of PUFAs on CRP levels and 11-44 SNPs available to investigate the effect of PUFAs on IL-6 levels. We report F-statistics for each IV, as a measure of instrument strength.Harmonized SNP information for each relationship is presented in Supplementary Tables S9-S20.

#### 1.10. Methods to investigate pleiotropy and heterogeneity

Presence of heterogeneity between the SNP effect estimates was assessed using Cochran’s Q test in the IVW analysis and the Rucker’s Q test in the MR-Egger analyses. We used the MR-Egger intercept and MR Pleiotropy Residual Sum and Outlier global test (MR-PRESSO) to investigate the presence of pleiotropy. See Supplementary Table S21 for a description of MR methods and sensitivity analyses as described in a previous paper ^[15]^.

#### 1.11. Multivariable mendelian randomization analysis

MVMR^[16]^ is an extension of MR that estimates the *direct effect* of each exposure on the outcome, rather than the overall total effect of the exposures (Supplementary Figure S11). As stated in the manuscript, we conducted three MVMR analyses to estimate the independent direct causal effect of total n-3 and n-6 PUFAs on the biomarkers of inflammation. The first, including only total n-3 and total n-6 as exposures, the second including total n-3, total n-6, triglycerides and LDL-cholesterol and the third including total n-3, total n-6, triglycerides, LDL-cholesterol, SFA and MUFAs.

Linkage disequilibrium score regression (LDSC) estimated the genetic correlation between total n-3 and total n-6 (0.334) which confirmed our assumption that they may act as a pleiotropic pathway. We used the same GWAS data as in the univariable MR. SNP instrument sets were generated for LDL and triglycerides using summary level data from European population genome-wide association studies (GWAS) which used data from UK Biobank (n= 440,546, and n= 441,016 respectively) ^[17]^.Iinstrument sets for MUFAs and SFAs using summary level data from European population genome-wide association studies (GWAS) which used data from UK Biobank (UKB; n=115,006) ^[18]^.

In all three MVMR analyses, the same thresholds for genome-wide significance, LD and palindromic SNPs were applied as in the univariable MR.

For the first GWAS, the n-3 SNPs and n-6 SNPs were combined, giving a final exposure instrument of 158 SNPs. The exposure instrument was then harmonised with the outcome data (GlycA/CRP/IL-6). After harmonization 31 SNPs were available to investigate the direct effect of total n-3 and total n-6 PUFAs on the outcome CRP, 78 SNPs were available to investigate the direct effect of total n-3 and total n-6 PUFAs on the outcome GlycA and 9 SNPs were available to investigate the direct effect of total n-3 and total n-6 PUFAs on IL-6.

For the second GWAS, n-3 SNPs, n-6 SNPs, triglyceride SNPs and LDL-cholesterol SNPs were combined to give a final exposure instrument of 1308 SNPs. The exposure instrument was then harmonised with the outcome data (GlycA/CRP/IL-6). After harmonization there were 282 SNPs for the exposures instrument on the outcome CRP, 319 SNPs for the exposure instrument on the outcome GlycA and 122 SNPs for the exposures instrument on IL-6.

For the third GWAS, the n-3 SNPs, n-6 SNPs, triglyceride SNPs, LDL-cholesterol SNPs, MUFA SNPs and SFA SNPs were combined, giving a final exposure instrument of 1974 SNPs. The exposure instrument was then harmonised with the outcome data (GlycA/CRP/IL-6). After harmonization there were 467 SNPs for the exposure instrument on the outcome CRP, 518 SNPs for the exposure instrument on the outcome GlycA and 165 SNPs for the exposure instrument on IL-6.

#### 1.12. MR-Lap analysis to investigate bias from sample overlap, weak instrument and winner’s curse

MR-Lap assesses potential bias in MR effect estimates which can arise due to sample overlap, weak instrument bias and winner’s curse^[19]^. If the MR-Lap corrected effect does not differ from the observed effect, then the IVW-MR estimate can be used. If there is a difference between the two estimates, it suggests that the IVW estimates may be biased and therefore, the MR-Lap corrected effect would be preferred ^[19]^. For this analysis, we used the same PUFA and inflammatory biomarker GWAS’ as used in the main analysis. Instrumental variables (IVs) were pruned to the distance threshold of 10000 Kb with an LD threshold of 0.01.

#### 1.13. Instrument selection for specific PUFA genes

Instrument variables which fell in/near (+/- 500kb) to the genes that encode the key fatty acid desaturase enzymes (FADS gene cluster of *FADS1*, *FADS2* and *FADS3*) (chromosome 11: 61 560 452–61 659 523) and elongase enzymes *ELOVL2* gene locus *(*chromosome 6: 10 980 992–11 044 624), were extracted from the DHA GWAS and the LA GWAS summary statistics.

A total of 2026 independent SNPs reached genome-wide significance (P< 5∙0 x10-8) within the *FADS* gene cluster of the DHA GWAS and the LA GWAS. However, after clumping using a European population reference panel to remove SNPs in LD, only 11 genome-wide significant SNPs remained from the DHA GWAS and 7 genome-wide significant SNPs remained available from the LA GWAS.

A total of 5399 SNPs reached genome-wide significance within the *ELOVL2* gene of the DHA GWAS and the LA GWAS. However, after clumping only 9 genome-wide significant SNPs remained available from the DHA GWAS and 17 genome-wide significant SNPs remained available from the LA GWAS.

SNPs were harmonized, aligning the genetic association for exposure and outcome on the effect allele using the effect allele frequency. As in the primary analysis, palindromic SNPs with intermediate allele frequencies of above 0.48 and below 0.52 were considered strand ambiguous and removed. SNPs that were unavailable in the outcome datasets were replaced by proxy SNPs at R^2^>0.80. Further, SNPs that had a minor allele frequency of 0.01 or less were excluded and we ensured that the SNPs were independent by clumping the harmonized SNPs for LD (r2=0.001). Steiger filtering was applied. The number of available SNPs for each analysis are presented in Supplementary Table S22.

Following harmonization, there were 1-11 SNPs available to investigate the effect of the *FADS* gene cluster from the DHA GWAS on CRP, GlycA and IL-6 levels. There were 6 and 7 SNPs respectively available to investigate the effect of the *FADS* gene cluster from the LA GWAS on CRP and GlycA levels, however when using the outcome IL-6 GWAS no SNPs were available post harmonization.

When using SNPs identified to be within the *ELOVL2* gene, following harmonization, there were 2-7 SNPs available from the DHA GWAS to investigate the effect of the *ELOVL2* gene on CRP, GlycA and IL-6 levels. There were 1-14 SNPs available to investigate the effect of the *ELOVL2* gene from the LA GWAS on CRP, GlycA and IL-6 levels. Harmonized SNP information for each relationship is presented in Supplementary Tables S23-S24.

#### 1.14. MR-Clust analysis to explore heterogeneity in MR instruments

SNPs can influence the outcome in distinct ways which can lead to high levels of heterogeneity. Therefore, when results between cohort and MR analyses were consistent, but the latter had strong evidence of heterogeneity we used MR-Clust^[20]^ to investigate whether individual causal estimates fell into distinct clusters based on effect magnitude. Gene mapping and gene set and tissue enrichment analyses were performed using FUMA v1.5.2 (<https://fuma.ctglab.nl>) for clusters of interest ^[21]^. See further details in eMethods.

Using the author’s guidelines we opted to use the conservative method, whereby only variants were assigned to a cluster if the conditional probability of cluster assignment was ≥ 0.89 and clusters were only recognised if they contained at least four variants. Variants that did not satisfy these criteria and did not fall into a null cluster were grouped into a “junk” cluster. The null cluster was a collection of variants that did not have an effect on the outcome.

#### 1.14.1. Functional annotation of loci identified within clusters of interest

Following MR-Clust, functional annotations of the genomic loci within clusters of interest were obtained using FUMA v1.5.2 (<https://fuma.ctglab.nl>) ^[22]^. First, the SNP2GENE function was used to map SNPs within each cluster, as well as SNPs correlated with these SNPs at *r^2^* ≥ 0.6, to prioritized genes. The GENE2FUNC function was then used for gene set enrichment analyses using hypergeometric tests to test for overrepresentation of biological functions using gene sets obtained from MsigDB (i.e., hallmark gene sets, positional gene sets, curated gene sets, motif gene sets, computational gene sets, GO gene sets, oncogenic signatures, and immunologic signatures) and WikiPathways. The set of background genes (i.e., the genes against which the set of prioritized genes are tested against) consisted of 20,260 protein-coding genes. Multiple testing correction (Benjamini–Hochberg false discovery rate) was performed per data source of tested gene sets. Prioritized genes were also tested against differentially expressed gene (DEG) sets using the hypergeometric test to test for enrichment in genes expressed in 54 tissue types from GTEx v8.

### 2. eResults

#### 2.1. Cohort analysis

When using ALSPAC data, there was no difference between the complete-case and imputed results (Supplementary Table S25). Distributions of the observed and imputed characteristics are presented in Supplementary Table S26. Distribution characteristics observed using UKB are presented in Supplementary Table S27.

#### 2.1.1. Results for additional analyses using ALSPAC cohort data

As in Model 2, where associations were adjusted for sex, age, maternal and paternal smoking patterns during pregnancy and SEP, DHA, LA, total n-3 and total n-6 were associated with lower levels of IL-6 in model 3 (adjusting for sex, age, maternal and paternal smoking patterns during pregnancy, SEP triglycerides and LDL-cholersterol) and in model 4 (adjusted for sex, age, maternal and paternal smoking patterns during pregnancy, SEP, triglycerides, LDL-cholesterol, SFAs and MUFAs). Additionally, the total n-6:n-3 ratio was associated with higher levels of all three biomarkers in model 3 and 4.

In model 2, DHA, LA and total n-6 was associated with higher levels of GlycA and CRP. However, in model 3 and 4, the effect of DHA on CRP attenuated to the null and LA, total n-3 and total n-6 were associated with lower CRP. We also found that in model 3, the effect of DHA, LA, total n-3 and total n-6 on GlycA remained consistent with model 2. However, after the introduction of SFAs and MUFAs, we observed a negative effect of DHA and total n-3 on GlycA and no effect of LA or total n-6 on GlycA.

This reversal of effect may be a consequence of multicollinearity as MUFAs and PUFAs are highly correlated. The finding further supports the use of model 2 as our main finding. Results are presented in Table 2.

#### 2.1.2. Results for secondary analysis using UK Biobank cohort data

When using data from UKB and, consistent with the results from ALSPAC, we found that DHA, LA, total omega-3, total omega-6 and the total n-6:n-ratio were positively associated with GlycA (Supplementary Table S28). However, unlike when using ALSPAC, we found no association between DHA, LA, total n-6 or the total n-6:n-3 ratio and CRP. We were unable to investigate associations between PUFAs and IL-6 due to data availability. Sex stratified results are presented in Supplementary Table S29. Results for the secondary analysis, including the additional covariates triglycerides, LDL-cholesterol, SFA and MUFA were not consistent with result when using ALSPAC data and are presented in the supplement (Supplementary Table S28).

#### 2.2. Instrument validity

Within the univariable analysis, SNP-level F statistics were all >10, indicating that IVW estimates were likely not subject to weak instrument bias. The total variance explained (R^2^) by each SNP in the univariable MR analysis was between 0.03% and 4.03% for DHA, 0.03% and 0.55% for LA, 0.02% and 5.21% for total n-3 PUFAs, and 0.02% and 0.62% for total n-6 PUFAs. Genetic instruments, sample sizes and instrument F-statistics for each GWAS are presented in Supplementary Table S30. All results are presented after Steiger filtering.

#### 2.3. Results from investigating the direct effects of total n-3 and total n-6 on inflammatory markers using multivariable Mendelian randomization

##### 2.3.1. **Adjusting for total n-3 and total n-6**

We conducted a Multivariable Mendelian Randomization (MVMR) analysis to estimate the individual direct causal effects of total n-3 and n-6 PUFAs on the biomarkers of inflammation independently of each other. We observed a positive direct effect of total n-6 PUFA levels on both GlycA (IVW estimate: 0.33; 0.19, 0.47) and CRP levels (IVW estimate: 0.20; 0.01, 0.39), but not IL-6 levels (IVW estimate: 0.17; -0.17, 0.50), after controlling for the effect of n-3 PUFAs. There was no direct effect of total n-3 PUFA levels on GlycA (IVW estimate: 0.06; -0.05, 0.16), CRP (IVW estimate:0.03; -0.06, 0.12) or IL-6 (IVW estimate: -0.18; -0.49, 0.14) levels after controlling for the effect of n-6 PUFAs. This suggests that the effect of total n-3 PUFAs found in the univariable MR analysis are not independent of the effect of n-6, and may be a result of pleiotropy (Figure 6).

There was evidence of heterogeneity when investigating the direct effect of total n-3 PUFAs and total n-6 PUFAs on GlycA (Q=1504.022, p=<0.001), CRP (Q=1296.58, p=<0.001) but no evidence for heterogeneity IL-6 (Q=10.36, p=0.110).

There was no evidence of weak instrument bias when using the overall conditional F-statistic for instrument strength for GlycA (n-3 PUFAs (F=67.71) and total n-6 PUFAs (F=99.85)), CRP (n-3 PUFAs (F=61.20) and total n-6 PUFAs (F=161.65)) or IL-6 (total n-3 PUFAs (13.55) and total n-6 PUFAs (12.17))

##### 2.3.2. **Adjusting for total n-3, total n-6, LDL and triglycerides**

We observed a positive direct effect of total n-6 PUFA levels on GlycA (IVW estimate: 0.21; 0.10, 0.33), but not CRP levels (IVW estimate: 0.06; -0.07, 0.19) or IL-6 levels (IVW estimate: 0.06; -0.07, 0.20), after controlling for the effect of n-3 PUFAs, LDL-cholesterol and triglycerides. There was no direct effect of total n-3 PUFA levels on GlycA (IVW estimate: 0.04; -0.02, 0.11), CRP (IVW estimate:0.05; -0.02, 0.13) or IL-6 (IVW estimate: -0.08; -0.21, 0.06) levels after controlling for the effect of n-6 PUFAs. There was evidence that LDL cholesterol independently reduced levels of CRP (-0.14; -0.25, -0.03) and GlycA (-0.13; -0.23, -0.03), but had no independent effect of IL-6 (-0.06; -0.16, 0.03). Additionally, triglycerides appeared to independently increase levels of CRP (0.11; 0.03, 0.20) and GlycA (0.46; 0.39, 0.53), but had no independent effect on IL-6 (0.06; 0.00, 0.12). Results are presented in Figure 5.

There was evidence of heterogeneity when investigating the direct effect of total n-3 PUFAs and total n-6 PUFAs on GlycA (Q=2284.98, p=<0.001), CRP (Q=10395, p=<0.001), but no evidence for heterogeneity for IL-6 (Q=10.36, p=0.110).

When investigating the effect of the exposures on GlycA, the overall conditional F-statistic for instrument strength was larger than the conventional threshold of 10 for LDL (13.50), triglycerides (40.11) and total n-3 PUFAs (F=17.06). However, there was evidence of weak instruments for total n-6 PUFAs (F=6.88).

When investigating the effect of the exposures on CRP the overall conditional F-statistic for instrument strength was larger than the conventional threshold of 10 for LDL (14.68), triglycerides (41.65) and total n-3 PUFAs (F=18.04). However, there was evidence of weak instruments for total n-6 PUFAs (F=7.48).

When investigating the effect of the exposures on IL-6 the overall F-statistic for instrument strength was lower than the conventional threshold of 10 for all exposures: total n-3 PUFAs (1.65), total n-6 PUFAs (1.56), LDL-cholesterol (3.34) and triglycerides (6.07) which suggests that the result may be biased by weak instruments.

##### 2.3.3. **Adjusting for triglycerides, LDL-cholesterol, MUFAs and SFAs**

After the additional adjustment for triglycerides, LDL-cholesterol, MUFAs and SFAs, we observed a positive direct effect of total n-3 PUFA levels on GlycA (IVW estimate: 0.16; 0.06, 0.27). There was also a direct negative effect of total n-3 PUFA levels on GlycA (IVW estimate: -0.25; -0.49, 0.02).

However, we observed no independent direct effect of total n-3 on CRP (IVW estimate:0.01; -0.11, 0.14) or IL-6 (IVW estimate: -0.06; -0.21, 0.09) levels after controlling for the effect of n-6 PUFA, triglycerides, LDL-cholesterol, MUFAs and SFAs. Additionally, we observed no independent direct effect of total n-6 on CRP (IVW estimate: 0.03; -0.20, 0.20) or IL-6 after controlling for the effect of n-6 PUFA, triglycerides, LDL-cholesterol, MUFAs and SFAs. Results are presented in Figure 6.

#### 2.4. Results from investigating heterogeneity and pleiotropy

The MR-Egger intercept detected no strong evidence of pleiotropy apart from the association between total n-3 levels and GlycA (Supplementary Table S31). Findings were fairly consistent with results using the MR-PRESSO test for pleiotropy (Supplementary Table S32). However, there was evidence of heterogeneity in all analyses apart from in analyses of total n-3 and LA levels on IL-6 (Supplementary Table S31).

#### 2.5. Results from investigating biases impacting MR using MR-Lap

Although recently published research has suggested that sample overlap does not incur as much bias as previously thought^[19]^, we used MR-lap to investigate whether sample overlap was introducing bias (weak instrument bias or winner’s curse) to our results. We were able to run MR-Lap for the analysis using CRP and GlycA, however the threshold limit set by the package resulted in a loss of all IVs in the IL-6 analysis.

The majority of analyses were unaffected by winner’s curse and weak instrument biases introduced through sample overlap. The correct IVW effect only differed from the observed IVW effect when investigating the effect of total n-6 PUFAs on GlycA levels and on CRP levels. In both cases, the observed effect estimate was in the same direction as the corrected estimate and the confidence intervals did not cross the null but were wider than in the main analysis.

##### 2.5.1. **Potential causal effect of DHA levels on inflammatory markers**

For the effect of DHA on CRP, there were 45 instrumental variables left after pruning and in the observed analysis there was no effect of DHA on CRP (IVW: 0.05; 95% CI:-0.02, 0.12). The corrected MR-Lap estimate suggested that biases were not affecting our results (p=0.508), and also found no effect of DHA on CRP levels (corrected IVW: 0.05; -0.02, 0.12).

For the effect of DHA on GlycA, there were 42 instrumental variables left after pruning and in the observed analysis there was no effect of DHA on GlycA (IVW:-0.04; -0.12, 0.05). The corrected MR-Lap estimate suggested that biases were not affecting the results difference (p=0.06) and also found no effect of DHA on GlycA levels (corrected IVW: -0.03; -0.12, 0.06).

##### 2.5.2. **Potential causal effect of LA levels on inflammatory markers**

For the effect of LA on CRP, there were 52 instrumental variables left after pruning and in the observed analysis there was no effect of LA on CRP (IVW: 0.01; -0.08, 0.10 ). The corrected MR-Lap estimate suggested that biases were not affected our results (p=0.751) and also found no effect of LA on CRP (corrected IVW: 0.01; -0.09, 0.11).

For the effect of LA on GlycA, there were 55 instrumental variables left after pruning and in the observed analysis there was an effect of LA on GlycA (IVW: -0.28; -0.39, -0.18). The corrected MR-Lap estimate suggested that biases were not affected our results (p=0.197) and also found an effect of LA on GlycA levels (corrected IVW: -0.27; -0.39, -0.16).

##### 2.5.3. **Potential causal effect of n-3 levels on inflammatory markers**

For the effect of total n-3 on CRP, there were 45 instrumental variables left after pruning an in the observed analysis there was an effect of n-3 on CRP levels (IVW: 0.08; 0.01, 0.14). The corrected MR-Lap estimate suggested that biases were not affecting our results (p=0.983) and also found an effect of n-3 on CRP (corrected IVW: 0.08; 0.01, 0.14).

For the effect of total n-3 on GlycA, there were 45 instrumental variables left after pruning and in the observed analysis there was an effect of total n-3 on GlycA levels (IVW: -0.20; -0.32, -0.09). The corrected MR-Lap estimate suggested that biases were not affecting our results (p=0.985) and also found an effect of total n-3 on GlycA (-0.20; -0.32, -0.20).

##### 2.5.4. **Potential causal effect of n-6 levels on inflammatory markers**

For the effect of total n-6 on CRP, there were 62 instrumental variables left after pruning and in the observed analysis there was no effect of n-6 on CRP levels (IVW: 0.05; -0.04, 0.14). The corrected MR-Lap estimate suggested that biases were affecting our results (p=2.52x10-6), but also found no effect of total n-6 on CRP (corrected IVW: 0.05; -0.04, 0.15).

For the effect of total n-6 on GlycA there were 61 instrumental variables left after pruning and in the observed analysis there was an effect of n-6 on GlycA levels (IVW: -0.30; -0.40, -0.20). The corrected MR-Lap estimate suggested that biases were affecting our results (p=0.006), but the effect estimate was in the same direction and the corrected estimate also found an effect of total n-6 on GlycA (corrected IVW: -0.29; -0.40, -0.18).

#### 2.6. Results from MR analysis focusing on specific PUFA genes

Estimates using only SNPs identified in the *FADS* gene region indicated that long-chain PUFA concentrations (DHA) increased CRP levels (IVW=0.04; 95% CI=0.01, 0.07), but short chain PUFA concentrations (LA) decreased CRP levels (-0.14; -0.20, -0.07). Estimates when using the SNPs identified to be in the *FADS* gene cluster from the DHA GWAS and the LA GWAS indicated the the gene had no effect on GlycA levels (-0.004; -0.03, 0.03 and 0.05; -0.06, 0.13 respectively). Similarly, estimates when using the SNPs identified to be in the *FADS* gene cluster from the DHA GWAS indicated the gene had no effect on IL-6 levels (Wald ratio = -0.01; -0.05, 0.03).

Estimates when using the SNPs identified to be in the *ELOVL2* gene from the DHA GWAS and LA GWAS indicated the gene had no effect on GlycA levels (I-0.11; -0.40, 0.18 and 0.18; -0.08, 0.44 respectively). This was consistent with their effect on CRP levels (-0.16; -0.35,0.03 and 0.02; -0.14, 0.18 respectively) or on IL-6 levels (0.33; -0.62, 1.29 and wald ratio = -0.78; -3.25, 1.69).

#### 2.7. Results from MR-Clust analysis to explore heterogeneity in MR instruments

Given that evidence of associations between the PUFAs and GlycA were largely consistent across cohort and MR analyses, we used MR-Clust^[20]^ to investigate whether there were distinct clusters of SNPs effects driving the observed heterogeneity. In the MR-Clust analysis investigating the effect of SNPs identified in the LA GWAS on GlycA levels, all clusters showed evidence of a positive effect (Supplementary Figure S6). Similarly, when using SNPs identified in the total n-3 PUFA GWAS and SNPs identified in the total n-6 PUFA GWAS, all clusters were positively associated with GlycA levels (Supplementary Figures S7 and S8). The positive effect on all clusters is consistent with the positive effect of the SNPs in the univariable MR analysis. In contrast, when using genetic instruments identified in the DHA GWAS, there were two clusters of five SNPs that were positively associated with GlycA and one cluster of six SNPs that was negatively associated with GlycA (Supplementary Figure S9). A breakdown of the number of SNPs present in each cluster identified by MR-Clust and the direction of the association is presented in Supplementary Table S2.

#### 2.8. Functional annotations of DHA cluster SNPs using FUMAGWAS

To characterise the positive and negative associated clusters within the DHA and GlycA analyses, we performed gene mapping and gene set and tissue enrichment analyses within the 3 identified MR-Clust clusters using FUMAGWAS ([https://fuma.ctglab.nl](https://fuma.ctglab.nl/)). Although there was some evidence of specific gene set enrichment in each cluster, all clusters showed evidence of enrichement in lipid related pathways (e.g., HDL remodelling [REACTOME], plasma lipoprotein assembly remodelling and clearance [REACTOME]) or were implicated in lipid related GWAS (e.g., cholesterol [GWAS catalog]) (Supplementary Tables S3-S6). Overall, most differentially expressed genes were expressed in the liver for cluster 1, breast for cluster 2, and kidney for cluster 5. However, no differentially expressed genes sets met statistical significance (Bonferroni-corrected P-value threshold = 1.67 × 10^−3^) (Supplementary Figure S9).

#### 2.9. Secondary MR analysis and results

We replicated the MR-analysis for outcomes CRP (<http://www.nealelab.is/uk-biobank/>) and IL-6 ^[23]^ using GWAS that used UKB data. There were 8-40 SNPs available to investigate the effect of PUFAs on CRP and 9-48 SNPs available to investigate the effect of PUFAs on IL-6.

SNP-level F statistics were all >10, indicating that IVW estimates were likely not subject to weak instrument bias. The total variance explained (R^2^) by each SNP in the univariable MR was between 0.03% and 4.03% for DHA, 0.03% and 0.55% for LA, 0.02% and 5.21% for total n-3 PUFAs, and 0.01% and 0.47% for total n-6 PUFAs. Genetic instruments, sample sizes and instrument F-statistics for each GWAS are presented in Supplementary Table S33 . Results are presented post Steiger filtering.

Results were consistent with the main analysis when using CRP as the outcome. We observed no strong evidence of effect of DHA, LA or total n-6 on CRP. We also replicated the effect of total n-3 on CRP, finding that higher levels of total n-3 increased levels of CRP (IVW estimate: 0.24; 95% CI: 0.08, 0.40).

When using IL-6 as the outcome, results were largely consistent with the main analysis, and we found no effect of LA, total n-3 or total n-6 on IL-6 levels. However, we did find that higher levels of DHA decreased levels of IL-6 (IVW estimate: -0.05, 95% CI: -0.09, -0.01). Results are presented in Supplementary Table S34.

The MR-Egger intercept detected no strong evidence of pleiotropy (Supplementary Table S35) and findings were fairly consistent with results using the MR-PRESSO test for pleiotropy (Supplementary Table S36). However, there was evidence of heterogeneity in all analyses apart from in analyses of DHA, total omega-3 and total omega-6 levels on IL-6 levels (Supplementary Table S35).
