## Supplementary material for "The Relationship between Polyunsaturated Fatty Acids and Inflammation: Evidence from cohort and Mendelian randomization analyses": tables_and_figures

| **Supplementary Table S1: Associations between PUFAs and inflammatory biomarkers using imputed ALSPAC data stratified by sex at birth** | | | | | | | | |
| --- | --- | --- | --- | --- | --- | --- | --- | --- |
|  | **Model 1 (Unadjusted)** | | | | **Model 2*** | | | |
|  | **Female (N = 1791)** | | **Male (N =** **1011)** | | **Female (N = 1791)** | | **Male (N =** **1011)** | |
|  | **Mean difference per SD increase in exposure (se)** | **95% CI** | **Mean difference per SD increase in exposure (se)** | **95% CI** | **Mean difference per SD increase in exposure (se)** | **95% CI** | **Mean difference per SD increase in exposure (se)** | **95% CI** |
| **Outcome** |  |  |  |  |  |  |  |  |
| **Exposure: DHA at age 24y** | | | | | | | | |
| IL-6 at 24y | -0.18 (0.02) | -0.23, -0.14 | -0.01 (0.04) | -0.08, 0.06 | -0.17 (0.02) | -0.21, -0.12 | -0.01 (0.04) | -0.08, 0.06 |
| CRP at 24y | 0.11 (0.02) | 0.06, 0.15 | 0.004 (0.04) | -0.07, 0.07 | 0.12 (0.02) | 0.08, 0.17 | 0.01 (0.04) | -0.06, 0.08 |
| GlycA at 24y | 0.27 (0.02) | 0.22, 0.31 | 0.38 (0.04) | 0.31, 0.46 | 0.29 (0.02) | 0.24, 0.33 | 0.38 (0.04) | 0.31, 0.46 |
| **Exposure: LA at age 24y** | | | | | | | | |
| IL-6 at 24y | -0.11 (0.02) | -0.16, -0.06 | -0.02 (0.03) | -0.08, 0.03 | -0.1 (0.02) | -0.15, -0.06 | -0.03 (0.03) | -0.08, 0.03 |
| CRP at 24y | 0.12 (0.02) | 0.07, 0.16 | -0.01 (0.03) | -0.07, 0.05 | 0.12 (0.02) | 0.07, 0.17 | -0.01 (0.03) | -0.07, 0.05 |
| GlycA at 24y | 0.43 (0.02) | 0.39, 0.47 | 0.56 (0.03) | 0.5, 0.61 | 0.43 (0.02) | 0.39, 0.47 | 0.56 (0.03) | 0.50, 0.61 |
| **Exposure: Total n-3 at age 24y** | | | | | | | | |
| IL-6 at 24y | -0.19 (0.02) | -0.24, -0.15 | -0.06 (0.03) | -0.13, -0.002 | -0.18 (0.02) | -0.22, -0.13 | -0.06 (0.03) | -0.12, 0.003 |
| CRP at 24y | 0.02 (0.02) | -0.02, 0.07 | -0.05 (0.03) | -0.11, 0.01 | 0.04 (0.02) | -0.01, 0.08 | -0.05 (0.03) | -0.11, 0.01 |
| GlycA at 24y | 0.25 (0.02) | 0.21, 0.3 | 0.43 (0.03) | 0.37, 0.49 | 0.27 (0.02) | 0.23, 0.31 | 0.43 (0.03) | 0.37, 0.49 |
| **Exposure: Total n-6 at age 24y** | | | | | | | | |
| IL-6 at 24y | -0.09 (0.02) | -0.13, -0.04 | -0.004 (0.03) | -0.07, 0.06 | -0.08 (0.02) | -0.13, -0.04 | -0.01 (0.03) | -0.07, 0.06 |
| CRP at 24y | 0.17 (0.02) | 0.13, 0.22 | 0.03 (0.03) | -0.03, 0.08 | 0.18 (0.02) | 0.13, 0.22 | 0.02 (0.03) | -0.04, 0.08 |
| GlycA at 24y | 0.47 (0.02) | 0.43, 0.51 | 0.6 (0.03) | 0.55, 0.66 | 0.47 (0.02) | 0.43, 0.51 | 0.6 (0.03) | 0.55, 0.66 |
| **Exposure: Total-6:total n-3 at age 24y** | | | | | | | | |
| IL-6 at 24y | 0.22 (0.02) | 0.18, 0.26 | 0.15 (0.03) | 0.08, 0.21 | 0.20 (0.02) | 0.16, 0.25 | 0.13 (0.03) | 0.06, 0.20 |
| CRP at 24y | 0.20 (0.02) | 0.15, 0.24 | 0.16 (0.03) | 0.09, 0.22 | 0.18 (0.02) | 0.14, 0.23 | 0.15 (0.03) | 0.08, 0.21 |
| GlycA at 24y | 0.19 (0.02) | 0.15, 0.23 | 0.15 (0.04) | 0.08, 0.23 | 0.17 (0.02) | 0.13, 0.21 | 0.14 (0.04) | 0.07, 0.21 |
| *SD, Standard deviation; se, Standard error; 95% CI, 95% confidence interval; DHA, Docosahexaenoic acid; LA, Linoleic acid; Total n-3, Total omega-3; Total n-6, Total omega-6; IL-6, Interleukin-6; CRP, C-reactive protein; GlycA, Glycoprotein acetyls*  ** Model 2: Estimates adjusted for household social class at birth, maternal highest education qualification at birth, maternal and paternal smoking status during pregnancy, offspring sex at birth, type of drinker at age 24 years, type of smoker at 24 years, and age in months at 24 year clinic.* | | | | | | | | |

| **Supplementary Table S2: Breakdown of the number of SNPs present in each cluster identified by MR-Clust and the direction of association** | | |
| --- | --- | --- |
| **Exposure GWAS** | **Number of SNPs in Cluster** | **Direction of association** |
| DHA | 5 | Positive |
| DHA | 4 | Positive |
| DHA | 6 | Negative |
| DHA | 24 | Null |
| LA | 4 | Positive |
| LA | 7 | Positive |
| LA | 5 | Positive |
| LA | 4 | Null |
| Total n-3 | 5 | Positive |
| Total n-3 | 15 | Positive |
| Total n-3 | 5 | Null |
| Total n-6 | 5 | Positive |
| Total n-6 | 5 | Positive |
| Total n-6 | 4 | Null |
| Total n-6 | 4 | Junk |
| *SNPS, single nucleiotide polymorphisms; MR, Mendelian randomization; DHA, Docosahexaenoic acid; LA, Linoleic acid; Total n-3, Total omega-3; Total n-6, Total omega-6; IL-6, Interleukin-6; CRP, C-reactive protein; GlycA, Glycoprotein acetyls.* | | |

| **Supplementary Table S3*:* Gene set enrichment of DHA cluster 1 associated genes** | | | | | | |
| --- | --- | --- | --- | --- | --- | --- |
| Category | Gene Set | N genes | N overlap | p-value | Adjusted p-value | Genes |
| Chemical_and_Genetic_ pertubation | ROVERSI_GLIOMA_COPY_NUMBER_UP | 96 | 4 | 2.43E-06 | 8.28E-03 | APOC4:APOC2:CLPTM1:RELB |
| Curated_gene_sets | ROVERSI_GLIOMA_COPY_NUMBER_UP | 96 | 4 | 2.43E-06 | 1.58E-02 | APOC4:APOC2:CLPTM1:RELB |
| GWAScatalog | Bipolar II disorder | 16 | 13 | 2.97E-39 | 1.31E-35 | RFXANK:NR2C2AP:NCAN: HAPLN4:TM6SF2:SUGP1: MAU2:GATAD2A:CILP2:PBX4:LPAR2:GMIP:ATP13A1 |
|  | Bipolar I disorder | 76 | 13 | 7.94E-28 | 1.76E-24 | RFXANK:NR2C2AP:NCAN: HAPLN4:TM6SF2:SUGP1: MAU2:GATAD2A:CILP2:PBX4:LPAR2:GMIP:ATP13A1 |
|  | Bipolar disorder | 471 | 13 | 3.52E-17 | 5.19E-14 | RFXANK:NR2C2AP:NCAN: HAPLN4:TM6SF2:SUGP1: MAU2:GATAD2A:CILP2:PBX4:LPAR2:GMIP:ATP13A1 |
|  | Response to fenofibrate (LDL cholesterol levels) | 4 | 4 | 7.77E-13 | 8.59E-10 | MAU2:GATAD2A:PBX4: ATP13A1 |
|  | LDL cholesterol | 146 | 7 | 7.38E-11 | 6.53E-08 | LIPG:NCAN:SUGP1:CILP2: PBX4:APOC4:APOC2 |
| Positional_ gene_sets | chr19p13 | 547 | 13 | 2.46E-16 | 7.38E-14 | RFXANK:NR2C2AP:NCAN: HAPLN4:TM6SF2:SUGP1: MAU2:GATAD2A:CILP2:PBX4:LPAR2:GMIP:ATP13A1 |
| Reactome | REACTOME_HDL_REMODELING | 10 | 2 | 4.42E-05 | 4.44E-02 | LIPG:APOC2 |
|  | REACTOME_PLASMA_LIPOPROTEIN_ASSEMBLY_REMODELING_AND_CLEARANCE | 73 | 3 | 5.37E-05 | 4.44E-02 | LIPG:APOC4:APOC2 |
| TF_targets | PBXIP1_TARGET_GENES | 197 | 4 | 4.17E-05 | 4.65E-02 | ZMIZ2:LIPG:MAU2:GMIP |
| Note: Results as reported by the Functional Mapping and Annotation (FUMA) of Genome-Wide Association Studies GENE2FUNC function. Top 5 gene sets based on lowest adjusted p values are presented per category. The set of background genes (i.e., the genes against which the set of prioritized genes are tested against) was 20,260 protein-coding genes. “Adjusted pval” refers to a Benjamini–Hochberg false discovery rate corrected p-value per category of tested gene sets. | | | | | | |

| **Supplementary Table S4*:* Gene set enrichment of DHA cluster 2 associated genes (1)** | | | | | | |
| --- | --- | --- | --- | --- | --- | --- |
| Category | Gene Set | N genes | N overlap | p-value | Adjusted p-value | Genes |
| Curated_gene_sets | WP_STATIN_INHIBITION_OF_CHOLESTEROL_PRODUCTION | 29 | 3 | 1.19E-07 | 7.71E-04 | CETP:APOE:APOC1 |
|  | REACTOME_NR1H3_NR1H2_REGULATE_GENE_EXPRESSION_LINKED_TO_CHOLESTEROL_TRANSPORT_AND_EFFLUX | 37 | 3 | 2.52E-07 | 8.18E-04 | CETP:APOE:APOC1 |
|  | REACTOME_NR1H2_AND_NR1H3_MEDIATED_SIGNALING | 47 | 3 | 5.25E-07 | 1.14E-03 | CETP:APOE:APOC1 |
|  | WP_CHOLESTEROL_METABOLISM | 72 | 3 | 1.92E-06 | 2.61E-03 | CETP:APOE:APOC1 |
|  | REACTOME_PLASMA_LIPOPROTEIN_ASSEMBLY_REMODELING_AND_CLEARANCE | 73 | 3 | 2.01E-06 | 2.61E-03 | CETP:APOE:APOC1 |
| Canonical_Pathways | WP_STATIN_INHIBITION_OF_CHOLESTEROL_PRODUCTION | 29 | 3 | 1.19E-07 | 3.67E-04 | CETP:APOE:APOC1 |
|  | REACTOME_NR1H3_NR1H2_REGULATE_GENE_EXPRESSION_LINKED_TO_CHOLESTEROL_TRANSPORT_AND_EFFLUX | 37 | 3 | 2.52E-07 | 3.89E-04 | CETP:APOE:APOC1 |
|  | REACTOME_NR1H2_AND_NR1H3_MEDIATED_SIGNALING | 47 | 3 | 5.25E-07 | 5.41E-04 | CETP:APOE:APOC1 |
|  | WP_CHOLESTEROL_METABOLISM | 72 | 3 | 1.92E-06 | 1.24E-03 | CETP:APOE:APOC1 |
|  | REACTOME_PLASMA_LIPOPROTEIN_ASSEMBLY_REMODELING_AND_CLEARANCE | 73 | 3 | 2.01E-06 | 1.24E-03 | CETP:APOE:APOC1 |
|  | REACTOME_HDL_REMODELING | 10 | 2 | 5.45E-06 | 2.41E-03 | CETP:APOE |
|  | WP_LIPID_PARTICLES_COMPOSITION | 10 | 2 | 5.45E-06 | 2.41E-03 | CETP:APOE |
| Reactome | REACTOME_NR1H3_NR1H2_REGULATE_GENE_EXPRESSION_LINKED_TO_CHOLESTEROL_TRANSPORT_AND_EFFLUX | 37 | 3 | 2.52E-07 | 4.17E-04 | CETP:APOE:APOC1 |
|  | REACTOME_NR1H2_AND_NR1H3_MEDIATED_SIGNALING | 47 | 3 | 5.25E-07 | 4.34E-04 | CETP:APOE:APOC1 |
|  | REACTOME_PLASMA_LIPOPROTEIN_ASSEMBLY_REMODELING_AND_CLEARANCE | 73 | 3 | 2.01E-06 | 1.11E-03 | CETP:APOE:APOC1 |
|  | REACTOME_HDL_REMODELING | 10 | 2 | 5.45E-06 | 2.25E-03 | CETP:APOE |
|  | REACTOME_PLASMA_LIPOPROTEIN_ASSEMBLY | 19 | 2 | 2.07E-05 | 6.84E-03 | APOE:APOC1 |
| GO_mf | GOMF_PHOSPHATIDYLCHOLINE_STEROL_O_ACYLTRANSFERASE_ACTIVATOR_ACTIVITY | 6 | 2 | 1.82E-06 | 3.22E-03 | APOE:APOC1 |
|  | GOMF_STEROL_TRANSFER_ACTIVITY | 23 | 2 | 3.06E-05 | 2.70E-02 | CETP:APOE |
|  | *Supplementary Table S4: Gene set enrichment of DHA cluster 2 associated genes cont. (2)* | | | | | |
|  | GOMF_PHOSPHATIDYLCHOLINE_BINDING | 28 | 2 | 4.57E-05 | 2.70E-02 | CETP:APOC1 |
|  | GOMF_STEROL_TRANSPORTER_ACTIVITY | 36 | 2 | 7.60E-05 | 3.37E-02 | CETP:APOE |
|  | GOMF_QUATERNARY_AMMONIUM_GROUP_BINDING | 50 | 2 | 0.000147 | 4.35E-02 | CETP:APOC1 |

| **Supplementary Table S5: Gene set enrichment of DHA cluster 2 associated genes continued (1)** | | | | | | |
| --- | --- | --- | --- | --- | --- | --- |
| Category | Gene Set | N genes | N overlap | p-value | Adjusted p-value | Genes |
| GO_mf | GOMF_LIPID_TRANSFER_ACTIVITY | 50 | 2 | 0.000147 | 4.35E-02 | CETP:APOE |
| GO_bp | GOBP_HIGH_DENSITY_LIPOPROTEIN_PARTICLE_REMODELING | 15 | 3 | 1.48E-08 | 1.15E-04 | CETP:APOE:APOC1 |
|  | GOBP_PROTEIN_CONTAINING_COMPLEX_REMODELING | 31 | 3 | 1.46E-07 | 5.66E-04 | CETP:APOE:APOC1 |
|  | GOBP_PROTEIN_LIPID_COMPLEX_SUBUNIT_ORGANIZATION | 47 | 3 | 5.25E-07 | 1.36E-03 | CETP:APOE:APOC1 |
|  | GOBP_CHOLESTEROL_EFFLUX | 52 | 3 | 7.15E-07 | 1.39E-03 | CETP:APOE:APOC1 |
|  | GOBP_REGULATION_OF_STEROL_TRANSPORT | 60 | 3 | 1.11E-06 | 1.71E-03 | CETP:APOE:APOC1 |
| Wikipathways | WP_STATIN_INHIBITION_OF_CHOLESTEROL_PRODUCTION | 29 | 3 | 1.19E-07 | 8.70E-05 | CETP:APOE:APOC1 |
|  | WP_CHOLESTEROL_METABOLISM | 72 | 3 | 1.92E-06 | 7.05E-04 | CETP:APOE:APOC1 |
|  | WP_LIPID_PARTICLES_COMPOSITION | 10 | 2 | 5.45E-06 | 1.33E-03 | CETP:APOE |
|  | WP_FAMILIAL_HYPERLIPIDEMIA_TYPE_3 | 13 | 2 | 9.45E-06 | 1.73E-03 | CETP:APOE |
|  | WP_METABOLIC_PATHWAY_OF_LDL_HDL_AND_TG_INCLUDING_DISEASES | 17 | 2 | 1.65E-05 | 2.41E-03 | CETP:APOE |
| Computational_gene_sets | MODULE_235 | 76 | 3 | 2.27E-06 | 1.94E-03 | CETP:APOE:APOC1 |
|  | MODULE_236 | 18 | 2 | 1.85E-05 | 7.94E-03 | CETP:APOE |
| GO_cc | GOCC_HIGH_DENSITY_LIPOPROTEIN_PARTICLE | 25 | 3 | 7.47E-08 | 7.54E-05 | CETP:APOE:APOC1 |
|  | GOCC_PROTEIN_LIPID_COMPLEX | 37 | 3 | 2.52E-07 | 1.27E-04 | CETP:APOE:APOC1 |
|  | GOCC_CHYLOMICRON | 12 | 2 | 7.99E-06 | 2.69E-03 | APOE:APOC1 |
|  | GOCC_TRIGLYCERIDE_RICH_PLASMA_LIPOPROTEIN_PARTICLE | 19 | 2 | 2.07E-05 | 5.22E-03 | APOE:APOC1 |
| GWAScatalog | Metabolic syndrome | 102 | 6 | 1.63E-13 | 7.21E-10 | GALNT2:HERPUD1:CETP:TOMM40:APOE:APOC1 |
|  | Serum metabolite levels (CMS) | 57 | 5 | 4.71E-12 | 1.04E-08 | GALNT2:CETP:TOMM40:APOE:APOC1 |
|  | HDL cholesterol | 196 | 6 | 8.79E-12 | 1.30E-08 | GALNT2:HERPUD1:CETP:TOMM40:APOE:APOC1 |
|  | Cerebral amyloid deposition positivity (PET imaging) | 3 | 3 | 3.26E-11 | 2.41E-08 | TOMM40:APOE:APOC1 |
|  | Cognitive impairment test score | 3 | 3 | 3.26E-11 | 2.41E-08 | TOMM40:APOE:APOC1 |
|  | Cardiovascular risk factors | 3 | 3 | 3.26E-11 | 2.41E-08 | HERPUD1:CETP:APOE |
|  | Lipid traits | 21 | 4 | 4.19E-11 | 2.65E-08 | CETP:TOMM40:APOE:APOC1 |

| *Supplementary Table S5: Gene set enrichment of DHA cluster 2 associated genes cont. (2)* | | | | | | |
| --- | --- | --- | --- | --- | --- | --- |
| Category | Gene Set | N genes | N overlap | p-value | Adjusted p-value | Genes |
| Positional_gene_sets | chr16q13 | 26 | 2 | 3.93E-05 | 1.18E-02 | HERPUD1:CETP |
| Cell_type_signature | HE_LIM_SUN_FETAL_LUNG_C2_APOE_POS_M1_MACROPHAGE_CELL | 14 | 3 | 1.19E-08 | 9.84E-06 | CETP:APOE:APOC1 |
|  | DURANTE_ADULT_OLFACTORY_NEUROEPITHELIUM_MACROPHAGES | 67 | 3 | 1.55E-06 | 6.42E-04 | HERPUD1:APOE:APOC1 |
|  | MANNO_MIDBRAIN_NEUROTYPES_HMGL | 549 | 4 | 2.45E-05 | 6.77E-03 | ZFAND2A:HERPUD1:APOE:APOC1 |
|  | MANNO_MIDBRAIN_NEUROTYPES_HENDO | 838 | 4 | 0.000128 | 2.21E-02 | ZFAND2A:HERPUD1:CETP:TOMM40 |
|  | FAN_OVARY_CL5_HEALTHY_SELECTABLE_FOLLICLE_THECAL_CELL | 296 | 3 | 0.000133 | 2.21E-02 | ZFAND2A:APOE:APOC1 |
|  | FAN_EMBRYONIC_CTX_BIG_GROUPS_MICROGLIA | 345 | 3 | 0.000209 | 2.90E-02 | HERPUD1:APOE:APOC1 |
| Cancer_modules | MODULE_235 | 76 | 3 | 2.27E-06 | 9.77E-04 | CETP:APOE:APOC1 |
|  | MODULE_236 | 18 | 2 | 1.85E-05 | 3.99E-03 | CETP:APOE |
|  | MODULE_17 | 351 | 3 | 0.00022 | 3.16E-02 | TOMM40:APOE:APOC1 |
| Chemical_and_Genetic_pertubation | ROVERSI_GLIOMA_COPY_NUMBER_UP | 96 | 3 | 4.59E-06 | 1.56E-02 | TOMM40:APOE:APOC1 |
|  | PICCALUGA_ANGIOIMMUNOBLASTIC_LYMPHOMA_UP | 202 | 3 | 4.27E-05 | 4.56E-02 | CETP:APOE:APOC1 |
|  | APPEL_IMATINIB_RESPONSE | 31 | 2 | 5.61E-05 | 4.56E-02 | APOE:APOC1 |
|  | ABE_VEGFA_TARGETS_2HR | 31 | 2 | 5.61E-05 | 4.56E-02 | HERPUD1:APOE |
|  | BLANCO_MELO_INFLUENZA_A_INFECTION_A594_CELLS_UP | 36 | 2 | 7.60E-05 | 4.56E-02 | ZFAND2A:HERPUD1 |
|  | ZHAN_MULTIPLE_MYELOMA_DN | 37 | 2 | 8.03E-05 | 4.56E-02 | APOE:APOC1 |
| Note: Results as reported by the Functional Mapping and Annotation (FUMA) of Genome-Wide Association Studies GENE2FUNC function. Top 5 gene sets based on lowest adjusted p values are presented per category. The set of background genes (i.e., the genes against which the set of prioritized genes are tested against) was 20,260 protein-coding genes. “Adjusted pval” refers to a Benjamini–Hochberg false discovery rate corrected p-value per category of tested gene sets. | | | | | | |

| **Supplementary Table S6*:* Gene set enrichment of DHA cluster 5 associated genes** | | | | | | |
| --- | --- | --- | --- | --- | --- | --- |
| Category | Gene Set | N genes | N overlap | p-value | Adjusted p-value | Genes |
| Cancer_gene_neighborhoods | MORF_CDC10 | 133 | 3 | 9.69E-05 | 4.14E-02 | USP1:TP53BP1:SPG11 |
| GWAScatalog | Triglycerides | 149 | 6 | 3.91E-10 | 1.73E-06 | DOCK7:ANGPTL3:TUBGCP4:TP53BP1:MAP1A:FRMD5 |
|  | Regular attendance at a gym or sports club | 30 | 3 | 1.07E-06 | 2.37E-03 | USP1:DOCK7:ANGPTL3 |
|  | Triglyceride levels | 527 | 5 | 1.91E-05 | 2.73E-02 | USP1:DOCK7:ANGPTL3:TP53BP1:FRMD5 |
|  | Lipoprotein phospholipase A2 activity in cardiovascular disease | 11 | 2 | 2.47E-05 | 2.73E-02 | TP53BP1:FRMD5 |
|  | Cholesterol | 98 | 3 | 3.90E-05 | 3.21E-02 | DOCK7:ANGPTL3:ST3GAL4 |
|  | HDL cholesterol levels x alcohol consumption (regular vs non-regular drinkers) interaction (2df) | 102 | 3 | 4.40E-05 | 3.21E-02 | DOCK7:ST3GAL4:TP53BP1 |
|  | HDL cholesterol levels x alcohol consumption (drinkers vs non-drinkers) interaction (2df) | 107 | 3 | 5.07E-05 | 3.21E-02 | DOCK7:ST3GAL4:TP53BP1 |
|  | Apolipoprotein A1 levels | 362 | 4 | 8.77E-05 | 4.62E-02 | DOCK7:ST3GAL4:TP53BP1:SPG11 |
|  | Lipid traits | 21 | 2 | 9.39E-05 | 4.62E-02 | USP1:DOCK7 |
| Positional_gene_sets | chr15q15 | 87 | 6 | 1.47E-11 | 4.40E-09 | TUBGCP4:TP53BP1:MAP1A:PPIP5K1:FRMD5:CASC4 |
|  | chr1p31 | 78 | 3 | 1.97E-05 | 2.95E-03 | USP1:DOCK7:ANGPTL3 |
| Note: Results as reported by the Functional Mapping and Annotation (FUMA) of Genome-Wide Association Studies GENE2FUNC function. Top 5 gene sets based on lowest adjusted p values are presented per category. The set of background genes (i.e., the genes against which the set of prioritized genes are tested against) was 20,260 protein-coding genes. “Adjusted pval” refers to a Benjamini–Hochberg false discovery rate corrected p-value per category of tested gene sets. | | | | | | |

**Supplementary Table S7: Analysis and auxiliary variables included to inform ALSPAC imputation model (1)**

**Main Variables**

DHA (mmol/l) at age 24 years

LA (mmol/l) at age 24 years

Total omega-3 (mmol/l) at age 24 years

Total omega-6 (mmol/l) at age 24 years

CRP (mg/l) at age 24 years

IL-6 (NPX values log2 scale) at age 24 years

GlycA (mmol/l) at 24 years

Household social class at child’s birth

Maternal highest education qualification at child’s birth

Maternal smoking status during pregnancy

Paternal smoking status during pregnancy

Sex at birth

Drinking status at age 24 years

Smoking status at age 24 years

Age in months at 24 year clinic attendence

LDL (mmol/l) at age 24 years

Triglycerides (mmol/l) at age 24 years

Monounsaturated fatty acids (mmol/l) at age 24 years

Saturated fatty acids (mmol/l) at age 24 years

**Auxiliary Variables**

Birthweight from obstetrics data (g)

BMI at age 7 years

BMI at age 9 years

BMI at age 10 years

BMI at age 11 years

BMI at age 12 years

BMI at age 13 years

BMI at age 15 years

BMI at age 17 years

BMI at age 24 years

Conjugated LA (mmol/l) at age 7 years

Conjugated LA (mmol/l) at age 15 years

Conjugated LA (mmol/l) at age 17 years

CRP (mg/l) at age 9 years

CRP (mg/l) at age 15 years

CRP (mg/l) at age 17 years

Daily DHA intake (g) from fish only from FFQ at 3 years

Daily DHA intake (g) from fish only from FFQ at 4 years

Daily DHA intake (g) from fish only from FFQ at 7years Version 2

Daily DHA intake (g) from fish only from FFQ at 9 years

Daily DHA intake from fish only (g) FFQ for mothers at 32 weeks gestation

DHA (mmol/l) at age 7 years

DHA (mmol/l) at age 15 years

DHA (mmol/l) at age 17 years

Energy intake (kcal) DD mean at age 10 years

Estimated degree of unsaturation at age 24 years

GlycA (mmol/l) at age 7 years

GlycA (mmol/l) at age 15 years

*Supplementary Table S7:* Analysis and auxiliary variables included to inform ALSPAC imputation model *cont. (2)*

GlycA (mmol/l) at age 17 years

IL-6 pg/ml at age 9 years

LA (mmol/l) at age 7 years

LA (mmol/l) at age 15 years

LA (mmol/l) at age 17 years

LDL (mmol/l) at age 7 years

LDL (mmol/l) at age 9 years

LDL (mmol/l) at age 15 years

LDL (mmol/l) at age 17 years

Maternal BMI when child was age 18 years

Monounsaturated fatty acids (mmol/l) at age 7 years

Monounsaturated fatty acids (mmol/l) at age 15 years

Monounsaturated fatty acids (mmol/l) at age 17 years

Omega-3 fatty acids (mmol/l) at age 7 years

Omega-6 fatty acids (mmol/l) at age 7 years

Omega-3 fatty acids (mmol/l) at age 15 years

Omega-6 fatty acids (mmol/l) at age 15 years

Omega-3 fatty acids (mmol/l) at age 17 years

Omega-6 fatty acids (mmol/l) at age 17 years

Polyunsaturated fatty acids (mmol/l) at age 24 years

Mother pre-pregnancy height (cm)

Mother pre-pregnancy weight (kg)

Ratio of conjugated LA to total fatty acids (%) at age 7 years

Ratio of DHA to total fatty acids (%) at age 7 years

Ratio of LA to total fatty acids (%) at age 7 years

Ratio of omega-3 fatty acids to total fatty acids (%) at age 7 years

Ratio of omega-6 fatty acids to total fatty acids (%) at age 7 years

Ratio of conjugated LA to total fatty acids (%) at age 15 years

Ratio of DHA to total fatty acids (%) at age 15 years

Ratio of LA to total fatty acids (%) at age 15 years

Ratio of omega-3 fatty acids to total fatty acids (%) at age 15 years

Ratio of omega-6 fatty acids to total fatty acids (%) at age 15 years

Ratio of conjugated LA to total fatty acids (%) at age 17 years

Ratio of DHA to total fatty acids (%) at age 17 years

Ratio of LA to total fatty acids (%) at age 17 years

Ratio of omega-3 fatty acids to total fatty acids (%) at age 17 years

Ratio of omega-6 fatty acids to total fatty acids (%) at age 17 years

Ratio of DHA to total fatty acids (%) at age 24 years

Ratio of LA to total fatty acids (%) at age 24 years

Ratio of omega-3 fatty acids to total fatty acids (%) at age 24 years

Ratio of omega-6 fatty acids to total fatty acids (%) at age 24 years

Ratio of polyunsaturated fatty acids to total fatty acids (%) at age 24 years

Total energy intake (kcal) at age 7 years

Total fatty acids (mmol/l) at age 24 years

Triglycerides (mmol/l) at age 7 years

Triglycerides (mmol/l) at age 9 years

Triglycerides (mmol/l) at age 15 years

Triglycerides (mmol/l) at age 17 years

ALSPAC, Avon longitudinal study of parents and children; DHA, docosahexaenoic acid; LA, linoleic acid; CRP, C-reactive protein; IL-6, Interleukin-6; GlycA, Glycoprotein acetyls; LDL, Low-density lipoprotein; BMI, body mass index; FFQ, Food Frequency Questionnaire

| **Supplementary Table S8: Number of SNPs available for each analysis and those removed due to not fulfilling selection criteria.** | | | | | | | |
| --- | --- | --- | --- | --- | --- | --- | --- |
| **Genome-wide significant SNPs (p<5x10^-8^)** | **Outcome** | **Available SNPs in outcome** | **Palindromic SNPs with intermediate allele frequencies** | **SNPs in LD** | **SNPs that did not pass Steiger filtering** | | **Available SNPs in the analysis** |
| Exposure GWAS: DHA | | | | | | | |
| 48 | CRP | 47 | 6 | 1 | 2 | | 38 |
| 48 | GlycA | 48 | 2 | 0 | 0 | | 46 |
| 48 | IL-6 | 11 | 0 | 0 | 0 | | 11 |
| Exposure GWAS: LA | | | | | | | |
| 57 | CRP | 53 | 9 | 4 | 3 | 37 | |
| 57 | GlycA | 57 | 3 | 4 | 0 | 50 | |
| 57 | IL-6 | 14 | 1 | 0 | 0 | 13 | |
| Exposure GWAS: total n-3 | | | | | | | |
| 52 | CRP | 50 | 7 | 0 | 3 | 40 | |
| 52 | GlycA | 52 | 1 | 2 | 0 | 49 | |
| 52 | IL-6 | 19 | 1 | 1 | 0 | 17 | |
| Exposure GWAS: total n-6 | | | | | | | |
| 63 | CRP | 58 | 10 | 4 | 4 | 40 | |
| 63 | GlycA | 63 | 1 | 4 | 0 | 58 | |
| 63 | IL-6 | 19 | 1 | 0 | 0 | 18 | |
| *SNPS, Single nucleotide polymorphisms; LD, Linkage disequilibrium; GWAS, genome-wide association study; DHA, Docosahexaenoic acid; LA, Linoleic acid; Total n-3, Total omega-3; Total n-6, Total omega-6; IL-6, Interleukin-6; CRP, C-reactive protein; GlycA, Glycoprotein acetyls.* | | | | | | | |

| **Supplementary Table S9: Harmonized SNPs for the association between DHA and GlycA (1)** | | | | | | | | | | |
| --- | --- | --- | --- | --- | --- | --- | --- | --- | --- | --- |
| **SNP** | **Effect Allele** | **Other Allele** | **Beta** | **EAF** | **SE** | **p-value** | **F statistic** | **R^2^ exposure** | **Steiger direction** | **Steiger p-value** |
| rs11122450 | G | T | 0.02 | 0.61 | 0.004 | 3.40E-08 | 27.98 | 2.65E-04 | TRUE | 3.30E-02 |
| rs11230829 | G | A | -0.09 | 0.03 | 0.01 | 9.60E-11 | 54.39 | 3.58E-04 | TRUE | 3.98E-04 |
| rs112875651 | A | G | -0.05 | 0.39 | 0.004 | 1.60E-32 | 132.32 | 1.22E-03 | FALSE | 6.93E-04 |
| rs11681659 | T | C | -0.03 | 0.72 | 0.004 | 5.30E-12 | 40.67 | 3.90E-04 | TRUE | 1.19E-05 |
| rs12226389 | C | T | -0.04 | 0.19 | 0.01 | 3.80E-18 | 66.72 | 6.25E-04 | TRUE | 8.09E-07 |
| rs1260326 | C | T | -0.05 | 0.60 | 0.004 | 8.70E-30 | 120.00 | 1.14E-03 | FALSE | 1.18E-18 |
| rs12914626 | T | C | -0.06 | 0.70 | 0.004 | 7.50E-45 | 178.68 | 1.67E-03 | TRUE | 1.85E-19 |
| rs13424225 | T | G | 0.02 | 0.45 | 0.004 | 1.10E-08 | 28.46 | 2.70E-04 | TRUE | 2.01E-03 |
| rs139974673 | C | T | 0.07 | 0.03 | 0.01 | 8.80E-09 | 29.25 | 2.77E-04 | TRUE | 8.24E-01 |
| rs143355652 | T | C | -0.15 | 0.01 | 0.02 | 6.60E-14 | 54.64 | 4.95E-04 | TRUE | 1.29E-06 |
| rs145659493 | A | C | 0.09 | 0.02 | 0.02 | 1.70E-09 | 32.19 | 3.05E-04 | TRUE | 2.52E-04 |
| rs145786300 | A | G | -0.13 | 0.01 | 0.02 | 4.00E-12 | 47.38 | 4.23E-04 | TRUE | 2.93E-05 |
| rs149820547 | G | T | -0.05 | 0.04 | 0.01 | 4.80E-08 | 27.49 | 2.58E-04 | TRUE | 2.54E-04 |
| rs1560390 | C | T | -0.05 | 0.22 | 0.005 | 2.50E-26 | 99.33 | 9.31E-04 | TRUE | 1.46E-12 |
| rs16940904 | T | C | -0.04 | 0.23 | 0.005 | 2.10E-17 | 63.08 | 5.93E-04 | TRUE | 5.10E-08 |
| rs174564 | G | A | -0.29 | 0.35 | 0.004 | 1.00E-200 | 4547.24 | 4.03E-02 | TRUE | 0.00E+00 |
| rs1800978 | G | C | -0.03 | 0.12 | 0.01 | 4.80E-08 | 28.22 | 2.65E-04 | TRUE | 9.45E-03 |
| rs182611493 | G | A | -0.16 | 0.01 | 0.02 | 1.60E-16 | 70.52 | 5.83E-04 | TRUE | 3.00E-03 |
| rs2232143 | C | T | 0.08 | 0.02 | 0.01 | 6.60E-09 | 35.52 | 3.09E-04 | TRUE | 3.03E-05 |
| rs2278426 | T | C | -0.06 | 0.04 | 0.01 | 3.70E-08 | 28.43 | 2.70E-04 | TRUE | 2.92E-03 |
| rs2394976 | T | G | -0.03 | 0.16 | 0.01 | 5.60E-11 | 36.86 | 3.51E-04 | FALSE | 6.10E-01 |
| rs261291 | C | T | 0.12 | 0.36 | 0.004 | 5.40E-172 | 703.70 | 6.58E-03 | TRUE | 6.94E-83 |
| rs273912 | T | G | 0.03 | 0.71 | 0.004 | 6.60E-10 | 31.76 | 3.02E-04 | TRUE | 4.16E-05 |
| rs2807967 | C | T | 0.03 | 0.72 | 0.004 | 1.20E-08 | 29.37 | 2.76E-04 | TRUE | 3.84E-04 |
| rs325 | C | T | 0.04 | 0.10 | 0.01 | 3.80E-09 | 31.22 | 2.97E-04 | FALSE | 2.51E-06 |
| rs34722314 | A | T | -0.05 | 0.14 | 0.01 | 6.80E-18 | 66.24 | 6.19E-04 | TRUE | 7.85E-09 |
| rs35177659 | T | C | 0.02 | 0.52 | 0.004 | 2.70E-08 | 30.45 | 2.65E-04 | TRUE | 1.71E-01 |
| rs3764261 | A | C | 0.04 | 0.32 | 0.004 | 1.90E-21 | 77.57 | 7.35E-04 | TRUE | 4.00E-05 |
| rs4986970 | T | A | -0.07 | 0.03 | 0.01 | 1.30E-10 | 35.94 | 3.39E-04 | TRUE | 7.20E-04 |
| rs525028 | A | G | -0.03 | 0.71 | 0.004 | 2.70E-12 | 41.48 | 3.92E-04 | FALSE | 2.98E-02 |
| rs55891451 | C | A | 0.03 | 0.20 | 0.005 | 5.60E-11 | 38.65 | 3.64E-04 | TRUE | 1.26E-04 |
| rs58542926 | T | C | -0.12 | 0.07 | 0.01 | 1.40E-57 | 224.22 | 2.12E-03 | TRUE | 2.05E-09 |
| *Supplementary Table S9: Harmonized SNPs for the association between DHA and GlycA cont. (2)* | | | | | | | | | | |
| rs629301 | T | G | 0.04 | 0.78 | 0.005 | 1.40E-19 | 76.11 | 7.23E-04 | TRUE | 3.99E-08 |
| rs638714 | T | G | -0.04 | 0.35 | 0.004 | 5.40E-20 | 74.43 | 7.00E-04 | TRUE | 8.31E-01 |
| rs673335 | C | T | -0.06 | 0.16 | 0.01 | 8.10E-25 | 95.00 | 9.00E-04 | TRUE | 2.34E-12 |
| rs6931604 | T | C | 0.02 | 0.60 | 0.004 | 2.70E-08 | 29.01 | 2.73E-04 | TRUE | 1.40E-03 |
| rs72789541 | A | T | -0.08 | 0.30 | 0.004 | 5.60E-71 | 291.04 | 2.74E-03 | TRUE | 2.09E-26 |
| rs72836561 | T | C | -0.06 | 0.03 | 0.01 | 3.60E-08 | 28.00 | 2.66E-04 | TRUE | 5.95E-01 |
| rs73045691 | A | G | 0.03 | 0.30 | 0.005 | 3.60E-10 | 41.29 | 3.55E-04 | TRUE | 1.09E-02 |
| rs73109460 | A | G | -0.04 | 0.12 | 0.01 | 7.30E-11 | 33.82 | 3.17E-04 | TRUE | 1.31E-02 |
| rs7412 | T | C | -0.08 | 0.08 | 0.01 | 5.40E-26 | 100.67 | 9.45E-04 | TRUE | 3.02E-05 |
| rs77960347 | G | A | 0.14 | 0.01 | 0.02 | 2.50E-17 | 60.50 | 5.75E-04 | TRUE | 1.62E-04 |
| rs78689694 | C | G | 0.04 | 0.13 | 0.01 | 4.40E-10 | 38.32 | 3.62E-04 | TRUE | 5.61E-01 |
| rs7924036 | T | G | 0.04 | 0.50 | 0.004 | 4.80E-21 | 73.12 | 6.94E-04 | TRUE | 1.15E-01 |
| rs9304381 | T | C | 0.05 | 0.82 | 0.01 | 5.80E-23 | 86.92 | 8.22E-04 | TRUE | 3.41E-09 |
| rs9987289 | G | A | 0.06 | 0.91 | 0.01 | 1.30E-17 | 64.54 | 6.11E-04 | FALSE | 2.96E-01 |
| *SNPS, Single nucleotide polymorphisms; DHA, Docosahexaenoic acid; GlycA, Glycoprotein acetyls; EAF, Effect Allele Frequency; SE, Standard error* | | | | | | | | | | |

|  | **Supplementary Table S10: Harmonized SNPs for the association of LA and GlycA (1)** | | | | | | | | | | |
| --- | --- | --- | --- | --- | --- | --- | --- | --- | --- | --- | --- |
|  | **SNP** | **Effect Allele** | **Other Allele** | **Beta** | **EAF** | **SE** | **p-value** | **F statistic** | **R^2^ exposure** | **Steiger direction** | **Steiger p-value** |
|  | rs1002687 | A | G | 0.09 | 0.64 | 1.80E-97 | 0.004 | 398.69 | 3.65E-03 | TRUE | 2.25E-16 |
|  | rs1065853 | T | G | -0.19 | 0.08 | 1.90E-143 | 0.007 | 611.02 | 5.53E-03 | TRUE | 3.82E-48 |
|  | rs1081105 | C | A | 0.12 | 0.03 | 8.20E-23 | 0.012 | 90.03 | 8.25E-04 | TRUE | 5.65E-11 |
|  | rs10838724 | T | G | -0.03 | 0.37 | 1.60E-10 | 0.004 | 36.86 | 3.32E-04 | TRUE | 1.58E-05 |
|  | rs112875651 | A | G | -0.05 | 0.39 | 5.00E-35 | 0.004 | 143.34 | 1.28E-03 | FALSE | 1.49E-03 |
|  | rs115478735 | T | A | 0.04 | 0.18 | 1.20E-17 | 0.005 | 63.28 | 5.82E-04 | TRUE | 1.14E-01 |
|  | rs11789603 | T | C | 0.05 | 0.11 | 1.70E-12 | 0.007 | 46.02 | 4.22E-04 | TRUE | 1.60E-05 |
|  | rs12948283 | C | G | 0.03 | 0.30 | 6.40E-09 | 0.005 | 33.25 | 2.77E-04 | TRUE | 5.91E-02 |
|  | rs13108218 | G | A | -0.03 | 0.62 | 5.90E-16 | 0.004 | 58.25 | 5.29E-04 | FALSE | 4.13E-01 |
|  | rs13217434 | C | G | 0.04 | 0.26 | 4.30E-20 | 0.005 | 82.92 | 7.35E-04 | TRUE | 4.17E-05 |
|  | rs141469619 | G | A | 0.12 | 0.01 | 3.80E-10 | 0.021 | 35.42 | 2.91E-04 | TRUE | 9.58E-01 |
|  | rs142158911 | A | G | -0.09 | 0.12 | 1.20E-48 | 0.006 | 199.22 | 1.81E-03 | TRUE | 5.17E-24 |
|  | rs1461729 | G | A | 0.07 | 0.90 | 5.40E-26 | 0.007 | 103.31 | 9.48E-04 | TRUE | 7.38E-01 |
|  | rs174564 | G | A | 0.08 | 0.35 | 5.10E-88 | 0.004 | 371.34 | 3.42E-03 | TRUE | 5.09E-43 |
|  | rs186696265 | T | C | -0.23 | 0.01 | 2.20E-41 | 0.017 | 168.00 | 1.53E-03 | TRUE | 8.58E-04 |
|  | rs1883711 | C | G | 0.09 | 0.03 | 3.70E-15 | 0.012 | 56.04 | 4.94E-04 | TRUE | 2.69E-02 |
|  | rs2378390 | A | G | -0.03 | 0.14 | 7.00E-09 | 0.006 | 29.39 | 2.69E-04 | TRUE | 1.05E-02 |
|  | rs247617 | A | C | 0.05 | 0.32 | 6.50E-33 | 0.004 | 129.42 | 1.19E-03 | TRUE | 4.68E-09 |
|  | rs261290 | C | T | -0.09 | 0.65 | 7.90E-99 | 0.004 | 414.45 | 3.77E-03 | TRUE | 1.11E-48 |
|  | rs2740488 | C | A | -0.05 | 0.27 | 1.00E-25 | 0.005 | 104.62 | 9.58E-04 | TRUE | 5.51E-10 |
|  | rs2986164 | A | G | -0.03 | 0.54 | 5.00E-10 | 0.004 | 40.71 | 3.15E-04 | TRUE | 1.17E-03 |
|  | rs3011437 | G | T | 0.04 | 0.29 | 1.30E-16 | 0.004 | 61.24 | 5.56E-04 | TRUE | 1.10E-03 |
|  | rs34232196 | T | C | -0.03 | 0.25 | 8.30E-11 | 0.005 | 39.85 | 3.62E-04 | TRUE | 4.86E-05 |
|  | rs35599691 | A | G | 0.04 | 0.58 | 6.60E-12 | 0.005 | 68.87 | 4.72E-04 | FALSE | 4.47E-02 |
|  | rs35633876 | T | G | -0.03 | 0.48 | 2.40E-17 | 0.004 | 68.23 | 6.23E-04 | TRUE | 2.03E-05 |
|  | rs36018387 | T | C | -0.04 | 0.11 | 6.40E-12 | 0.007 | 42.41 | 3.88E-04 | TRUE | 1.95E-02 |
|  | rs4008004 | A | C | 0.03 | 0.22 | 1.90E-10 | 0.005 | 37.93 | 3.46E-04 | TRUE | 1.84E-02 |
|  | rs4299376 | T | G | -0.04 | 0.68 | 2.50E-19 | 0.004 | 75.61 | 6.93E-04 | TRUE | 9.71E-09 |
|  | rs4665972 | C | T | -0.05 | 0.60 | 1.30E-35 | 0.004 | 145.10 | 1.32E-03 | FALSE | 7.80E-13 |
| *Supplementary Table S10: Harmonized SNPs for the association of LA and GlycA cont. (2)* | | | | | | | | | | | |
|  | rs4704210 | C | G | 0.05 | 0.37 | 2.00E-35 | 0.004 | 140.95 | 1.30E-03 | TRUE | 1.32E-13 |
|  | rs4947302 | T | C | 0.05 | 0.06 | 4.80E-10 | 0.008 | 29.45 | 2.70E-04 | TRUE | 6.74E-01 |
|  | rs534417 | G | A | 0.04 | 0.88 | 2.50E-09 | 0.006 | 32.61 | 3.02E-04 | TRUE | 8.66E-01 |
|  | rs55747707 | A | G | -0.04 | 0.20 | 1.50E-18 | 0.005 | 75.54 | 6.91E-04 | FALSE | 3.30E-08 |
|  | rs56322906 | A | G | -0.09 | 0.04 | 2.90E-15 | 0.011 | 60.86 | 5.59E-04 | TRUE | 3.70E-06 |
|  | rs58542926 | T | C | -0.11 | 0.07 | 7.50E-48 | 0.008 | 191.30 | 1.76E-03 | TRUE | 5.40E-07 |
|  | rs602633 | G | T | 0.05 | 0.78 | 7.50E-28 | 0.005 | 110.65 | 1.02E-03 | TRUE | 1.00E-11 |
|  | rs633695 | G | A | 0.07 | 0.29 | 2.00E-54 | 0.004 | 231.62 | 2.11E-03 | TRUE | 2.25E-26 |
|  | rs6471717 | A | G | -0.03 | 0.66 | 1.40E-13 | 0.004 | 48.94 | 4.48E-04 | TRUE | 4.04E-03 |
|  | rs6602911 | T | C | 0.02 | 0.36 | 1.40E-08 | 0.004 | 30.04 | 2.77E-04 | TRUE | 2.58E-01 |
|  | rs6882345 | A | G | 0.04 | 0.63 | 2.00E-26 | 0.004 | 104.15 | 9.60E-04 | TRUE | 2.61E-08 |
|  | rs693 | A | G | 0.06 | 0.52 | 3.40E-54 | 0.004 | 217.11 | 1.99E-03 | TRUE | 5.07E-19 |
|  | rs7139079 | A | G | -0.03 | 0.59 | 4.80E-10 | 0.004 | 37.42 | 3.43E-04 | TRUE | 8.93E-02 |
|  | rs740516 | G | C | -0.03 | 0.15 | 1.90E-09 | 0.006 | 33.40 | 3.05E-04 | TRUE | 1.12E-01 |
|  | rs7750288 | G | A | 0.03 | 0.29 | 1.10E-09 | 0.004 | 33.17 | 3.06E-04 | TRUE | 5.04E-03 |
|  | rs77960347 | G | A | 0.25 | 0.01 | 1.10E-45 | 0.018 | 186.00 | 1.71E-03 | TRUE | 1.83E-15 |
|  | rs7816447 | C | T | -0.05 | 0.10 | 8.20E-13 | 0.007 | 49.50 | 4.57E-04 | FALSE | 1.38E-03 |
|  | rs79429216 | A | G | 0.15 | 0.01 | 1.60E-16 | 0.018 | 62.26 | 5.76E-04 | TRUE | 1.05E-03 |
|  | rs9302635 | C | T | -0.03 | 0.18 | 3.40E-08 | 0.005 | 27.58 | 2.55E-04 | FALSE | 1.87E-44 |
|  | rs9304381 | T | C | 0.06 | 0.82 | 1.00E-33 | 0.005 | 135.15 | 1.24E-03 | TRUE | 7.31E-14 |
|  | rs964184 | C | G | -0.15 | 0.87 | 2.00E-136 | 0.006 | 564.21 | 5.15E-03 | TRUE | 7.55E-07 |
|  | *SNPS, Single nucleotide polymorphisms; LA, Linolic acid; GlycA, Glycoprotein acetyls; EAF, Effect Allele Frequency; SE, Standard error* | | | | | | | | | | |

| **Supplementary Table S11: Harmonized SNPs for the association of total n-3 and GlycA (1)** | | | | | | | | | | |
| --- | --- | --- | --- | --- | --- | --- | --- | --- | --- | --- |
| **SNP** | **Effect Allele** | **Other Allele** | **Beta** | **EAF** | **SE** | **p-value** | **F statistic** | **R^2^ exposure** | **Steiger direction** | **Steiger p-value** |
| rs10184054 | G | C | -0.04 | 0.22 | 5.60E-15 | 0.005 | 52.28 | 4.80E-04 | TRUE | 9.35E-02 |
| rs10455872 | G | A | -0.06 | 0.08 | 2.80E-17 | 0.008 | 66.36 | 6.07E-04 | FALSE | 1.60E-01 |
| rs11230829 | G | A | -0.10 | 0.03 | 3.40E-12 | 0.015 | 65.90 | 4.19E-04 | TRUE | 9.06E-05 |
| rs11242109 | T | G | 0.02 | 0.48 | 2.40E-09 | 0.004 | 33.24 | 3.04E-04 | TRUE | 3.35E-04 |
| rs112875651 | A | G | -0.09 | 0.39 | 3.50E-98 | 0.004 | 420.12 | 3.71E-03 | TRUE | 4.15E-03 |
| rs1132899 | C | T | 0.03 | 0.51 | 8.60E-11 | 0.004 | 42.18 | 3.79E-04 | TRUE | 2.04E-02 |
| rs11563251 | T | C | 0.03 | 0.11 | 3.20E-08 | 0.006 | 27.68 | 2.54E-04 | TRUE | 1.53E-03 |
| rs1167998 | A | C | 0.07 | 0.64 | 3.60E-66 | 0.004 | 268.89 | 2.44E-03 | TRUE | 2.19E-08 |
| rs11681659 | T | C | -0.03 | 0.72 | 2.00E-08 | 0.004 | 29.50 | 2.73E-04 | TRUE | 3.11E-04 |
| rs117143374 | C | T | -0.04 | 0.14 | 2.20E-10 | 0.006 | 38.63 | 3.50E-04 | TRUE | 2.66E-02 |
| rs117733303 | G | A | -0.12 | 0.02 | 1.40E-15 | 0.015 | 56.21 | 5.14E-04 | FALSE | 6.18E-01 |
| rs12226389 | C | T | -0.05 | 0.19 | 1.10E-22 | 0.005 | 89.19 | 8.08E-04 | TRUE | 8.79E-09 |
| rs1260326 | C | T | -0.08 | 0.60 | 8.40E-88 | 0.004 | 371.62 | 3.38E-03 | FALSE | 3.24E-03 |
| rs13424225 | T | G | 0.02 | 0.45 | 2.20E-08 | 0.004 | 27.87 | 2.55E-04 | TRUE | 2.89E-03 |
| rs139974673 | C | T | 0.12 | 0.03 | 2.30E-21 | 0.013 | 80.89 | 7.37E-04 | TRUE | 6.04E-03 |
| rs143355652 | T | C | -0.15 | 0.01 | 9.40E-14 | 0.020 | 56.62 | 4.96E-04 | TRUE | 1.25E-06 |
| rs144018203 | C | G | 0.11 | 0.01 | 4.20E-08 | 0.020 | 27.83 | 2.38E-04 | FALSE | 7.08E-01 |
| rs157592 | C | A | 0.03 | 0.19 | 3.60E-09 | 0.005 | 28.07 | 2.44E-04 | TRUE | 1.64E-02 |
| rs1672811 | C | T | 0.03 | 0.75 | 3.00E-08 | 0.005 | 27.47 | 2.50E-04 | TRUE | 2.36E-04 |
| rs16940904 | T | C | -0.04 | 0.23 | 3.90E-14 | 0.005 | 50.90 | 4.61E-04 | TRUE | 1.95E-06 |
| rs174564 | G | A | -0.34 | 0.35 | 1.00E-200 | 0.004 | 6243.53 | 5.21E-02 | TRUE | 0.00E+00 |
| rs1800978 | G | C | -0.04 | 0.12 | 5.20E-09 | 0.006 | 34.84 | 3.16E-04 | TRUE | 3.17E-03 |
| rs182611493 | G | A | -0.21 | 0.01 | 1.10E-27 | 0.020 | 125.01 | 9.95E-04 | TRUE | 2.07E-06 |
| rs2394976 | T | G | -0.05 | 0.16 | 1.20E-15 | 0.006 | 66.40 | 6.10E-04 | TRUE | 3.57E-01 |
| rs261290 | C | T | -0.11 | 0.65 | 3.90E-161 | 0.004 | 684.36 | 6.17E-03 | TRUE | 1.06E-78 |
| rs3018731 | G | A | -0.04 | 0.72 | 2.00E-14 | 0.005 | 58.23 | 5.21E-04 | TRUE | 3.24E-04 |
| rs34663616 | A | C | 0.04 | 0.14 | 4.40E-10 | 0.006 | 34.75 | 3.05E-04 | TRUE | 3.57E-02 |
| rs35135293 | T | C | -0.02 | 0.52 | 3.90E-08 | 0.004 | 25.06 | 2.27E-04 | TRUE | 1.49E-01 |
| rs4000713 | A | G | -0.03 | 0.30 | 1.00E-11 | 0.004 | 39.77 | 3.63E-04 | TRUE | 1.24E-01 |
| rs55891451 | C | A | 0.03 | 0.20 | 4.60E-12 | 0.005 | 43.30 | 3.94E-04 | TRUE | 5.98E-05 |
| *Supplementary Table S11: Harmonized SNPs for the association of total n-3 and GlycA cont. (2)* | | | | | | | | | | |
| rs58542926 | T | C | -0.17 | 0.07 | 1.40E-113 | 0.008 | 468.55 | 4.25E-03 | TRUE | 2.84E-26 |
| rs6129624 | A | G | -0.03 | 0.34 | 5.10E-10 | 0.004 | 34.02 | 3.01E-04 | TRUE | 2.51E-02 |
| rs62466318 | T | C | -0.07 | 0.20 | 1.20E-45 | 0.005 | 194.78 | 1.76E-03 | FALSE | 7.65E-02 |
| rs629301 | T | G | 0.04 | 0.78 | 1.30E-14 | 0.005 | 58.26 | 5.34E-04 | TRUE | 4.56E-06 |
| rs633695 | G | A | 0.08 | 0.29 | 9.10E-80 | 0.004 | 336.77 | 3.04E-03 | TRUE | 8.90E-38 |
| rs6601924 | C | T | 0.04 | 0.85 | 8.50E-10 | 0.006 | 36.89 | 3.36E-04 | TRUE | 1.02E-02 |
| rs6693447 | G | T | 0.02 | 0.46 | 4.80E-09 | 0.004 | 30.11 | 2.75E-04 | TRUE | 6.95E-05 |
| rs673335 | C | T | -0.07 | 0.16 | 1.10E-34 | 0.006 | 138.76 | 1.27E-03 | TRUE | 6.03E-17 |
| rs6882345 | A | G | 0.03 | 0.63 | 1.90E-13 | 0.004 | 44.60 | 4.09E-04 | TRUE | 2.86E-03 |
| rs72789541 | A | T | -0.08 | 0.30 | 5.60E-75 | 0.004 | 316.07 | 2.87E-03 | TRUE | 8.85E-28 |
| rs73109460 | A | G | -0.03 | 0.12 | 9.20E-10 | 0.006 | 30.44 | 2.75E-04 | TRUE | 2.86E-02 |
| rs737338 | T | C | -0.07 | 0.04 | 3.50E-11 | 0.011 | 41.24 | 3.77E-04 | TRUE | 2.90E-04 |
| rs77960347 | G | A | 0.16 | 0.01 | 7.20E-22 | 0.018 | 78.66 | 7.21E-04 | TRUE | 8.24E-06 |
| rs7819706 | G | A | -0.04 | 0.12 | 1.80E-10 | 0.006 | 37.71 | 3.45E-04 | FALSE | 1.74E-04 |
| rs7924036 | T | G | 0.02 | 0.50 | 5.50E-10 | 0.004 | 31.36 | 2.87E-04 | FALSE | 4.98E-01 |
| rs7970695 | A | G | -0.03 | 0.62 | 1.20E-10 | 0.004 | 34.69 | 3.16E-04 | TRUE | 1.22E-01 |
| rs9304381 | T | C | 0.05 | 0.82 | 5.20E-24 | 0.005 | 95.67 | 8.72E-04 | TRUE | 9.54E-10 |
| rs964184 | C | G | -0.12 | 0.87 | 8.90E-87 | 0.006 | 361.40 | 3.31E-03 | TRUE | 1.26E-01 |
| rs9987289 | G | A | 0.06 | 0.91 | 3.20E-16 | 0.007 | 61.10 | 5.59E-04 | FALSE | 1.91E-01 |
| *SNPS, Single nucleotide polymorphisms; n-3, Total omega-3; GlycA, Glycoprotein acetyls; EAF, Effect Allele Frequency; SE, Standard error* | | | | | | | | | | |

| **Supplementary Table S12: Harmonized SNPS for the association of total n-6 and GlycA (1)** | | | | | | | | | | |
| --- | --- | --- | --- | --- | --- | --- | --- | --- | --- | --- |
| **SNP** | **Effect Allele** | **Other Allele** | **Beta** | **EAF** | **SE** | **p-value** | **F statistic** | **R^2^ exposure** | **Steiger direction** | **Steiger p-value** |
| rs1002687 | A | G | 0.09 | 0.64 | 0.004 | 1.00E-107 | 437.62 | 4.04E-03 | TRUE | 3.18E-19 |
| rs1065853 | T | G | -0.20 | 0.08 | 0.01 | 3.60E-160 | 676.17 | 6.16E-03 | TRUE | 9.72E-55 |
| rs1081105 | C | A | 0.12 | 0.03 | 0.01 | 1.80E-22 | 87.22 | 8.06E-04 | TRUE | 9.53E-11 |
| rs112875651 | A | G | -0.06 | 0.39 | 0.004 | 2.20E-53 | 222.88 | 2.01E-03 | FALSE | 3.10E-01 |
| rs114863007 | A | G | -0.05 | 0.09 | 0.01 | 7.30E-12 | 41.19 | 3.78E-04 | TRUE | 3.91E-04 |
| rs115478735 | T | A | 0.04 | 0.18 | 0.01 | 2.20E-17 | 61.58 | 5.71E-04 | TRUE | 1.27E-01 |
| rs11789603 | T | C | 0.05 | 0.11 | 0.01 | 9.70E-14 | 51.16 | 4.73E-04 | TRUE | 4.13E-06 |
| rs1260326 | C | T | -0.06 | 0.60 | 0.004 | 3.90E-55 | 227.58 | 2.11E-03 | FALSE | 3.91E-09 |
| rs12740374 | T | G | -0.06 | 0.22 | 0.005 | 1.50E-32 | 129.94 | 1.21E-03 | TRUE | 5.54E-14 |
| rs13108218 | G | A | -0.04 | 0.62 | 0.004 | 3.60E-18 | 67.35 | 6.16E-04 | FALSE | 7.05E-01 |
| rs141469619 | G | A | 0.11 | 0.01 | 0.02 | 1.40E-08 | 28.22 | 2.34E-04 | FALSE | 7.10E-01 |
| rs142158911 | A | G | -0.09 | 0.12 | 0.01 | 5.20E-52 | 211.68 | 1.94E-03 | TRUE | 1.25E-25 |
| rs1461729 | G | A | 0.08 | 0.90 | 0.01 | 2.80E-36 | 145.67 | 1.35E-03 | TRUE | 7.92E-02 |
| rs1800961 | T | C | -0.07 | 0.03 | 0.01 | 3.30E-10 | 37.35 | 3.47E-04 | TRUE | 4.19E-03 |
| rs183130 | T | C | 0.06 | 0.32 | 0.004 | 1.40E-48 | 190.95 | 1.77E-03 | TRUE | 1.28E-14 |
| rs1883711 | C | G | 0.09 | 0.03 | 0.01 | 3.20E-16 | 59.18 | 5.26E-04 | TRUE | 1.72E-02 |
| rs200730299 | C | A | -0.04 | 0.19 | 0.01 | 7.10E-12 | 48.10 | 3.95E-04 | TRUE | 7.02E-06 |
| rs2378390 | A | G | -0.03 | 0.14 | 0.01 | 3.20E-09 | 30.93 | 2.85E-04 | TRUE | 7.44E-03 |
| rs261290 | C | T | -0.10 | 0.65 | 0.004 | 1.00E-116 | 486.78 | 4.47E-03 | TRUE | 2.41E-57 |
| rs2737245 | T | G | -0.03 | 0.28 | 0.005 | 1.40E-09 | 34.91 | 3.23E-04 | FALSE | 6.90E-01 |
| rs2740488 | C | A | -0.05 | 0.27 | 0.005 | 5.40E-28 | 112.79 | 1.04E-03 | TRUE | 7.00E-11 |
| rs28383314 | C | T | 0.04 | 0.62 | 0.004 | 1.70E-18 | 81.62 | 7.58E-04 | FALSE | 2.98E-01 |
| rs2986164 | A | G | -0.03 | 0.54 | 0.004 | 3.40E-09 | 35.78 | 2.80E-04 | TRUE | 2.71E-03 |
| rs35603463 | C | T | 0.03 | 0.57 | 0.005 | 4.60E-10 | 63.93 | 4.20E-04 | TRUE | 1.71E-01 |
| rs3734854 | A | G | 0.05 | 0.06 | 0.01 | 5.90E-11 | 31.68 | 2.93E-04 | TRUE | 5.53E-01 |
| rs3770586 | T | C | -0.02 | 0.48 | 0.004 | 7.10E-09 | 30.31 | 2.79E-04 | TRUE | 3.94E-03 |
| rs3817335 | A | T | -0.03 | 0.35 | 0.004 | 9.80E-12 | 40.37 | 3.77E-04 | TRUE | 5.42E-06 |
| rs4008004 | A | C | 0.03 | 0.22 | 0.005 | 8.40E-12 | 43.15 | 3.97E-04 | TRUE | 7.50E-03 |
| rs4299376 | T | G | -0.04 | 0.68 | 0.004 | 1.10E-16 | 63.10 | 5.83E-04 | TRUE | 1.84E-07 |
| rs4439799 | T | C | 0.02 | 0.50 | 0.004 | 1.30E-08 | 28.92 | 2.67E-04 | TRUE | 2.75E-01 |
| rs4704210 | C | G | 0.05 | 0.37 | 0.004 | 6.10E-30 | 117.73 | 1.09E-03 | TRUE | 2.15E-11 |
| *Supplementary Table S12: Harmonized SNPS for the association of total n-6 and GlycA cont. (2)* | | | | | | | | | | |
| rs4860948 | A | T | 0.03 | 0.24 | 0.005 | 1.90E-09 | 33.08 | 3.06E-04 | TRUE | 8.16E-05 |
| rs534417 | G | A | 0.04 | 0.88 | 0.01 | 9.30E-11 | 38.35 | 3.59E-04 | TRUE | 5.89E-01 |
| rs55747707 | A | G | -0.05 | 0.20 | 0.01 | 1.70E-22 | 91.41 | 8.43E-04 | FALSE | 1.15E-06 |
| rs56322906 | A | G | -0.10 | 0.04 | 0.01 | 1.20E-19 | 78.65 | 7.29E-04 | TRUE | 5.62E-08 |
| rs5754102 | A | C | -0.03 | 0.18 | 0.01 | 9.90E-10 | 34.38 | 3.12E-04 | TRUE | 3.99E-04 |
| rs58542926 | T | C | -0.13 | 0.07 | 0.01 | 2.50E-65 | 260.26 | 2.41E-03 | TRUE | 1.69E-11 |
| rs633695 | G | A | 0.07 | 0.29 | 0.004 | 1.30E-59 | 250.77 | 2.30E-03 | TRUE | 9.95E-29 |
| rs6471717 | A | G | -0.03 | 0.66 | 0.004 | 4.00E-12 | 43.31 | 4.00E-04 | TRUE | 9.45E-03 |
| rs6547409 | T | C | -0.08 | 0.05 | 0.01 | 2.40E-20 | 74.07 | 6.72E-04 | TRUE | 9.97E-07 |
| rs6602911 | T | C | 0.03 | 0.36 | 0.004 | 1.30E-09 | 35.15 | 3.27E-04 | TRUE | 1.40E-01 |
| rs672889 | G | T | 0.08 | 0.86 | 0.01 | 1.30E-41 | 161.75 | 1.50E-03 | TRUE | 4.30E-20 |
| rs6882345 | A | G | 0.04 | 0.63 | 0.004 | 1.20E-27 | 107.73 | 1.00E-03 | TRUE | 1.03E-08 |
| rs6934962 | T | C | 0.02 | 0.40 | 0.004 | 2.20E-08 | 28.78 | 2.68E-04 | TRUE | 4.27E-01 |
| rs6938647 | C | A | -0.05 | 0.78 | 0.005 | 1.90E-23 | 90.93 | 8.23E-04 | TRUE | 3.19E-04 |
| rs7139079 | A | G | -0.03 | 0.59 | 0.004 | 3.30E-13 | 49.13 | 4.54E-04 | TRUE | 1.79E-02 |
| rs72997616 | A | C | -0.05 | 0.09 | 0.01 | 1.60E-13 | 52.13 | 4.79E-04 | TRUE | 3.48E-06 |
| rs740516 | G | C | -0.03 | 0.15 | 0.01 | 1.40E-08 | 29.42 | 2.71E-04 | TRUE | 1.77E-01 |
| rs75406471 | A | G | -0.03 | 0.15 | 0.01 | 2.70E-08 | 29.67 | 2.74E-04 | TRUE | 2.57E-02 |
| rs7750288 | G | A | 0.02 | 0.29 | 0.004 | 1.30E-08 | 29.12 | 2.71E-04 | TRUE | 1.06E-02 |
| rs77960347 | G | A | 0.28 | 0.01 | 0.02 | 2.80E-56 | 228.54 | 2.12E-03 | TRUE | 1.14E-19 |
| rs7831074 | G | C | 0.03 | 0.76 | 0.01 | 4.60E-08 | 32.61 | 2.64E-04 | TRUE | 7.79E-02 |
| rs79429216 | A | G | 0.15 | 0.01 | 0.02 | 1.30E-17 | 65.71 | 6.13E-04 | TRUE | 5.39E-04 |
| rs870526 | T | C | -0.03 | 0.52 | 0.004 | 7.70E-16 | 58.40 | 5.42E-04 | TRUE | 4.59E-03 |
| rs9295128 | T | G | -0.20 | 0.02 | 0.02 | 3.40E-36 | 146.97 | 1.32E-03 | TRUE | 1.21E-02 |
| rs9304381 | T | C | 0.07 | 0.82 | 0.01 | 7.20E-42 | 168.83 | 1.56E-03 | TRUE | 1.66E-17 |
| rs9616847 | T | A | 0.02 | 0.39 | 0.004 | 1.40E-08 | 31.01 | 2.85E-04 | TRUE | 5.02E-02 |
| rs964184 | C | G | -0.14 | 0.87 | 0.01 | 1.10E-125 | 513.18 | 4.73E-03 | TRUE | 2.45E-05 |
| *SNPS, Single nucleotide polymorphisms; n-6, Total omega-6; GlycA, Glycoprotein acetyls; EAF, Effect Allele Frequency; SE, Standard error* | | | | | | | | | | |

| **Supplementary Table S13:Harmonized SNPs for the association between DHA and CRP (1)** | | | | | | | | | | |
| --- | --- | --- | --- | --- | --- | --- | --- | --- | --- | --- |
| **SNP** | **Effect Allele** | **Other Allele** | **Beta** | **EAF** | **SE** | **p-value** | **F statistic** | **R^2^ exposure** | **Steiger direction** | **Steiger p-value** |
| rs11122450 | G | T | 0.02 | 0.61 | 0.004 | 3.40E-08 | 27.98 | 2.65E-04 | TRUE | 2.18E-04 |
| rs11230829 | G | A | -0.09 | 0.03 | 0.01 | 9.60E-11 | 54.39 | 3.58E-04 | TRUE | 1.86E-08 |
| rs112875651 | A | G | -0.05 | 0.39 | 0.004 | 1.60E-32 | 132.32 | 1.22E-03 | TRUE | 5.81E-12 |
| rs11681659 | T | C | -0.03 | 0.72 | 0.004 | 5.30E-12 | 40.67 | 3.90E-04 | TRUE | 1.02E-06 |
| rs12226389 | C | T | -0.04 | 0.19 | 0.01 | 3.80E-18 | 66.72 | 6.25E-04 | TRUE | 1.11E-12 |
| rs1260326 | C | T | -0.05 | 0.60 | 0.004 | 8.70E-30 | 120.00 | 1.14E-03 | FALSE | 5.31E-07 |
| rs12914626 | T | C | -0.06 | 0.70 | 0.004 | 7.50E-45 | 178.68 | 1.67E-03 | TRUE | 1.24E-32 |
| rs13424225 | T | G | 0.02 | 0.45 | 0.004 | 1.10E-08 | 28.46 | 2.70E-04 | TRUE | 6.78E-05 |
| rs139974673 | C | T | 0.07 | 0.03 | 0.01 | 8.80E-09 | 29.25 | 2.77E-04 | TRUE | 4.55E-01 |
| rs143355652 | T | C | -0.15 | 0.01 | 0.02 | 6.60E-14 | 54.64 | 4.95E-04 | TRUE | 1.29E-11 |
| rs145659493 | A | C | 0.09 | 0.02 | 0.02 | 1.70E-09 | 32.19 | 3.05E-04 | TRUE | 5.11E-07 |
| rs145786300 | A | G | -0.13 | 0.01 | 0.02 | 4.00E-12 | 47.38 | 4.23E-04 | TRUE | 2.95E-10 |
| rs149820547 | G | T | -0.05 | 0.04 | 0.01 | 4.80E-08 | 27.49 | 2.58E-04 | TRUE | 5.08E-06 |
| rs1560390 | C | T | -0.05 | 0.22 | 0.005 | 2.50E-26 | 99.33 | 9.31E-04 | TRUE | 4.07E-17 |
| rs16940904 | T | C | -0.04 | 0.23 | 0.005 | 2.10E-17 | 63.08 | 5.93E-04 | TRUE | 5.10E-12 |
| rs174564 | G | A | -0.29 | 0.35 | 0.004 | 1.00E-200 | 4547.24 | 4.03E-02 | TRUE | 0.00E+00 |
| rs182611493 | G | A | -0.16 | 0.01 | 0.02 | 1.60E-16 | 70.52 | 5.83E-04 | TRUE | 8.42E-10 |
| rs2232143 | C | T | 0.08 | 0.02 | 0.01 | 6.60E-09 | 35.52 | 3.09E-04 | TRUE | 3.34E-06 |
| rs2278426 | T | C | -0.06 | 0.04 | 0.01 | 3.70E-08 | 28.43 | 2.70E-04 | TRUE | 9.60E-06 |
| rs261291 | C | T | 0.12 | 0.36 | 0.004 | 5.40E-172 | 703.70 | 6.58E-03 | TRUE | 7.89E-112 |
| rs273912 | T | G | 0.03 | 0.71 | 0.004 | 6.60E-10 | 31.76 | 3.02E-04 | TRUE | 5.68E-05 |
| rs2807967 | C | T | 0.03 | 0.72 | 0.004 | 1.20E-08 | 29.37 | 2.76E-04 | TRUE | 2.42E-03 |
| rs325 | C | T | 0.04 | 0.10 | 0.01 | 3.80E-09 | 31.22 | 2.97E-04 | TRUE | 6.06E-04 |
| rs35177659 | T | C | 0.02 | 0.52 | 0.004 | 2.70E-08 | 30.45 | 2.65E-04 | TRUE | 1.60E-04 |
| rs3764261 | A | C | 0.04 | 0.32 | 0.004 | 1.90E-21 | 77.57 | 7.35E-04 | TRUE | 8.43E-13 |
| rs525028 | A | G | -0.03 | 0.71 | 0.004 | 2.70E-12 | 41.48 | 3.92E-04 | TRUE | 9.60E-08 |
| rs55891451 | C | A | 0.03 | 0.20 | 0.005 | 5.60E-11 | 38.65 | 3.64E-04 | TRUE | 9.40E-07 |
| rs58542926 | T | C | -0.12 | 0.07 | 0.01 | 1.40E-57 | 224.22 | 2.12E-03 | TRUE | 6.72E-26 |
| rs629301 | T | G | 0.04 | 0.78 | 0.005 | 1.40E-19 | 76.11 | 7.23E-04 | TRUE | 1.69E-08 |
| rs638714 | T | G | -0.04 | 0.35 | 0.004 | 5.40E-20 | 74.43 | 7.00E-04 | TRUE | 1.44E-08 |
| rs673335 | C | T | -0.06 | 0.16 | 0.01 | 8.10E-25 | 95.00 | 9.00E-04 | TRUE | 1.26E-12 |
| rs6931604 | T | C | 0.02 | 0.60 | 0.004 | 2.70E-08 | 29.01 | 2.73E-04 | TRUE | 1.58E-03 |
| rs72836561 | T | C | -0.06 | 0.03 | 0.01 | 3.60E-08 | 28.00 | 2.66E-04 | TRUE | 1.40E-06 |
| rs73045691 | A | G | 0.03 | 0.30 | 0.005 | 3.60E-10 | 41.29 | 3.55E-04 | TRUE | 2.07E-05 |
| Supplementary Table S13:Harmonized SNPs for the association between DHA and CRP cont. (2) | | | | | | | | | | |
| rs73109460 | A | G | -0.04 | 0.12 | 0.01 | 7.30E-11 | 33.82 | 3.17E-04 | TRUE | 1.71E-03 |
| rs7412 | T | C | -0.08 | 0.08 | 0.01 | 5.40E-26 | 100.67 | 9.45E-04 | TRUE | 9.82E-01 |
| rs77960347 | G | A | 0.14 | 0.01 | 0.02 | 2.50E-17 | 60.50 | 5.75E-04 | TRUE | 3.22E-08 |
| rs7924036 | T | G | 0.04 | 0.50 | 0.004 | 4.80E-21 | 73.12 | 6.94E-04 | TRUE | 5.09E-10 |
| rs9304381 | T | C | 0.05 | 0.82 | 0.01 | 5.80E-23 | 86.92 | 8.22E-04 | TRUE | 5.73E-12 |
| rs9987289 | G | A | 0.06 | 0.91 | 0.01 | 1.30E-17 | 64.54 | 6.11E-04 | FALSE | 2.28E-01 |
| *SNPS, Single nucleotide polymorphisms; DHA, Docosahexaenoic acid; CRP, C-reactive protein; EAF, Effect Allele Frequency; SE, Standard error* | | | | | | | | | | |

| **Supplementary Table S14:Harmonised SNPs for the association between LA and CRP** | | | | | | | | | | | | | | |
| --- | --- | --- | --- | --- | --- | --- | --- | --- | --- | --- | --- | --- | --- | --- |
| **SNP** | **Effect Allele** | **Other Allele** | **Beta** | **EAF** | **Proxy Outcome** | **Proxy SNP** | **Proxy Effect Allele** | **Proxy Other Allele** | **p-value** | **SE** | **F statistic** | **R^2^ exposure** | **Steiger direction** | **Steiger p-value** |
| rs1002687 | A | G | 0.09 | 0.64 | NA | NA | NA | NA | 1.80E-97 | 0.004 | 398.69 | 3.65E-03 | TRUE | 1.41E-35 |
| rs10838724 | T | G | -0.03 | 0.37 | TRUE | rs4752845 | C | T | 1.60E-10 | 0.004 | 36.86 | 3.32E-04 | TRUE | 4.99E-02 |
| rs115478735 | T | A | 0.04 | 0.18 | TRUE | rs651007 | T | C | 1.20E-17 | 0.01 | 63.28 | 5.82E-04 | TRUE | 3.34E-02 |
| rs142158911 | A | G | -0.09 | 0.12 | TRUE | rs6511720 | T | G | 1.20E-48 | 0.01 | 199.22 | 1.81E-03 | TRUE | 6.69E-14 |
| rs1461729 | G | A | 0.07 | 0.90 | NA | NA | NA | NA | 5.40E-26 | 0.01 | 103.31 | 9.48E-04 | TRUE | 4.43E-01 |
| rs174564 | G | A | 0.08 | 0.35 | TRUE | rs174583 | T | C | 5.10E-88 | 0.004 | 371.34 | 3.42E-03 | TRUE | 2.59E-33 |
| rs2378390 | A | G | -0.03 | 0.14 | TRUE | rs2277862 | T | C | 7.00E-09 | 0.01 | 29.39 | 2.69E-04 | TRUE | 1.48E-03 |
| rs2740488 | C | A | -0.05 | 0.27 | TRUE | rs2575876 | A | G | 1.00E-25 | 0.005 | 104.62 | 9.58E-04 | TRUE | 1.10E-10 |
| rs4008004 | A | C | 0.03 | 0.22 | NA | NA | NA | NA | 1.90E-10 | 0.005 | 37.93 | 3.46E-04 | TRUE | 2.87E-05 |
| rs4299376 | T | G | -0.04 | 0.68 | TRUE | rs6544713 | T | C | 2.50E-19 | 0.004 | 75.61 | 6.93E-04 | TRUE | 4.28E-06 |
| rs4665972 | C | T | -0.05 | 0.60 | TRUE | rs1260326 | T | C | 1.30E-35 | 0.004 | 145.10 | 1.32E-03 | FALSE | 1.24E-08 |
| rs4704210 | C | G | 0.05 | 0.37 | TRUE | rs4704219 | T | C | 2.00E-35 | 0.004 | 140.95 | 1.30E-03 | TRUE | 6.10E-16 |
| rs4947302 | T | C | 0.05 | 0.06 | TRUE | rs9391709 | C | G | 4.80E-10 | 0.01 | 29.45 | 2.70E-04 | TRUE | 3.24E-04 |
| rs56322906 | A | G | -0.09 | 0.04 | TRUE | rs737338 | T | C | 2.90E-15 | 0.01 | 60.86 | 5.59E-04 | TRUE | 3.04E-08 |
| rs58542926 | T | C | -0.11 | 0.07 | TRUE | rs3794991 | T | C | 7.50E-48 | 0.01 | 191.30 | 1.76E-03 | TRUE | 1.91E-14 |
| rs602633 | G | T | 0.05 | 0.78 | NA | NA | NA | NA | 7.50E-28 | 0.005 | 110.65 | 1.02E-03 | TRUE | 2.19E-08 |
| rs633695 | G | A | 0.07 | 0.29 | NA | NA | NA | NA | 2.00E-54 | 0.004 | 231.62 | 2.11E-03 | TRUE | 2.24E-22 |
| rs6471717 | A | G | -0.03 | 0.66 | NA | NA | NA | NA | 1.40E-13 | 0.004 | 48.94 | 4.48E-04 | TRUE | 4.33E-07 |
| rs693 | A | G | 0.06 | 0.52 | NA | NA | NA | NA | 3.40E-54 | 0.004 | 217.11 | 1.99E-03 | TRUE | 2.49E-24 |
| rs7139079 | A | G | -0.03 | 0.59 | TRUE | rs2393791 | C | T | 4.80E-10 | 0.004 | 37.42 | 3.43E-04 | FALSE | 8.86E-103 |
| rs740516 | G | C | -0.03 | 0.15 | NA | NA | NA | NA | 1.90E-09 | 0.01 | 33.40 | 3.05E-04 | TRUE | 5.07E-05 |
| rs7816447 | C | T | -0.05 | 0.10 | NA | NA | NA | NA | 8.20E-13 | 0.01 | 49.50 | 4.57E-04 | TRUE | 6.79E-04 |
| rs9302635 | C | T | -0.03 | 0.18 | NA | NA | NA | NA | 3.40E-08 | 0.01 | 27.58 | 2.55E-04 | TRUE | 6.93E-03 |
| rs9304381 | T | C | 0.06 | 0.82 | TRUE | rs7240405 | A | G | 1.00E-33 | 0.01 | 135.15 | 1.24E-03 | TRUE | 1.58E-14 |
| rs964184 | C | G | -0.15 | 0.87 | NA | NA | NA | NA | 2.00E-136 | 0.01 | 564.21 | 5.15E-03 | TRUE | 4.12E-54 |
| *SNPS, Single nucleotide polymorphisms; LA, Linolic Acid; CRP, C-reactive protein; EAF, Effect Allele Frequency; SE, Standard error.* | | | | | | | | | | | | | | |

| **Supplementary Table S15: Harmonised SNPs for the association between total n-3 and CRP (1)** | | | | | | | | | | |
| --- | --- | --- | --- | --- | --- | --- | --- | --- | --- | --- |
| **SNP** | **Effect Allele** | **Other Allele** | **Beta** | **EAF** | **SE** | **p-value** | **F statistic** | **R^2^ exposure** | **Steiger direction** | **Steiger p-value** |
| rs10455872 | G | A | -0.06 | 0.08 | 2.80E-17 | 0.01 | 66.36 | 0.001 | TRUE | 5.61E-13 |
| rs11230829 | G | A | -0.10 | 0.03 | 3.40E-12 | 0.01 | 65.90 | 0.0004 | TRUE | 1.02E-09 |
| rs11242109 | T | G | 0.02 | 0.48 | 2.40E-09 | 0.00 | 33.24 | 0.0003 | TRUE | 4.23E-04 |
| rs112875651 | A | G | -0.09 | 0.39 | 3.50E-98 | 0.004 | 420.12 | 0.004 | TRUE | 1.25E-50 |
| rs1132899 | C | T | 0.03 | 0.51 | 8.60E-11 | 0.004 | 42.18 | 0.0004 | TRUE | 9.33E-07 |
| rs11563251 | T | C | 0.03 | 0.11 | 3.20E-08 | 0.01 | 27.68 | 0.0003 | TRUE | 1.63E-06 |
| rs1167998 | A | C | 0.07 | 0.64 | 3.60E-66 | 0.004 | 268.89 | 0.002 | TRUE | 1.37E-37 |
| rs11681659 | T | C | -0.03 | 0.72 | 2.00E-08 | 0.004 | 29.50 | 0.0003 | TRUE | 1.01E-04 |
| rs117143374 | C | T | -0.04 | 0.14 | 2.20E-10 | 0.01 | 38.63 | 0.0003 | TRUE | 4.62E-08 |
| rs117733303 | G | A | -0.12 | 0.02 | 1.40E-15 | 0.02 | 56.21 | 0.001 | TRUE | 8.42E-10 |
| rs12226389 | C | T | -0.05 | 0.19 | 1.10E-22 | 0.01 | 89.19 | 0.001 | TRUE | 2.98E-16 |
| rs1260326 | C | T | -0.08 | 0.60 | 8.40E-88 | 0.004 | 371.62 | 0.003 | TRUE | 1.03E-02 |
| rs13424225 | T | G | 0.02 | 0.45 | 2.20E-08 | 0.004 | 27.87 | 0.0003 | TRUE | 1.22E-04 |
| rs139974673 | C | T | 0.12 | 0.03 | 2.30E-21 | 0.01 | 80.89 | 0.001 | TRUE | 6.21E-05 |
| rs143355652 | T | C | -0.15 | 0.01 | 9.40E-14 | 0.02 | 56.62 | 0.0005 | TRUE | 1.24E-11 |
| rs157592 | C | A | 0.03 | 0.19 | 3.60E-09 | 0.01 | 28.07 | 0.0002 | FALSE | 2.33E-129 |
| rs1672811 | C | T | 0.03 | 0.75 | 3.00E-08 | 0.005 | 27.47 | 0.0002 | TRUE | 8.25E-05 |
| rs16940904 | T | C | -0.04 | 0.23 | 3.90E-14 | 0.005 | 50.90 | 0.0005 | TRUE | 1.82E-09 |
| rs174564 | G | A | -0.34 | 0.35 | 1.00E-200 | 0.004 | 6243.53 | 0.05 | TRUE | 0.00E+00 |
| rs182611493 | G | A | -0.21 | 0.01 | 1.10E-27 | 0.02 | 125.01 | 0.001 | TRUE | 3.35E-17 |
| rs261290 | C | T | -0.11 | 0.65 | 3.90E-161 | 0.004 | 684.36 | 0.01 | TRUE | 3.90E-103 |
| rs3018731 | G | A | -0.04 | 0.72 | 2.00E-14 | 0.005 | 58.23 | 0.001 | TRUE | 8.71E-12 |
| rs34663616 | A | C | 0.04 | 0.14 | 4.40E-10 | 0.01 | 34.75 | 0.0003 | TRUE | 8.09E-07 |
| rs35135293 | T | C | -0.02 | 0.52 | 3.90E-08 | 0.004 | 25.06 | 0.0002 | TRUE | 5.91E-05 |
| rs4000713 | A | G | -0.03 | 0.30 | 1.00E-11 | 0.004 | 39.77 | 0.0004 | TRUE | 1.91E-08 |
| rs55891451 | C | A | 0.03 | 0.20 | 4.60E-12 | 0.01 | 43.30 | 0.0004 | TRUE | 2.81E-07 |
| rs58542926 | T | C | -0.17 | 0.07 | 1.40E-113 | 0.01 | 468.55 | 0.004 | TRUE | 5.40E-61 |
| rs6129624 | A | G | -0.03 | 0.34 | 5.10E-10 | 0.004 | 34.02 | 0.0003 | TRUE | 2.21E-05 |
| rs62466318 | T | C | -0.07 | 0.20 | 1.20E-45 | 0.01 | 194.78 | 0.002 | TRUE | 1.73E-18 |
| rs629301 | T | G | 0.04 | 0.78 | 1.30E-14 | 0.005 | 58.26 | 0.001 | TRUE | 7.82E-06 |
| rs633695 | G | A | 0.08 | 0.29 | 9.10E-80 | 0.004 | 336.77 | 0.003 | TRUE | 6.50E-58 |
| rs6601924 | C | T | 0.04 | 0.85 | 8.50E-10 | 0.01 | 36.89 | 0.0003 | TRUE | 1.91E-08 |
| *Supplementary Table S15: Harmonised SNPs for the association between total n-3 and CRP cont. (2)* | | | | | | | | | | |
| rs6693447 | G | T | 0.02 | 0.46 | 4.80E-09 | 0.004 | 30.11 | 0.0003 | TRUE | 4.76E-07 |
| rs673335 | C | T | -0.07 | 0.16 | 1.10E-34 | 0.01 | 138.76 | 0.001 | TRUE | 9.23E-19 |
| rs6882345 | A | G | 0.03 | 0.63 | 1.90E-13 | 0.004 | 44.60 | 0.0004 | TRUE | 2.38E-09 |
| rs73109460 | A | G | -0.03 | 0.12 | 9.20E-10 | 0.01 | 30.44 | 0.0003 | TRUE | 5.77E-03 |
| rs737338 | T | C | -0.07 | 0.04 | 3.50E-11 | 0.01 | 41.24 | 0.0004 | TRUE | 9.78E-08 |
| rs77960347 | G | A | 0.16 | 0.01 | 7.20E-22 | 0.02 | 78.66 | 0.001 | TRUE | 1.40E-10 |
| rs7819706 | G | A | -0.04 | 0.12 | 1.80E-10 | 0.01 | 37.71 | 0.0003 | TRUE | 2.68E-05 |
| rs7924036 | T | G | 0.02 | 0.50 | 5.50E-10 | 0.00 | 31.36 | 0.0003 | TRUE | 9.44E-04 |
| rs7970695 | A | G | -0.03 | 0.62 | 1.20E-10 | 0.004 | 34.69 | 0.0003 | FALSE | 1.28E-137 |
| rs9304381 | T | C | 0.05 | 0.82 | 5.20E-24 | 0.01 | 95.67 | 0.001 | TRUE | 8.51E-13 |
| rs9987289 | G | A | 0.06 | 0.91 | 3.20E-16 | 0.01 | 61.10 | 0.001 | FALSE | 0.12 |
| *SNPS, Single nucleotide polymorphisms; n-3, Total omega-3; CRP, C-reactive protein; EAF, Effect Allele Frequency; SE, Standard error.* | | | | | | | | | | |

| **Supplementary Table S16: Harmonized SNPs for the association between total n-6 and CRP (1)** | | | | | | | | | | |
| --- | --- | --- | --- | --- | --- | --- | --- | --- | --- | --- |
| **SNP** | **Effect Allele** | **Other Allele** | **Beta** | **EAF** | **SE** | **p-value** | **F statistic** | **R^2^ exposure** | **Steiger direction** | **Steiger p-value** |
| rs1002687 | A | G | 0.09 | 0.64 | 0.004 | 1.00E-107 | 437.62 | 0.004 | TRUE | 2.97E-66 |
| rs1065853 | T | G | -0.20 | 0.08 | 0.007 | 3.60E-160 | 676.17 | 0.006 | TRUE | 2.06E-51 |
| rs1081105 | C | A | 0.12 | 0.03 | 0.012 | 1.80E-22 | 87.22 | 0.001 | FALSE | 5.90E-09 |
| rs112875651 | A | G | -0.06 | 0.39 | 0.004 | 2.20E-53 | 222.88 | 0.002 | TRUE | 2.43E-23 |
| rs11789603 | T | C | 0.05 | 0.11 | 0.006 | 9.70E-14 | 51.16 | 0.000 | TRUE | 5.15E-10 |
| rs1260326 | C | T | -0.06 | 0.60 | 0.004 | 3.90E-55 | 227.58 | 0.002 | FALSE | 2.17E-01 |
| rs12740374 | T | G | -0.06 | 0.22 | 0.005 | 1.50E-32 | 129.94 | 0.001 | TRUE | 4.81E-16 |
| rs13108218 | G | A | -0.04 | 0.62 | 0.004 | 3.60E-18 | 67.35 | 0.001 | TRUE | 2.72E-07 |
| rs142158911 | A | G | -0.09 | 0.12 | 0.006 | 5.20E-52 | 211.68 | 0.002 | TRUE | 1.15E-40 |
| rs1461729 | G | A | 0.08 | 0.90 | 0.007 | 2.80E-36 | 145.67 | 0.001 | TRUE | 1.70E-03 |
| rs1800961 | T | C | -0.07 | 0.03 | 0.012 | 3.30E-10 | 37.35 | 0.000 | FALSE | 9.72E-02 |
| rs183130 | T | C | 0.06 | 0.32 | 0.004 | 1.40E-48 | 190.95 | 0.002 | TRUE | 4.00E-32 |
| rs200730299 | C | A | -0.04 | 0.19 | 0.005 | 7.10E-12 | 48.10 | 0.000 | TRUE | 2.22E-09 |
| rs2378390 | A | G | -0.03 | 0.14 | 0.006 | 3.20E-09 | 30.93 | 0.000 | TRUE | 4.38E-06 |
| rs261290 | C | T | -0.10 | 0.65 | 0.004 | 1.00E-116 | 486.78 | 0.004 | TRUE | 8.00E-72 |
| rs2737245 | T | G | -0.03 | 0.28 | 0.005 | 1.40E-09 | 34.91 | 0.000 | TRUE | 3.94E-01 |
| rs2740488 | C | A | -0.05 | 0.27 | 0.005 | 5.40E-28 | 112.79 | 0.001 | TRUE | 4.43E-20 |
| rs2986164 | A | G | -0.03 | 0.54 | 0.004 | 3.40E-09 | 35.78 | 0.000 | TRUE | 5.42E-06 |
| rs3770586 | T | C | -0.02 | 0.48 | 0.004 | 7.10E-09 | 30.31 | 0.000 | TRUE | 5.88E-02 |
| rs4008004 | A | C | 0.03 | 0.22 | 0.005 | 8.40E-12 | 43.15 | 0.000 | TRUE | 2.68E-09 |
| rs4299376 | T | G | -0.04 | 0.68 | 0.004 | 1.10E-16 | 63.10 | 0.001 | TRUE | 1.02E-09 |
| rs4439799 | T | C | 0.02 | 0.50 | 0.004 | 1.30E-08 | 28.92 | 0.000 | TRUE | 2.00E-06 |
| rs534417 | G | A | 0.04 | 0.88 | 0.006 | 9.30E-11 | 38.35 | 0.000 | TRUE | 2.14E-08 |
| rs55747707 | A | G | -0.05 | 0.20 | 0.005 | 1.70E-22 | 91.41 | 0.001 | TRUE | 2.06E-06 |
| rs56322906 | A | G | -0.10 | 0.04 | 0.011 | 1.20E-19 | 78.65 | 0.001 | TRUE | 1.26E-14 |
| rs5754102 | A | C | -0.03 | 0.18 | 0.005 | 9.90E-10 | 34.38 | 0.000 | TRUE | 1.58E-07 |
| rs58542926 | T | C | -0.13 | 0.07 | 0.008 | 2.50E-65 | 260.26 | 0.002 | TRUE | 1.76E-30 |
| rs633695 | G | A | 0.07 | 0.29 | 0.004 | 1.30E-59 | 250.77 | 0.002 | TRUE | 2.14E-43 |
| rs6471717 | A | G | -0.03 | 0.66 | 0.004 | 4.00E-12 | 43.31 | 0.000 | TRUE | 9.03E-09 |
| rs6547409 | T | C | -0.08 | 0.05 | 0.009 | 2.40E-20 | 74.07 | 0.001 | TRUE | 1.41E-12 |
| rs6602911 | T | C | 0.03 | 0.36 | 0.004 | 1.30E-09 | 35.15 | 0.000 | TRUE | 2.71E-08 |
| *Supplementary Table S16: Harmonized SNPs for the association between total n-6 and CRP cont. (2)* | | | | | | | | | | |
| rs672889 | G | T | 0.08 | 0.86 | 0.006 | 1.30E-41 | 161.75 | 0.002 | TRUE | 3.66E-33 |
| rs6882345 | A | G | 0.04 | 0.63 | 0.004 | 1.20E-27 | 107.73 | 0.001 | TRUE | 1.89E-21 |
| rs6934962 | T | C | 0.02 | 0.40 | 0.004 | 2.20E-08 | 28.78 | 0.000 | TRUE | 1.51E-01 |
| rs6938647 | C | A | -0.05 | 0.78 | 0.005 | 1.90E-23 | 90.93 | 0.001 | TRUE | 5.87E-13 |
| rs7139079 | A | G | -0.03 | 0.59 | 0.004 | 3.30E-13 | 49.13 | 0.000 | FALSE | 2.14E-106 |
| rs72997616 | A | C | -0.05 | 0.09 | 0.007 | 1.60E-13 | 52.13 | 0.000 | TRUE | 2.27E-06 |
| rs75406471 | A | G | -0.03 | 0.15 | 0.006 | 2.70E-08 | 29.67 | 0.000 | TRUE | 3.39E-07 |
| rs7750288 | G | A | 0.02 | 0.29 | 0.004 | 1.30E-08 | 29.12 | 0.000 | TRUE | 1.04E-03 |
| rs77960347 | G | A | 0.28 | 0.01 | 0.018 | 2.80E-56 | 228.54 | 0.002 | TRUE | 3.55E-35 |
| rs79429216 | A | G | 0.15 | 0.01 | 0.018 | 1.30E-17 | 65.71 | 0.001 | TRUE | 1.88E-05 |
| rs870526 | T | C | -0.03 | 0.52 | 0.004 | 7.70E-16 | 58.40 | 0.001 | TRUE | 2.20E-09 |
| rs9295128 | T | G | -0.20 | 0.02 | 0.016 | 3.40E-36 | 146.97 | 0.001 | TRUE | 3.28E-24 |
| rs9304381 | T | C | 0.07 | 0.82 | 0.005 | 7.20E-42 | 168.83 | 0.002 | TRUE | 1.19E-24 |
| *SNPS, Single nucleotide polymorphisms; n-6, Total omega-6; CRP, C-reactive protein; EAF, Effect Allele Frequency; SE, Standard error.* | | | | | | | | | | |

| **Supplementary Table S17: Harmonized SNPs for the association between DHA and IL-6** | | | | | | | | | | |
| --- | --- | --- | --- | --- | --- | --- | --- | --- | --- | --- |
| **SNP** | **Effect Allele** | **Other Allele** | **Beta** | **EAF** | **SE** | **p-value** | **F statistic** | **R^2^ exposure** | **Steiger direction** | **Steiger p-value** |
| rs1260326 | C | T | -0.05 | 0.60 | 0.004 | 8.70E-30 | 120.00 | 1.14E-03 | TRUE | 7.98E-05 |
| rs1560390 | C | T | -0.05 | 0.22 | 0.005 | 2.50E-26 | 99.33 | 9.31E-04 | TRUE | 1.27E-05 |
| rs16940904 | T | C | -0.04 | 0.23 | 0.005 | 2.10E-17 | 63.08 | 5.93E-04 | TRUE | 1.71E-04 |
| rs1800978 | G | C | -0.03 | 0.12 | 0.006 | 4.80E-08 | 28.22 | 2.65E-04 | TRUE | 8.79E-02 |
| rs2278426 | T | C | -0.06 | 0.04 | 0.011 | 3.70E-08 | 28.43 | 2.70E-04 | TRUE | 1.23E-02 |
| rs273912 | T | G | 0.03 | 0.71 | 0.004 | 6.60E-10 | 31.76 | 3.02E-04 | TRUE | 7.26E-02 |
| rs325 | C | T | 0.04 | 0.10 | 0.007 | 3.80E-09 | 31.22 | 2.97E-04 | TRUE | 3.10E-03 |
| rs3764261 | A | C | 0.04 | 0.32 | 0.004 | 1.90E-21 | 77.57 | 7.35E-04 | TRUE | 1.41E-06 |
| rs4986970 | T | A | -0.07 | 0.03 | 0.011 | 1.30E-10 | 35.94 | 3.39E-04 | TRUE | 2.59E-03 |
| rs629301 | T | G | 0.04 | 0.78 | 0.005 | 1.40E-19 | 76.11 | 7.23E-04 | TRUE | 1.09E-05 |
| rs9987289 | G | A | 0.06 | 0.91 | 0.007 | 1.30E-17 | 64.54 | 6.11E-04 | TRUE | 1.33E-03 |
| *SNPS, Single nucleotide polymorphisms; DHA, Docosahexaenoi acidc; IL-6, Interleukin-6; EAF, Effect Allele Frequency; SE, Standard error* | | | | | | | | | | |

| **Supplementary Table S18: Harmonized SNPs for the association between LA and IL-6** | | | | | | | | | | |
| --- | --- | --- | --- | --- | --- | --- | --- | --- | --- | --- |
| **SNP** | **Effect Allele** | **Other Allele** | **Beta** | **EAF** | **SE** | **p-value** | **F statistic** | **R^2^ exposure** | **Steiger direction** | **Steiger p-value** |
| rs1002687 | A | G | 0.09 | 0.64 | 1.80E-97 | 0.004 | 398.69 | 3.65E-03 | TRUE | 1.78E-30 |
| rs11789603 | T | C | 0.05 | 0.11 | 1.70E-12 | 0.007 | 46.02 | 4.22E-04 | TRUE | 1.53E-01 |
| rs1461729 | G | A | 0.07 | 0.90 | 5.40E-26 | 0.007 | 103.31 | 9.48E-04 | TRUE | 2.38E-06 |
| rs4008004 | A | C | 0.03 | 0.22 | 1.90E-10 | 0.005 | 37.93 | 3.46E-04 | TRUE | 5.11E-04 |
| rs4299376 | T | G | -0.04 | 0.68 | 2.50E-19 | 0.004 | 75.61 | 6.93E-04 | TRUE | 1.47E-05 |
| rs602633 | G | T | 0.05 | 0.78 | 7.50E-28 | 0.005 | 110.65 | 1.02E-03 | TRUE | 7.92E-08 |
| rs633695 | G | A | 0.07 | 0.29 | 2.00E-54 | 0.004 | 231.62 | 2.11E-03 | TRUE | 3.83E-18 |
| rs6471717 | A | G | -0.03 | 0.66 | 1.40E-13 | 0.004 | 48.94 | 4.48E-04 | TRUE | 2.20E-02 |
| rs693 | A | G | 0.06 | 0.52 | 3.40E-54 | 0.004 | 217.11 | 1.99E-03 | TRUE | 3.00E-15 |
| rs740516 | G | C | -0.03 | 0.15 | 1.90E-09 | 0.006 | 33.40 | 3.05E-04 | TRUE | 7.94E-03 |
| rs7816447 | C | T | -0.05 | 0.10 | 8.20E-13 | 0.007 | 49.50 | 4.57E-04 | TRUE | 2.51E-04 |
| rs9302635 | C | T | -0.03 | 0.18 | 3.40E-08 | 0.005 | 27.58 | 2.55E-04 | TRUE | 1.65E-02 |
| rs964184 | C | G | -0.15 | 0.87 | 2.00E-136 | 0.006 | 564.21 | 5.15E-03 | TRUE | 4.86E-37 |
| *SNPS, Single nucleotide polymorphisms; LA, Linolic acid; IL-6, Interleukin-6; EAF, Effect Allele Frequency; SE, Standard error* | | | | | | | | | | |

| **Supplementary Table S19: Harmonized SNPs for the association between total n-3 and IL-6** | | | | | | | | | | |
| --- | --- | --- | --- | --- | --- | --- | --- | --- | --- | --- |
| **SNP** | **Effect Allele** | **Other Allele** | **Beta** | **EAF** | **SE** | **p-value** | **F statistic** | **R^2^ exposure** | **Steiger direction** | **Steiger p-value** |
| rs10184054 | G | C | -0.04 | 0.22 | 5.60E-15 | 0.005 | 52.28 | 4.80E-04 | TRUE | 1.45E-03 |
| rs10455872 | G | A | -0.06 | 0.08 | 2.80E-17 | 0.008 | 66.36 | 6.07E-04 | TRUE | 4.51E-04 |
| rs11563251 | T | C | 0.03 | 0.11 | 3.20E-08 | 0.006 | 27.68 | 2.54E-04 | TRUE | 2.75E-03 |
| rs1167998 | A | C | 0.07 | 0.64 | 3.60E-66 | 0.004 | 268.89 | 2.44E-03 | TRUE | 8.56E-21 |
| rs1260326 | C | T | -0.08 | 0.60 | 8.40E-88 | 0.004 | 371.62 | 3.38E-03 | TRUE | 8.05E-18 |
| rs1672811 | C | T | 0.03 | 0.75 | 3.00E-08 | 0.005 | 27.47 | 2.50E-04 | TRUE | 3.08E-02 |
| rs16940904 | T | C | -0.04 | 0.23 | 3.90E-14 | 0.005 | 50.90 | 4.61E-04 | TRUE | 1.32E-03 |
| rs1800978 | G | C | -0.04 | 0.12 | 5.20E-09 | 0.006 | 34.84 | 3.16E-04 | TRUE | 4.68E-02 |
| rs3018731 | G | A | -0.04 | 0.72 | 2.00E-14 | 0.005 | 58.23 | 5.21E-04 | TRUE | 1.82E-05 |
| rs6129624 | A | G | -0.03 | 0.34 | 5.10E-10 | 0.004 | 34.02 | 3.01E-04 | TRUE | 1.71E-03 |
| rs629301 | T | G | 0.04 | 0.78 | 1.30E-14 | 0.005 | 58.26 | 5.34E-04 | TRUE | 2.33E-04 |
| rs633695 | G | A | 0.08 | 0.29 | 9.10E-80 | 0.004 | 336.77 | 3.04E-03 | TRUE | 1.56E-25 |
| rs6601924 | C | T | 0.04 | 0.85 | 8.50E-10 | 0.006 | 36.89 | 3.36E-04 | TRUE | 3.07E-03 |
| rs737338 | T | C | -0.07 | 0.04 | 3.50E-11 | 0.011 | 41.24 | 3.77E-04 | TRUE | 3.40E-03 |
| rs7819706 | G | A | -0.04 | 0.12 | 1.80E-10 | 0.006 | 37.71 | 3.45E-04 | TRUE | 5.19E-04 |
| rs964184 | C | G | -0.12 | 0.87 | 8.90E-87 | 0.006 | 361.40 | 3.31E-03 | TRUE | 1.42E-23 |
| rs9987289 | G | A | 0.06 | 0.91 | 3.20E-16 | 0.007 | 61.10 | 5.59E-04 | TRUE | 2.68E-03 |
| *SNPS, Single nucleotide polymorphisms; n-3, Total omega-3; IL-6, Interleukin-6; EAF, Effect Allele Frequency; SE, Standard error* | | | | | | | | | | |

| **Supplemenmtary Table S20: Harmonized SNPs for the association between total n-6 and IL-6** | | | | | | | | | | |
| --- | --- | --- | --- | --- | --- | --- | --- | --- | --- | --- |
| **SNP** | **Effect Allele** | **Other Allele** | **Beta** | **EAF** | **SE** | **p-value** | **F statistic** | **R^2^ exposure** | **Steiger direction** | **Steiger p-value** |
| rs1002687 | A | G | 0.090971 | 0.644748 | 0.004213 | 1.00E-107 | 437.6214 | 0.004038 | TRUE | 1.49E-33 |
| rs11789603 | T | C | 0.04788 | 0.108831 | 0.006491 | 9.70E-14 | 51.15959 | 0.000473 | TRUE | 0.097106 |
| rs1260326 | C | T | -0.06426 | 0.60401 | 0.004122 | 3.90E-55 | 227.579 | 0.002109 | TRUE | 3.71E-10 |
| rs12740374 | T | G | -0.05725 | 0.221071 | 0.004852 | 1.50E-32 | 129.9393 | 0.001209 | TRUE | 2.63E-09 |
| rs1461729 | G | A | 0.083547 | 0.899221 | 0.006706 | 2.80E-36 | 145.6664 | 0.001348 | TRUE | 5.09E-09 |
| rs1800961 | T | C | -0.07446 | 0.030192 | 0.011781 | 3.30E-10 | 37.34588 | 0.000347 | TRUE | 0.000781 |
| rs2737245 | T | G | -0.02748 | 0.278598 | 0.004508 | 1.40E-09 | 34.90787 | 0.000323 | TRUE | 0.004484 |
| rs3734854 | A | G | 0.047712 | 0.064679 | 0.008213 | 5.90E-11 | 31.68167 | 0.000293 | TRUE | 0.008042 |
| rs3817335 | A | T | -0.02776 | 0.350559 | 0.004215 | 9.80E-12 | 40.37145 | 0.000377 | TRUE | 0.000622 |
| rs4008004 | A | C | 0.032959 | 0.221842 | 0.004875 | 8.40E-12 | 43.1469 | 0.000397 | TRUE | 0.000195 |
| rs4299376 | T | G | -0.03539 | 0.676279 | 0.00432 | 1.10E-16 | 63.09935 | 0.000583 | TRUE | 8.83E-05 |
| rs5754102 | A | C | -0.0316 | 0.183245 | 0.005274 | 9.90E-10 | 34.38305 | 0.000312 | TRUE | 0.001479 |
| rs633695 | G | A | 0.072518 | 0.292348 | 0.00445 | 1.30E-59 | 250.7677 | 0.002303 | TRUE | 1.13E-19 |
| rs6471717 | A | G | -0.02903 | 0.663091 | 0.004278 | 4.00E-12 | 43.31267 | 0.0004 | TRUE | 0.03859 |
| rs6547409 | T | C | -0.08135 | 0.051258 | 0.009248 | 2.40E-20 | 74.06892 | 0.000672 | TRUE | 8.31E-07 |
| rs740516 | G | C | -0.03157 | 0.151132 | 0.005659 | 1.40E-08 | 29.41866 | 0.000271 | TRUE | 0.013744 |
| rs870526 | T | C | -0.03189 | 0.521426 | 0.00404 | 7.70E-16 | 58.40176 | 0.000542 | TRUE | 0.000119 |
| rs964184 | C | G | -0.1389 | 0.867229 | 0.005944 | 1.10E-125 | 513.1765 | 0.004725 | TRUE | 6.56E-34 |
| *SNPS, Single nucleotide polymorphisms; n-6, Total omega-6; IL-6, Interleukin-6; EAF, Effect Allele Frequency; SE, Standard error* | | | | | | | | | | |

| **Supplementary Table S21: Description of MR, MR methods and sensitivity analyses as seen in Crick et al.(2) (1)** | | |
| --- | --- | --- |
| **Method** | **Description** | |
| Main analysis | | |
| Mendelian Randomisation (MR) | MR is an instrumental variable approach , using genetic variants as instruments for a modifiable exposure. It interrogates the causal effect of an exposure on an outcome, by utilising the random assortment of genetic variants (during gamete formation) from parents to offspring and as a result, the potential bias from confounding and reverse causation are minimised. MR is bound by three assumptions: (i) the genetic variants are statistically strongly associated with the exposure of interest and relevant to the population to which inference is being made (the relevance assumption); (ii) there is no confounding of the SNP-outcome association; and (iii) any effect of the genetic instrument on the outcome is only via the exposure(3) (supplementary figure 1). | |
| Inverse Variance Weighted (IVW) | A Wald ratio estimate is calculated for each genetic variant and summarised using the weighted regression of the SNP-exposure estimates on SNP-outcome estimates, where the intercept was constrained to zero. IVW assumes that there is no correlation between the association of the SNP exposure and SNP-pleiotropic path (the Instrument Strength Independent of Direct Effect (InSIDE) assumption) in the presence of horizontal pleiotropic paths. | |
| **Methods to explore MR assumption** | | |
| F-Statistic | Relevance assumption and weak instrument bias | Investigating the strength of genetic instruments using the F-statistics and proportion explained (R^2^) by each SNP. F-statistics >10 indicate that the estimates are not substantially biased by weak instruments(4). |
| MR-Egger | Horizontal pleiotropy. | Unlike the IVW, MR-Egger does not constrain the regression line to go through zero. Therefore, the MR-Egger regression represents the estimate of the causal effect controlling for unbalanced horizontal pleiotropy. A non-null MR-Egger intercept provides evidence of horizontal pleiotropy(5). MR-Egger has less statistical power than IVW and there should be no violation of the InSIDE assumption |
| Weighted Median and weighted mode | Horizontal pleiotropy | The weighted median and weighted mode assume that SNPs that display no pleiotropic effects, will show more uniform and homogenous effects on the exposure and outcome and make them cluster toward the median/mode. In contrast, the pleiotropic effects of certain SNPs on the outcome are less |
| *Supplementary Table S21: Description of MR, MR methods and sensitivity analyses as seen in Crick et al.(2) cont. (2)* | | |
|  |  | likely to converge on common median or model estimate (6, 7).  The weighted median provides an unbiased estimate when up to 50% of the SNPs used in the instrument violate the IV assumption whereas, the weighted mode uses the mode of the IVW empirical density function as the effect estimate. |
| Cochran’s Q / Rucker’s Q statistic | Horizontal pleiotropy | Test for between-SNP heterogeneity in the IVW and MR-Egger analysis. |
| MR-Egger intercept | Horizontal pleiotropy | Under the assumption that pleiotropy is independent of the SNP-exposure association will give evidence for pleiotropic effects. |
| Sample definition/ underlying population | Confounding (via population stratification) | To mitigate bias caused by population stratification, which can confound the genetic instrument-outcome association and therefore violate the independence assumption, we restricted analyses to GWAS’ of the same underlying population (European ancestry participants only). |
| Radial MR | Horizontal pleiotropy | The method is similar to IVW but using a simulation-based approach fits a radial IVW model and provides an effect estimate, allowing outliers to be identified using Cochran’s Q statistic. The same can be done for a radial MR-Egger model. |
| Steiger filtering | Directionality of effects | Determines whether the proportion of variance explained by each SNP was larger in the exposure than that in the outcome. SNPs that did not pass Steiger filtering were excluded(8). |
| MR Pleiotropy Residual Sum and Outlier global test (MR-PRESSO) | Horizontal pleiotropy | This method evaluates horizontal pleiotropy and corrects for it via outlier removal. It then tests for significant distortion in the causal estimate before and after outlier removal. |
| MR-LAP | Sample overlap, Winners Curse and weak instrument bias | MR-Lap uses cross-trait LD-score regression (LDSC) to simultaneously account and correct for winner’s curse, weak instrument bias and sample overlap(9). |

| **Supplementary Table S22: Number of SNPs available for each analysis and those removed due to not fulfilling selection criteria for specific PUFA genes.** | | | | | | | |
| --- | --- | --- | --- | --- | --- | --- | --- |
| Genome-wide significant SNPs (p<5x10^-8^) | Outcome | Available SNPs in outcome | Palindromic SNPs with intermediate allele frequencies | SNPs in LD | SNPs that did not pass Steiger filtering | | Available SNPs in the analysis |
| Exposure GWAS: DHA FADS gene cluster | | | | | | | |
| 11 | CRP | 9 | 2 | 0 | 0 | | 7 |
| 11 | GlycA | 11 | 0 | 0 | 0 | | 11 |
| 11 | IL-6 | 1 | 0 | 0 | 0 | | 1 |
| Exposure GWAS: LA FADS gene cluster | | | | | | | |
| 7 | CRP | 6 | 0 | 0 | 1 | 5 | |
| 7 | GlycA | 7 | 0 | 0 | 0 | 7 | |
| 7 | IL-6 | 0 | 0 | 0 | 0 | 0 | |
| Exposure GWAS: DHA ELOVL2 gene | | | | | | | |
| 9 | CRP | 7 | 2 | 0 | 0 | 5 | |
| 9 | GlycA | 9 | 0 | 0 | 2 | 7 | |
| 9 | IL-6 | 3 | 0 | 0 | 1 | 2 | |
| Exposure GWAS: LA ELOVL2 gene | | | | | | | |
| 17 | CRP | 15 | 5 | 0 | 1 | 9 | |
| 17 | GlycA | 17 | 0 | 0 | 3 | 14 | |
| 17 | IL-6 | 1 | 0 | 0 | 0 | 1 | |
| *SNPS, Single nucleotide polymorphisms; LD, Linkage disequilibrium; GWAS, genome-wide association study; DHA, Docosahexaenoic acid; LA, Linoleic acid; IL-6, Interleukin-6; CRP, C-reactive protein; GlycA, Glycoprotein acetyls; FADs, Fatty acid desaturase; ELOVL2, Elongation of Very Long Chain Fatty acids.* | | | | | | | |

| **Supplementary Table S23: Harmonized SNPs for the analysis of the effect of SNPs from the *FADS* gene cluster on inflammatory biomarkers with palindromic SNPS and SNPs in LD removed (1)** | | | | | | | | | | | | |
| --- | --- | --- | --- | --- | --- | --- | --- | --- | --- | --- | --- | --- |
| **Exposure** | **Outcome** | **SNP** | **Effect Allele** | **Other Allele** | **Beta** | **EAF** | **SE** | **p-value** | **F statistic** | **R^2^ exposure** | **Steiger direction** | **Steiger p-value** |
| DHA | GlycA | rs118173529 | T | C | 0.03 | 0.02 | 0.01 | 0.01 | 4.88 | 4.67x10^-05^ | TRUE | 0.161748 |
| DHA | GlycA | rs146148808 | G | C | -0.04 | 0.01 | 0.03 | 0.12 | 1.74 | 1.54x10^-05^ | TRUE | 0.424083 |
| DHA | GlycA | rs150156331 | A | G | 0.11 | 0.02 | 0.01 | 1.30x10^-15^ | 60.56 | 0.000534 | TRUE | 1.72x10^-07^ |
| DHA | GlycA | rs150684478 | T | G | -0.16 | 0.01 | 0.03 | 1.20x10^-10^ | 45.46 | 0.000351 | TRUE | 0.000101 |
| DHA | GlycA | rs174528 | C | T | -0.27 | 0.38 | 0.004 | 1x10^-300^ | 3929.49 | 0.035043 | TRUE | 0 |
| DHA | GlycA | rs28720292 | A | G | -0.03 | 0.02 | 0.01 | 0.037 | 4.54 | 4.34x10^-05^ | TRUE | 0.533439 |
| DHA | GlycA | rs543711028 | C | G | -0.18 | 0.01 | 0.02 | 1.50E^-22^ | 94.30 | 0.000838 | TRUE | 3.05x10^-11^ |
| DHA | GlycA | rs72918068 | G | A | 0.06 | 0.04 | 0.01 | 1.30x10^-09^ | 32.89 | 0.000313 | TRUE | 8.44x10^-05^ |
| DHA | GlycA | rs78151430 | T | A | 0.04 | 0.04 | 0.01 | 0.00041 | 11.58 | 9.86x10^-05^ | TRUE | 0.046416 |
| DHA | GlycA | rs78615324 | T | C | -0.07 | 0.01 | 0.02 | 5.40x10^-05^ | 14.44 | 0.000138 | TRUE | 0.008829 |
| DHA | GlycA | rs79136768 | G | A | -0.05 | 0.02 | 0.01 | 9.60x10^-05^ | 13.64 | 0.000127 | TRUE | 0.156779 |
| DHA | CRP | rs118173529 | T | C | 0.03 | 0.02 | 0.01 | 0.01 | 4.88 | 4.67x10^-05^ | TRUE | 0.048921 |
| DHA | CRP | rs150156331 | A | G | 0.11 | 0.02 | 0.01 | 1.30x10^-15^ | 60.56 | 0.000534 | TRUE | 2.52x10^-12^ |
| DHA | CRP | rs174528 | C | T | -0.27 | 0.38 | 0.004 | 1x10^-300^ | 3929.49 | 0.035043 | TRUE | 0 |
| DHA | CRP | rs28720292 | A | G | -0.03 | 0.02 | 0.01 | 0.037 | 4.54 | 4.34x10^-05^ | TRUE | 0.388448 |
| DHA | CRP | rs72918068 | G | A | 0.06 | 0.04 | 0.01 | 1.30x10^-09^ | 32.89 | 0.000313 | TRUE | 3.05x10^-06^ |
| DHA | CRP | rs78615324 | T | C | -0.07 | 0.01 | 0.02 | 5.40x10^-05^ | 14.44 | 0.000138 | TRUE | 0.000887 |
| DHA | CRP | rs79136768 | G | A | -0.05 | 0.02 | 0.01 | 9.60x10^-05^ | 13.64 | 0.000127 | TRUE | 0.028902 |
| DHA | IL-6 | rs174528 | C | T | -0.27 | 0.38 | 0.004 | 1x10^-300^ | 3928.49 | 0.035 | TRUE | 1.20x10^-276^ |
| LA | GlycA | rs12791961 | A | C | 0.05 | 0.99 | 0.02 | 0.0099 | 6.76 | 6.14x10^-05^ | TRUE | 0.27 |
| LA | GlycA | rs141144647 | A | G | 0.06 | 0.01 | 0.02 | 0.0011 | 10.81 | 9.36x10^-05^ | TRUE | 0.05 |
| LA | GlycA | rs148623923 | A | G | -0.04 | 0.01 | 0.02 | 0.029 | 5.30 | 4.18x10^-05^ | TRUE | 0.31 |
| LA | GlycA | rs150684478 | T | G | 0.08 | 0.01 | 0.03 | 0.0038 | 10.08 | 7.53x10^-05^ | TRUE | 0.14 |
| LA | GlycA | rs174564 | G | A | 0.08 | 0.35 | 0.004 | 5.10x10^-88^ | 371.34 | 3.42x10^-03^ | TRUE | 5.09x10^-43^ |
| LA | GlycA | rs198751 | A | G | -0.02 | 0.12 | 0.01 | 0.00048 | 12.88 | 1.18x10^-04^ | TRUE | 0.05 |
| LA | GlycA | rs72918068 | G | A | -0.005 | 0.04 | 0.01 | 0.66 | 0.24 | 2.15x10^-06^ | TRUE | 0.97 |
| *Supplementary Table S23: Harmonized SNPs for the analysis of the effect of SNPs from the FADS gene cluster on inflammatory biomarkers with palindromic SNPS and SNPs in LD removed cont. (2)* | | | | | | | | | | | | |
| LA | CRP | rs12791961 | A | C | 0.05 | 0.99 | 0.02 | 0.01 | 6.76 | 6.14x10^-05^ | TRUE | 0.02 |
| LA | CRP | rs141144647 | A | G | 0.06 | 0.01 | 0.02 | 0.001 | 10.81 | 9.36x10^-05^ | TRUE | 0.04 |
| LA | CRP | rs148623923 | A | G | -0.04 | 0.01 | 0.02 | 0.03 | 5.30 | 4.18x10^-05^ | TRUE | 0.08 |
| LA | CRP | rs174564 | G | A | 0.08 | 0.35 | 0.004 | 5.10x10^-88^ | 371.34 | 0.003 | TRUE | 7.19x10^-56^ |
| LA | CRP | rs198751 | A | G | -0.02 | 0.12 | 0.01 | 0.0005 | 12.88 | 0.0001 | TRUE | 0.003 |
| LA | CRP | rs72918068 | G | A | -0.005 | 0.04 | 0.01 | 0.66 | 0.23 | 2.15x10^-06^ | FALSE | 0.72 |
| *SNPS, Single nucleotide polymorphisms; LD, Linkage disequilibrium; GWAS, genome-wide association study; DHA, Docosahexaenoic acid; LA, Linoleic acid; Total n-3, Total omega-3; Total n-6, Total omega-6; IL-6, Interleukin-6; CRP, C-reactive protein; GlycA, Glycoprotein acetyls; FADs, Fatty acid desaturase.* | | | | | | | | | | | | |

| **Supplementary Table S24: Harmonized SNPs for the analysis of the effect of SNPs from the *ELOVL2* gene cluster on inflammatory biomarkers with palindromic SNPS and SNPs in LD removed (1)** | | | | | | | | | | | | |
| --- | --- | --- | --- | --- | --- | --- | --- | --- | --- | --- | --- | --- |
| **Exposure** | **Outcome** | **SNP** | **Effect Allele** | **Other Allele** | **Beta** | **EAF** | **SE** | **p-value** | **F statistic** | **R^2^ exposure** | **Steiger direction** | **Steiger p-value** |
| DHA | GlycA | rs1226086 | A | G | 0.01 | 0.75 | 0.005 | 0.04 | 3.79 | 3.52x10^-05^ | TRUE | 0.21 |
| DHA | GlycA | rs145062461 | C | G | -0.003 | 0.02 | 0.02 | 0.85 | 0.03 | 2.43x10^-07^ | FALSE | 1.00 |
| DHA | GlycA | rs187149103 | A | G | -0.02 | 0.01 | 0.02 | 0.22 | 1.24 | 9.89x10^-06^ | FALSE | 0.77 |
| DHA | GlycA | rs533715981 | A | C | -0.01 | 0.03 | 0.01 | 0.29 | 1.03 | 8.76x10^-06^ | TRUE | 0.80 |
| DHA | GlycA | rs6935281 | T | C | 0.01 | 0.52 | 0.004 | 0.01 | 5.94 | 5.61x10^-05^ | TRUE | 0.13 |
| DHA | GlycA | rs7765241 | C | G | 0.01 | 0.39 | 0.004 | 0.04 | 3.36 | 3.19x10^-05^ | TRUE | 0.49 |
| DHA | GlycA | rs79666600 | T | C | -0.04 | 0.03 | 0.01 | 0.0004 | 12.37 | 1.13x10^-04^ | TRUE | 0.03 |
| DHA | GlycA | rs9380082 | T | C | -0.02 | 0.22 | 0.005 | 0.0001 | 13.52 | 1.24x10^-04^ | TRUE | 0.03 |
| DHA | GlycA | rs9466618 | C | T | -0.04 | 0.02 | 0.01 | 0.01 | 6.75 | 6.31x10^-05^ | TRUE | 0.20 |
| DHA | CRP | rs1226086 | A | G | 0.01 | 0.75 | 0.005 | 0.04 | 3.79 | 3.52x10^-05^ | TRUE | 0.12 |
| DHA | CRP | rs6935281 | T | C | 0.01 | 0.52 | 0.004 | 0.01 | 5.94 | 5.61x10^-05^ | TRUE | 0.19 |
| DHA | CRP | rs79666600 | T | C | -0.04 | 0.03 | 0.01 | 0.0004 | 12.37 | 1.13x10^-04^ | TRUE | 0.001 |
| DHA | CRP | rs9380082 | T | C | -0.02 | 0.22 | 0.005 | 0.0001 | 13.52 | 1.24x10^-04^ | TRUE | 0.001 |
| DHA | CRP | rs9466618 | C | T | -0.04 | 0.02 | 0.01 | 0.01 | 6.75 | 6.31x10^-05^ | TRUE | 0.05 |
| DHA | IL-6 | rs6935281 | T | C | 0.01 | 0.52 | 0.004 | 0.01 | 5.94 | 5.61x10^-05^ | TRUE | 0.34 |
| DHA | IL-6 | rs7765241 | C | G | 0.01 | 0.39 | 0.004 | 0.04 | 3.36 | 3.19x10^-05^ | FALSE | 0.12 |
| DHA | IL-6 | rs9466618 | C | T | -0.04 | 0.02 | 0.01 | 0.01 | 6.75 | 6.31x10^-05^ | TRUE | 0.90 |
| LA | GlycA | rs115705479 | T | C | 0.07 | 0.01 | 0.02 | 0.002 | 11.21 | 9.47x10^-05^ | TRUE | 0.22 |
| LA | GlycA | rs13202931 | C | G | -0.02 | 0.02 | 0.02 | 0.13 | 1.98 | 1.68x10^-05^ | TRUE | 0.33 |
| LA | GlycA | rs138237564 | A | T | 0.01 | 0.005 | 0.03 | 0.90 | 0.12 | 1.02x10^-06^ | TRUE | 0.84 |
| LA | GlycA | rs141011534 | A | G | -0.01 | 0.02 | 0.02 | 0.53 | 0.47 | 4.05x10^-06^ | FALSE | 0.94 |
| LA | GlycA | rs148830072 | T | C | -0.04 | 0.01 | 0.02 | 0.04 | 4.65 | 3.85x10^-05^ | TRUE | 0.72 |
| LA | GlycA | rs190357339 | C | T | 0.003 | 0.01 | 0.02 | 0.73 | 0.02 | 1.33x10^-07^ | FALSE | 0.43 |
| LA | GlycA | rs35126712 | G | A | 0.01 | 0.05 | 0.01 | 0.18 | 1.49 | 1.37x10^-05^ | TRUE | 0.39 |
| LA | GlycA | rs546469416 | G | A | -0.06 | 0.01 | 0.02 | 0.02 | 5.87 | 5.21x10^-05^ | TRUE | 0.37 |
| *Supplementary Table S24: Harmonized SNPs for the analysis of the effect of SNPs from the ELOVL2 gene cluster on inflammatory biomarkers with palindromic SNPS and SNPs in LD removed cont. (2)* | | | | | | | | | | | | |
| LA | GlycA | rs55714642 | C | T | -0.02 | 0.02 | 0.01 | 0.12 | 1.90 | 1.76x10^-05^ | TRUE | 0.50 |
| LA | GlycA | rs72827179 | G | A | -0.02 | 0.02 | 0.02 | 0.37 | 1.05 | 9.67x10^-06^ | TRUE | 0.71 |
| LA | GlycA | rs72832919 | C | T | 0.02 | 0.03 | 0.01 | 0.04 | 3.72 | 3.45x10^-05^ | TRUE | 0.24 |
| LA | GlycA | rs73362883 | G | A | -0.04 | 0.02 | 0.01 | 0.01 | 6.80 | 5.75x10^-05^ | TRUE | 0.20 |
| LA | GlycA | rs75321218 | T | C | 0.03 | 0.03 | 0.01 | 0.10 | 4.32 | 3.91x10^-05^ | TRUE | 0.18 |
| LA | GlycA | rs79795291 | C | G | 0.01 | 0.002 | 0.04 | 0.93 | 0.05 | 4.57x10^-07^ | FALSE | 0.66 |
| LA | GlycA | rs79945859 | A | C | -0.02 | 0.09 | 0.01 | 0.01 | 8.29 | 7.53x10^-05^ | TRUE | 0.23 |
| LA | GlycA | rs80056894 | C | G | 0.03 | 0.03 | 0.01 | 0.02 | 4.74 | 4.16x10^-05^ | TRUE | 0.34 |
| LA | GlycA | rs9467242 | G | C | 0.02 | 0.02 | 0.02 | 0.25 | 0.99 | 9.08x10^-06^ | TRUE | 0.89 |
| LA | CRP | rs115705479 | C | T | 0.07 | 0.01 | 0.02 | 0.002 | 11.21 | 9.47x10^-05^ | TRUE | 0.01 |
| LA | CRP | rs141011534 | G | A | -0.01 | 0.02 | 0.02 | 0.53 | 0.47 | 4.05x10^-06^ | TRUE | 0.65 |
| LA | CRP | rs190357339 | T | C | 0.003 | 0.01 | 0.02 | 0.73 | 0.02 | 1.33x10^-07^ | FALSE | 1.00 |
| LA | CRP | rs35126712 | A | G | 0.01 | 0.05 | 0.01 | 0.18 | 1.49 | 1.37x10^-05^ | TRUE | 0.28 |
| LA | CRP | rs55714642 | T | C | -0.02 | 0.02 | 0.01 | 0.12 | 1.90 | 1.76x10^-05^ | TRUE | 0.38 |
| LA | CRP | rs72827179 | A | G | -0.02 | 0.02 | 0.02 | 0.37 | 1.05 | 9.67x10^-06^ | TRUE | 0.60 |
| LA | CRP | rs72832919 | T | C | 0.02 | 0.03 | 0.01 | 0.04 | 3.72 | 3.45x10^-05^ | TRUE | 0.07 |
| LA | CRP | rs73362883 | A | G | -0.04 | 0.02 | 0.01 | 0.01 | 6.80 | 5.75x10^-05^ | TRUE | 0.04 |
| LA | CRP | rs75321218 | C | T | 0.03 | 0.03 | 0.01 | 0.10 | 4.32 | 3.91x10^-05^ | TRUE | 0.06 |
| LA | CRP | rs79945859 | C | A | -0.02 | 0.09 | 0.01 | 0.01 | 8.29 | 7.53x10^-05^ | TRUE | 0.02 |
| LA | IL-6 | rs9467242 | C | G | 0.02 | 0.02 | 0.02 | 0.25 | 0.99 | 9.08x10^-06^ | TRUE | 0.95 |
| *SNPS, Single nucleotide polymorphisms; LD, Linkage disequilibrium; GWAS, Genome-wide association study; DHA, Docosahexaenoic acid; LA, Linoleic acid; Total n-3, Total omega-3; Total n-6, Total omega-6; IL-6, Interleukin-6; CRP, C-reactive protein; GlycA, Glycoprotein acetyls; ELOVL2, Elongation of Very Long Chain Fatty acids.* | | | | | | | | | | | | |

| **Supplementary Table S25: Associations of polyunsaturated fatty acids and inflammatory biomarkers in the ALSPAC complete case analysis (N = 1755)** | | | | | | | | |
| --- | --- | --- | --- | --- | --- | --- | --- | --- |
|  | **Model 1 (Unadjusted)** | | **Model 2*** | | **Model 3**** | | **Model 4***** | |
|  | **Mean difference per SD increase in exposure (se)** | **95% CI** | **Mean difference per SD increase in exposure (se)** | **95% CI** | **Mean difference per SD increase in exposure (se)** | **95% CI** | **Mean difference per SD increase in exposure (se)** | **95% CI** |
| **Outcome** |  |  |  |  |  |  |  |  |
| **Exposure: DHA at age 24y** | | | | | | | | |
| IL-6 at 24y | -0.13 (0.02) | -0.18, -0.08 | -0.13 (0.03) | -0.18, -0.09 | -0.20 (0.03) | -0.25, -0.15 | -0.16 (0.03) | -0.23, -0.09 |
| CRP at 24y | 0.11 (0.02) | 0.07, 0.16 | 0.08 (0.02) | 0.03, 0.13 | -0.0005 (0.03) | -0.05, 0.05 | -0.03 (0.03) | -0.10, 0.03 |
| GlycA at 24y | 0.28 (0.02) | 0.24, 0.33 | 0.32 (0.02) | 0.27, 0.36 | 0.10 (0.02) | 0.06, 0.13 | -0.08 (0.02) | -0.12, -0.03 |
| **Exposure: LA at age 24y** | | | | | | | | |
| IL-6 at 24y | -0.10 (0.02) | -0.15, -0.06 | -0.10 (0.02) | -0.15, -0.05 | -0.30 (0.03) | -0.37, -0.24 | -0.37 (0.05) | -0.46, -0.27 |
| CRP at 24y | 0.06 (0.02) | 0.02, 0.11 | 0.05 (0.02) | 0.01, 0.09 | -0.16 (0.03) | -0.22, -0.10 | -0.46 (0.05) | -0.56, -0.37 |
| GlycA at 24y | 0.47 (0.02) | 0.43, 0.51 | 0.48 (0.02) | 0.44, 0.52 | 0.20 (0.02) | 0.16, 0.24 | -0.01 (0.03) | -0.08, 0.05 |
| **Exposure: Total n-3 at age 24y** | | | | | | | | |
| IL-6 at 24y | -0.16 (0.02) | -0.20, -0.11 | -0.15 (0.02) | -0.19, -0.10 | -0.28 (0.03) | -0.33, -0.22 | -0.30 (0.04) | -0.38, -0.23 |
| CRP at 24y | -0.003 (0.02) | -0.05, 0.04 | -0.01 (0.02) | -0.05, 0.04 | -0.16 (0.03) | -0.21, -0.11 | -0.33 (0.04) | -0.40, -0.26 |
| GlycA at 24y | 0.32 (0.02) | 0.28, 0.36 | 0.33 (0.02) | 0.29, 0.38 | 0.04 (0.02) | 0.01, 0.08 | -0.19 (0.03) | -0.24, -0.14 |
| **Exposure: Total n-6 at age 24y** | | | | | | | | |
| IL-6 at 24y | -0.08 (0.02) | -0.13, -0.04 | -0.08 (0.02) | -0.13, -0.04 | -0.31 (0.03) | -0.37, -0.24 | -0.54 (0.07) | -0.68, -0.40 |
| CRP at 24y | 0.12 (0.02) | 0.07, 0.16 | 0.09 (0.02) | 0.05, 0.14 | -0.09 (0.03) | -0.16, -0.03 | -0.61 (0.07) | -0.74, -0.47 |
| GlycA at 24y | 0.49 (0.02) | 0.45, 0.53 | 0.51 (0.02) | 0.47, 0.55 | 0.24 (0.02) | 0.20, 0.29 | -0.05 (0.05) | -0.14, 0.05 |
| **Exposure: Total n-6:total n-3 ratio at age 24y** | | | | | | | | |
| IL-6 at 24y | 0.18 (0.02) | 0.14, 0.23 | 0.17 (0.03) | 0.13, 0.22 | 0.17 (0.02) | 0.13, 0.22 | 0.16 (0.03) | 0.11, 0.21 |
| CRP at 24y | 0.19 (0.02) | 0.15, 0.24 | 0.16 (0.02) | 0.12, 0.21 | 0.16 (0.02) | 0.11, 0.20 | 0.15 (0.02) | 0.11, 0.20 |
| GlycA at 24y | 0.16 (0.02) | 0.11, 0.20 | 0.15 (0.02) | 0.10, 0.19 | 0.12 (0.02) | 0.09, 0.16 | 0.14 (0.02) | 0.11, 0.17 |
| *ALSPAC, Avon longitudinal study of parents and children; SD, Standard deviation; se, Standard error; 95% CI, 95% confidence interval; DHA, Docosahexaenoic acid; LA, Linoleic acid; Total n-3, Total omega-3; Total n-6, Total omega-6; IL-6, Interleukin-6; CRP, C-reactive protein; GlycA, Glycoprotein acetyls.*  ** Model 2: Estimates adjusted for household social class at birth, maternal highest education qualification at birth, maternal and paternal smoking status during pregnancy, offspring sex at birth, type of drinker at age 24 years, type of smoker at 24 years, and at age in months at 24 year clinic*  *** Model 3: As Model 2 plus additional adjustment for low-density lipoprotein-cholesterol (mmol/l) and triglycerides (mmol/l) at the 24 year clinic.*  **** Model 4: As Model 3 plus additional adjustment for saturated fatty acids (mmol/l) and monounsaturated fatty acids (mmol/l) at the 24 year clinic.* | | | | | | | | |

| **Supplementary Table S26: Distribution of characteristics in the observed and imputed ALSPAC dataset** | | | | | |
| --- | --- | --- | --- | --- | --- |
|  |  | **Observed** | |  | **Imputed (N=2,802)** |
| **Continuous variables** |  | **N** | **Mean (SD)** | **% data imputed** | **Mean (SD)*** |
| BMI at 24y clinic (kg/m^2^) |  | 2772 | 24.95 (4.98) | 1.07 | 24.96 (4.99) |
| Age at 24y clinic (months) |  | 2802 | 294.18 (9.85) | 0.00 | - |
| Average GlycA at age 24y (mmol/L) |  | 2802 | 1.24 (0.17) | 0.00 | - |
| Average CRP at age 24y (mg/L) |  | 2802 | 2.26 (5.24) | 0.00 | - |
| Average IL-6 at age 24y (NpX) |  | 2802 | 3.48 (0.77) | 0.00 | - |
| Average DHA at age 24y (mmol/L) |  | 2802 | 0.11 (0.03) | 0.00 | - |
| Average LA at age 24y (mmol/L) |  | 2802 | 2.36 (0.55) | 0.00 | - |
| Average Total n-3 at age 24y (mmol/L) |  | 2802 | 0.30 (0.08) | 0.00 | - |
| Average Total n-6 at age 24y (mmol/L) |  | 2802 | 2.91 (0.62) | 0.00 | - |
| Average LDL at age 24y (mmol/L) |  | 2801 | 2.47 (0.77) | 0.04 | 2.47 (0.77) |
| Average Triglycerides at age 24y (mmol/L) |  | 2802 | 0.99 (0.57) | 0.00 | - |
| Average MUFA at age 24y (mmol/L) |  | 2802 | 2.18 (0.66) | 0.00 | - |
| Average SFA at age 24y (mmol/L) |  | 2802 | 3.25 (0.72) | 0.00 | - |
| **Categorical variables** | **Categories** | **N** | **%** | **% data imputed** | **N (%)*** |
| Sex at birth | Female | 1791 | 63.92 | 0.00 | - |
|  | Male | 1011 | 36.08 |  | - |
| Maternal smoking during pregnancy | No | 2084 | 82.18 | 9.49 | 2301 (82.12) |
|  | Yes | 452 | 17.82 |  | 501 (17.88) |
| Paternal smoking during pregnancy | No | 1211 | 60.10 | 28.09 | 1686 (60.17) |
|  | Yes | 804 | 39.90 |  | 1116 (39.83) |
| Maternal self-reported highest education qualification | Less than degree | 1913 | 78.59 | 13.13 | 2211 (78.91) |
|  | Degree or above | 521 | 21.41 |  | 591 (21.09) |
| Smoking Status at 24y | Non-smoker | 1968 | 71.30 | 1.50 | 1994 (71.16) |
|  | Current smoker | 792 | 28.70 |  | 808 (28.84) |
| Drinking Status at 24y | Non/infrequent drinker | 1581 | 57.76 | 2.32 | 1601 (57.14) |
|  | Frequent/heavy drinker | 1156 | 42.24 |  | 1201 (42.86) |
| Household social economic position | Non-manual | 2143 | 88.70 | 13.78 | 2475 (88.33) |
|  | Manual | 273 | 11.30 |  | 327 (11.67) |
| *ALSPAC, Avon longitudinal study of parents and children; SD, standard deviation; BMI, body mass index; GlycA, Glycoprotein acetyls; CRP, C-reactive protein; IL-6, Interleukin-6; DHA, Docosahexaenoic acid; LA, Linoleic acid; Total n-3, Total omega-3; Total n-6, Total omega-6; LDL, Low-density lipoprotein; MUFA, Monounsaturated fatty acids; SFA, Saturated fatty acids*  ** Based on imputed dataset 32 (chosen at random)* | | | | | |

| **Supplementary Table S27: Distribution of characteristics in the observed UK Biobank dataset** | | | |
| --- | --- | --- | --- |
| **Continuous Variables** |  | **N** | **Mean (SD)** |
| BMI at baseline (kg/m^2^) |  | 12401 | 27.46 |
| Age at baseline clinic (years) |  | 12451 | 56.93 |
| Average GlycA at baseline (mmol/L) |  | 12451 | 0.01 |
| Average CRP at baseline (mg/L) |  | 12451 | 0.06 |
| Average DHA at baseline (mmol/L) |  | 12451 | -0.002 |
| Average LA at baseline (mmol/L) |  | 12451 | 0.001 |
| Average total n-3 at baseline (mmol/L) |  | 12451 | 0.003 |
| Average total n-6 at baseline (mmol/L) |  | 12451 | 0.004 |
| Average LDL at baseline (mmol/L) |  | 12451 | 0.003 |
| Average Triglycerides at baseline (mmol/L) |  | 12451 | 0.01 |
| Average MUFA at baseline (mmol/L) |  | 12451 | 0.01 |
| Average SFA at baseline (mmol/L) |  | 12451 | 0.01 |
| **Categorical variables** | **Categories** | **N** | **%** |
| Sex at birth | Females | 6788 | 54.52 |
|  | Males | 5663 | 45.48 |
| Maternal smoking during pregnancy | No | 7458 | 69.47 |
|  | Yes | 3278 | 30.53 |
| Self-reported highest education qualification | Less than degree | 6350 | 62.67 |
|  | Degree or above | 3751 | 37.13 |
| Smoking Status at baseline | Non-smoker | 6851 | 55.22 |
|  | Current Smoker | 5555 | 44.78 |
| Drinking Status at baseline | Non-drinker | 421 | 3.38 |
|  | Current Drinker | 12019 | 96.62 |
| *SD, standard deviation; BMI, body mass index; GlycA, Glycoprotein acetyls; CRP, C-reactive protein; DHA, Docosahexaenoic acid; LA, Linoleic acid ; Total n-3, Total omega-3; Total n-6, Total omega-6; LDL, Low-density lipoprotein; MUFA, monounsaturated fatty acids; SFA, saturated fatty acids* | | | |

| **Supplementary Table S28: Associations of PUFA and inflammatory biomarkers using UK Biobank data (N = 12451)** | | | | | | | | |
| --- | --- | --- | --- | --- | --- | --- | --- | --- |
|  | **Model 1 (Unadjusted)** | | **Model 2*** | | **Model 3**** | | **Model 4***** | |
|  | **Mean difference per SD increase in exposure (SE)** | **95% CI** | **Mean difference per SD increase in exposure (SE)** | **95% CI** | **Mean difference per SD increase in exposure (SE)** | **95% CI** | **Mean difference per SD increase in exposure (SE)** | **95% CI** |
| **Outcome** |  |  |  |  |  |  |  |  |
| **Exposure: DHA at baseline** | | | | | | | | |
| CRP at baseline | -0.01 (0.01) | -0.02, 0.01 | -0.02 (0.01) | -0.04, 0.004 | -0.01 (0.01) | -0.03, 0.01 | -0.02 (0.09) | -0.37, -0.03 |
| GlycA at baseline | 0.09 (0.01) | 0.07, 0.10 | 0.09 (0.01) | 0.07, 0.11 | 0.10 (0.01) | 0.07, 0.12 | 0.15 (0.01) | 0.13, 0.18 |
| **Exposure: LA at baseline** | | | | | | | | |
| CRP at baseline | 0.002 (0.01) | -0.02, 0.02 | -0.01 (0.01) | -0.03, 0.01 | -0.01 (0.01) | -0.02, 0.01 | -0.005 (0.01) | -0.02, 0.01 |
| GlycA at baseline | 0.41 (0.01) | 0.39, 0.42 | 0.41 (0.01) | 0.39, 0.42 | 0.05 (0.01) | 0.03, 0.07 | -0.06 (0.01) | -0.07, -0.04 |
| **Exposure: Total n-3 at baseline** | | | | | | | | |
| CRP at baseline | -0.003 (0.01) | -0.02, 0.02 | -0.01 (0.01) | -0.03, 0.01 | -0.01 (0.01) | -0.03, 0.01 | -0.01 (0.01) | -0.03, 0.005 |
| GlycA at baseline | 0.31 (0.01) | 0.30, 0.33 | 0.13 (0.01) | 0.11, 0.15 | 0.13 (0.01) | 0.11, 0.15 | 0.15 (0.01) | 0.13, 0.18 |
| **Exposure: Total n-6 at baseline** | | | | | | | | |
| CRP at baseline | 0.004 (0.01) | -0.01, 0.02 | -0.01 (0.01) | -0.03, 0.01 | -0.004 (0.01) | -0.02, 0.01 | -0.004 (0.005) | -0.01, 0.005 |
| GlycA at baseline | 0.45 (0.01) | 0.44, 0.47 | 0.45 (0.01) | 0.43, 0.47 | 0.12 (0.01) | 0.11, 0.14 | 0.003 (0.01) | -0.01, 0.02 |
| **Exposure: Total-6:total n-3 at baseline** | | | | | | | | |
| CRP at baseline | 0.004 (0.01) | -0.01, 0.02 | 0.01 (0.01) | -0.001, 0.03 | 0.01 (0.01) | -0.001, 0.03 | 0.01 (0.01) | -0.001, 0.03 |
| GlycA at baseline | 0.01 (0.01) | -0.002, 0.02 | 0.02 (0.07) | 0.002, 0.03 | 0.01 (0.01) | -0.01, 0.03 | 0.002 (0.01) | -0.02, 0.02 |
| *SD, standard deviation; se, standard error; 95% CI, 95% confidence interval; DHA, Docosahexaenoic acid; LA, Linoleic acid; Total n-3, Total omega-3; Total n-6, Total omega-6; IL-6, interleukin-6; CRP, C-reactive protein; GlycA, Glycoprotein acetyls*  ** Model 2: Estimates adjusted for household social class, highest education qualification at baseline, maternal smoking status during pregnancy, sex at birth, type of drinker at baseline, type of smoker at baseline, and at age in years at baseline clinic*  *** Model 3: As Model 2 plus additional adjustment for low density lipoprotein-cholesterol (mmol/l) and triglycerides (mmol/l) at baseline.*  **** Model 4: As Model 3 plus additional adjustment for saturated fatty acids (mmol/l) and monounsaturated fatty acids (mmol/l) at baseline.* | | | | | | | | |

| **Supplementary Table S29: Associations between PUFAs and inflammatory biomarkers using UK Biobank data stratified by sex at birth** | | | | | | | | |
| --- | --- | --- | --- | --- | --- | --- | --- | --- |
|  | **Model 1 (Unadjusted)** | | | | **Model 2*** | | | |
|  | **Female (N = 6787)** | | **Male (N = 5662)** | | **Female (N = 6787)** | | **Male (N = 5662)** | |
|  | **Mean difference per SD increase in exposure (se)** | **95% CI** | **Mean difference per SD increase in exposure (se)** | **95% CI** | **Mean difference per SD increase in exposure (se)** | **95% CI** | **Mean difference per SD increase in exposure (se)** | **95% CI** |
| **Outcome** |  |  |  |  |  |  |  |  |
| **Exposure: DHA at age 24y** | | | | | | | | |
| CRP at baseline | -0.002 (0.01) | -0.03, 0.02 | -0.01 (0.01) | -0.04, 0.02 | -0.01 (0.01) | -0.04, 0.01 | -0.02 (0.02) | -0.05, 0.01 |
| GlycA at baseline | 0.09 (0.01) | 0.06, 0.11 | 0.09 (0.01) | 0.06, 0.12 | 0.09 (0.01) | 0.06, 0.12 | 0.08 (0.02) | 0.05, 0.01 |
| **Exposure: LA at age 24y** | | | | | | | | |
| CRP at baseline | -0.01 (0.01) | -0.03, 0.02 | 0.02 (0.01) | -0.01, 0.04 | -0.01 (0.01) | -0.04, 0.02 | -0.01 (0.02) | -0.04, 0.02 |
| GlycA at baseline | 0.40 (0.01) | 0.37, 0.42 | 0.42 (0.01) | 0.40, 0.44 | 0.41 (0.01) | 0.38, 0.43 | 0.41 (0.01) | 0.38, 0.43 |
| **Exposure: Total n-3 at age 24y** | | | | | | | | |
| CRP at baseline | -0.004 (0.01) | -0.03, 0.02 | -0.0001 (0.01) | -0.03, 0.03 | -0.01 (0.01) | -0.04, 0.01 | -0.01 (0.02) | -0.04, 0.02 |
| GlycA at baseline | 0.31 (0.01) | 0.29, 0.33 | 0.32 (0.01) | 0.29, 0.34 | 0.32 (0.01) | 0.29, 0.35 | 0.31 (0.02) | 0.27, 0.33 |
| **Exposure: Total n-6 at age 24y** | | | | | | | | |
| CRP at baseline | -0.01 (0.01) | -0.03, 0.02 | 0.01 (0.01) | -0.01, 0.05 | -0.01 (0.01) | -0.04, 0.02 | -0.01 (0.02) | -0.04, 0.02 |
| GlycA at baseline | 0.44 (0.01) | 0.42, 0.47 | 0.46 (0.01) | 0.55, 0.66 | 0.46 (0.01) | 0.43, 0.48 | 0.45 (0.01) | 0.42, 0.48 |
| **Exposure: Total-6:total n-3 at age 24y** | | | | | | | | |
| CRP at baseline | 0.01 (0.01) | -0.01, 0.03 | -0.003 (0.01) | -0.01, 0.01 | 0.03 (0.01) | 0.003, 0.05 | 0.004 (0.01) | -0.02, 0.01 |
| GlycA at baseline | 0.02 (0.01) | -0.001, 0.04 | 0.0001 (0.03) | -0.02, 0.02 | 0.03 (0.01) | 0.005, 0.06 | 0.001 (0.01) | -0.01, 0.01 |
| *SD, standard deviation; se, standard error; 95% CI, 95% confidence interval; DHA, Docosahexaenoic acid; LA, Linoleic acid; Total n-3, Total omega-3; Total n-6, Total omega-6; IL-6, interleukin-6; CRP, C-reactive protein; GlycA, Glycoprotein acetyls*  ** Model 2: Estimates adjusted for household social class, highest education qualification at baseline, maternal smoking status during pregnancy, sex at birth, type of drinker at baseline, type of smoker at baseline, and at age in years at baseline clinic.* | | | | | | | | |

| **Supplementary Table S30*:* Genetic instruments and sample sizes used to estimate SNP-exposure and SNP-outcome associations** | | | | | |
| --- | --- | --- | --- | --- | --- |
| **GWAS** | **Instrument** | **Outcome** | **Number of SNPs** | **Instrument F-statistics ^a^** | **Exposure**  **Sample Size** |
| Borges (2022)(10) | DHA | CRP | 38 | Min = 27.49  Median = 50.88  Max = 4547.24 | 114,999 |
| Borges (2022)(10) | DHA | GlycA | 46 | Min= 27.49  Median = 50.88  Max= 4547.24 | 114,999 |
| Borges (2022)(10) | DHA | IL-6 | 11 | Min = 28.22  Median = 63.09  Max = 120 | 114,999 |
| Borges (2022)(10) | LA | CRP | 37 | Min = 27.58  Median = 75.54  Max =611.02 | 114,999 |
| Borges (2022)(10) | LA | GlycA | 50 | Min = 29.40  Median = 68.23  Max = 611.02 | 114,999 |
| Borges (2022)(10) | LA | IL-6 | 13 | Min = 27.58  Median = 75.61  Max = 564.21 | 114,999 |
| Borges (2022)(10) | n-3 | CRP | 40 | Min =25.06  Median = 53.55  Max =6243.53 | 114,999 |
| Borges (2022)(10) | n-3 | GlycA | 49 | Min = 25.06  Median = 50.90  Max = 6243.53 | 114,999 |
| Borges (2022)(10) | n-3 | IL-6 | 17 | Min = 27.47  Median = 52.28  Max = 371.62 | 114,999 |
| Borges (2022)(10) | n-6 | CRP | 40 | Min = 28.78  Median = 70.71  Max = 676.17 | 114,999 |
| Borges (2022)(10) | n-6 | GlycA | 58 | Min = 28.78  Median = 59.18  Max = 676.17 | 114,999 |
| Borges (2022)(10) | n-6 | IL-6 | 18 | Min = 29.42  Median = 54.78  Max = 513.18 | 114,999 |
| *^a^Instrument strength F-statistics are based on the formulae* $R^{2}=2*MAF*\left( 1-MAF \right)*{beta}^{2}$*, where MAF=Minor allele frequency, and* $F= \frac{R^{2}* ( N-2 )}{1-R^{2}}$ *; as described in Shim et al.^17^ and Palmer et al.^18^ previously.*  *These results are post-Steiger filtering.*  *SNP, Single nucleotide polymorphism, GWAS, Genome-wide association study; DHA, Docosahexaenoic acid; LA, Linoleic acid; Total n-3, Total omega-3; Total n-6, Total omega-6; IL-6, Interleukin-6; CRP, C-reactive protein; GlycA, Glycoprotein acetyls.* | | | | | |

| **Supplementary Table S31: Results when investigating pleiotropy and heterogeneity in the MR analyses of the effect of PUFAs on different biomarkers of inflammation** | | | | | | |
| --- | --- | --- | --- | --- | --- | --- |
|  | Investigation into pleiotropy | | Investigation into heterogeneity using Cochrane’s Q and Rucker’s Q | | | |
| **PUFAs** | **MR-Egger Intercept** | | **Cochrane’s Q** | | **Rucker’s Q** | |
|  | Beta | p-value | Q | p-value | Q | p-value |
| **Exposure: DHA** | | | | | | |
| CRP | -0.002 | 0.613 | 1286.331 | p<0.005 | 1277.11 | p<0.005 |
| GlycA | -0.001 | 0.870 | 374.36 | p<0.005 | 374.09 | p<0.001 |
| IL-6 | 0.011 | 0.385 | 22.24 | 0.014 | 20.35 | 0.016 |
| **Exposure: LA** | |  |  |  |  |  |
| CRP | 0.005 | 0.363 | 1682.56 | p<0.005 | 1642.72 | p<0.005 |
| GlycA | 0.003 | 0.613 | 606.78 | p<0.005 | 602.96 | p<0.005 |
| IL-6 | 0.002 | 0.705 | 19.49 | 0.08 | 19.23 | 0.06 |
| **Exposure: n-3** | |  |  |  |  |  |
| CRP | 0.01 | 0.362 | 1682.56 | p<0.005 | 1642.72 | p<0.005 |
| GlycA | 0.011 | 0.025 | 812.92 | p<0.005 | 713.91 | p<0.005 |
| IL-6 | 0.001 | 0.813 | 21.41 | 0.163 | 21.33 | 0.127 |
| **Exposure: n-6** | |  |  |  |  |  |
| CRP | 0.01 | 0.079 | 1861.37 | p<0.001 | 1713.91 | p<0.001 |
| GlycA | 0.006 | 0.206 | 634.44 | p<0.005 | 613.87 | p<0.005 |
| IL-6 | 0.002 | 0.570 | 28.56 | 0.039 | 27.98 | 0.032 |
| *MR, Mendelian randomization; PUFAs, polyunsaturated fatty acids; GWAS, Genome-wide association study; DHA, Docosahexaenoic acid; LA, Linoleic acid; Total n-3, Total omega-3; Total n-6, Total omega-6; IL-6, Interleukin-6; CRP, C-reactive protein; GlycA, Glycoprotein acetyls.* | | | | | | |

| **Supplementary Table S32: Results when investigating pleiotropy using MR-Presso in the MR analyses of the effect of PUFAs on different biomarkers of inflammation** | | | | | | |
| --- | --- | --- | --- | --- | --- | --- |
|  | Number of Outliers | Global test | Distortion Test | | IVW results after outlier correction | |
|  |  |  | p-value | | IVW beta (CI) | p-value |
| **Exposure: DHA** | |  |  |  |  |  |
| CRP | 20 | p<0.001 | p<0.001 | | 0.09 (0.04, 0.14) | 0.002 |
| GlycA | 10 | p<0.001 | 0.175 | | -0.01 (-0.04, 0.03) | 0.630 |
| IL-6 | 1 | 0.015 | 0.031 | | -0.004 (-0.14, 0.13) | 0.950 |
| **Exposure: LA** | |  |  |  |  |  |
| CRP | 18 | p<0.001 | p<0.001 | | 0.02 (-0.01, 0.06) | 0.219 |
| GlycA | 14 | p<0.001 | 0.457 | | 0.223 (0.16, 0.29) | p<0.001 |
| IL-6 | 0 | 0.124 |  |  |  |  |
| **Exposure: Total n-3** | |  |  |  |  |  |
| CRP | 12 | p<0.001 | p<0.001 | | 0.12 (0.09, 0.15) | p<0.001 |
| GlycA | 13 | p<0.001 | 0.023 | | 0.141 (0.06, 0.22) | 0.002 |
| IL-6 | 0 | 0.125 |  |  |  |  |
| **Exposure: Total n-6** | |  |  |  |  |  |
| CRP | 18 | p<0.001 | 0.239 | | 0.03 (-0.01, 0.07) | 0.111 |
| GlycA | 13 | p<0.001 | 0.049 |  | 0.28 (0.22, 0.35) | P<0.001 |
| IL-6 | 1 | 0.039 | 0.156 | | 0.008 (-0.05, 0.06) | 0.775 |
| *MR, Mendelian randomization; PUFAs, polyunsaturated fatty acids; GWAS, Genome-wide association study; DHA, Docosahexaenoic acid; LA, Linoleic acid; Total n-3, Total omega-3; Total n-6, Total omega-6; IL-6, Interleukin-6; CRP, C-reactive protein; GlycA, Glycoprotein acetyls.* | | | | | | |

| **Supplementary Table S33*:* Genetic instruments and sample sizes used to estimate SNP-exposure and SNP-outcome associations in replication MR using UKB** | | | | | |
| --- | --- | --- | --- | --- | --- |
| **Exposure GWAS** | **Instrument** | **Outcome** | **Number of SNPs** | **Instrument F-statistics ^a^** | **Exposure**  **Sample Size** |
| Borges (2022)(10) | DHA | CRP | 40 | Min = 27.49  Median = 51.01  Max = 4547.24 | 114,999 |
| Borges (2022)(10) | DHA | IL-6 | 45 | Min = 27.49  Median = 47.38  Max = 4547.24 | 114,999 |
| Borges (2022)(10) | LA | CRP | 40 | Min = 27.58  Median = 75.58  Max =611.02 | 114,999 |
| Borges (2022)(10) | LA | IL-6 | 50 | Min = 27.58  Median = 68.55  Max = 611.02 | 114,999 |
| Borges (2022)(10) | n-3 | CRP | 41 | Min = 25.06  Median = 56.21  Max = 6243.53 | 114,999 |
| Borges (2022)(10) | n-3 | IL-6 | 48 | Min = 25.06  Median = 51.59  Max = 6243.53 | 114,999 |
| Borges (2022)(10) | n-6 | CRP | 8 | Min = 107.73  Median =228.06  Max = 486.78 | 114,999 |
| Borges (2022)(10) | n-6 | IL-6 | 9 | Min = 107.73  Median = 228.54  Max = 513.18 | 114,999 |
| *MR, Mendelian randomization; PUFAs, polyunsaturated fatty acids; GWAS, Genome-wide association study; DHA, Docosahexaenoic acid; LA, Linoleic acid; Total n-3, Total omega-3; Total n-6, Total omega-6; IL-6, Interleukin-6; CRP, C-reactive protein; GlycA, Glycoprotein acetyls*  *^a^Instrument strength F-statistics are based on the formulae* $R^{2}=2*MAF*\left( 1-MAF \right)*{beta}^{2}$*, where MAF=Minor allele frequency, and* $F= \frac{R^{2}* ( N-2 )}{1-R^{2}}$ *; as described in Shim et al.^17^ and Palmer et al.^18^ previously.*  *These results are post-Steiger filtering*. | | | | | |

| **Supplementary Table S34*:* Replication MR-IVW and sensitivity analyses investigating the effect of PUFAs on CRP and IL-6.** | | | | | |
| --- | --- | --- | --- | --- | --- |
| Exposure | SNP N | Method | Beta (SE) | 95% CI | p-value |
| Outcome: IL-6 | | | | | |
| DHA | 45 | Inverse variance weighted | -0.05 (0.02) | -0.09, -0.01 | 0.026 |
|  |  | MR Egger | -0.03 (0.03) | -0.090.03 | 0.323 |
|  |  | Weighted median | -0.03 (0.03) | -0.08, 0.02 | 0.228 |
|  |  | Weighted mode | -0.03 (0.03) | -0.08, 0.02 | 0.303 |
| LA | 50 | Inverse variance weighted | -0.02 (0.03) | -0.07, 0.04 | 0.581 |
|  |  | MR Egger | -0.01 (0.06) | -0.12, 0.10 | 0.842 |
|  |  | Weighted median | 0.02 (0.04) | -0.05, 0.09 | 0.539 |
|  |  | Weighted mode | 0.03 (0.04) | -0.05, 0.11 | 0.430 |
| Omega-3 | 48 | Inverse variance weighted | -0.04 (0.02) | -0.07, 0.002 | 0.063 |
|  |  | MR Egger | -0.03 (0.03) | -0.08, 0.02 | 0.301 |
|  |  | Weighted median | -0.03 (0.02) | -0.07, 0.02 | 0.237 |
|  |  | Weighted mode | -0.03 (0.02) | -0.07, 0.02 | 0.234 |
| Omega-6 | 9 | Inverse variance weighted | -0.04 (0.04) | -0.12, 0.03 | 0.273 |
|  |  | MR Egger | -0.07 (0.011) | -0.28, 0.14 | 0.524 |
|  |  | Weighted median | 0.01 (0.05) | -0.08, 0.11 | 0.765 |
|  |  | Weighted mode | 0.04 (0.07) | -0.10, 0.17 | 0.612 |
| Outcome: CRP | | | | | |
| DHA | 40 | Inverse variance weighted | 0.16 (0.12) | -0.07, 0.40 | 0.177 |
|  |  | MR Egger | 0.10 (0.17) | -0.24, 0.43 | 0.572 |
|  |  | Weighted median | 0.12 (0.04) | 0.05, 0.20 | 0.001 |
|  |  | Weighted mode | 0.14 (0.04) | 0.07, 0.21 | 0.0004 |
| LA | 40 | Inverse variance weighted | -0.08 (0.24) | -0.55, 0.38 | 0.723 |
|  |  | MR Egger | -0.12 (0.49) | -1.07, 0.83 | 0.808 |
|  |  | Weighted median | 0.15 (0.08) | -0.0001, 0.31 | 0.050 |
|  |  | Weighted mode | 0.26 (0.08) | 0.10, 0.42 | 0.002 |
| Omega-3 | 43 | Inverse variance weighted | 0.24 (0.08) | 0.08, 0.40 | 0.003 |
|  |  | MR Egger | 0.15 (0.11) | -0.07, 0.38 | 0.181 |
|  |  | Weighted median | 0.12 (0.03) | 0.06, 0.18 | 0.0003 |
|  |  | Weighted mode | 0.12 (0.03) | 0.05, 0.18 | 0.001 |
| Omega-6 | 8 | Inverse variance weighted | 0.58 (0.34) | -0.09, 1.25 | 0.088 |
|  |  | MR Egger | -0.40 (0.92) | -2.21, 1.41 | 0.682 |
|  |  | Weighted median | 0.39 (0.10) | 0.21, 0.58 | 4.01x10^-5^ |
|  |  | Weighted mode | 0.40 (0.11) | 0.18, 0.62 | 0.008 |
| *MR, Mendelian randomization; IVW, Inverse-variance weighted;PUFAs, polyunsaturated fatty acids; DHA, Docosahexaenoic acid; LA, Linoleic acid; IL-6, Interleukin-6; CRP, C-reactive protein.* | | | | | |

| **Supplementary Table S35: Results when investigating pleiotropy and heterogeneity in the MR analyses of the effect of PUFAs on different biomarkers of inflammation** | | | | | | |
| --- | --- | --- | --- | --- | --- | --- |
|  | Investigation into pleiotropy | | Investigation into heterogeneity using Cochrane’s Q and Rucker’s Q | | | |
| **PUFAs** | **MR-Egger Intercept** | | **Cochrane’s Q** | | **Rucker’s Q** | |
|  | Beta | p-value | Q | p-value | Q | p-value |
| **Exposure: DHA** | | | | | | |
| CRP | 0.007 | 0.591 | 645.54 | p<0.005 | 640.59 | p<0.005 |
| IL-6 | -0.002 | 0.456 | 49.90 | 0.250 | 49.25 | 0.237 |
| **Exposure: LA** | |  |  |  |  |  |
| CRP | 0.035 | 0.096 | 907.85 | p<0.005 | 843.31 | p<0.005 |
| IL-6 | -0.0003 | 0.929 | 75.00 | 0.01 | 74.99 | 0.008 |
| **Exposure: n-3** | |  |  |  |  |  |
| CRP | 0.01 | 0.262 | 444.29 | p<0.005 | 430.04 | p<0.005 |
| IL-6 | -0.001 | 0.696 | 61.99 | 0.070 | 61.78 | 0.060 |
| **Exposure: n-6** | |  |  |  |  |  |
| CRP | 0.08 | 0.299 | 246.91 | p<0.001 | 203.14 | p<0.001 |
| IL-6 | 0.002 | 0.773 | 8.92 | 0.349 | 8.81 | 0.267 |
| *MR, Mendelian randomization; IVW, Inverse-variance weighted;PUFAs, polyunsaturated fatty acids; DHA, Docosahexaenoic acid; LA, Linoleic acid; Total n-3, Total omega-3; Total n-6, Total omega-6; IL-6, Interleukin-6; CRP, C-reactive protein.* | | | | | | |

| **Supplementary Table S36: Results when investigating pleiotropy using MR-Presso in the MR analyses of the effect of PUFAs on different biomarkers of inflammation** | | | | | | |
| --- | --- | --- | --- | --- | --- | --- |
|  | Number of Outliers | Global test | Distortion Test | | IVW results after outlier correction | |
|  |  |  | p-value | | IVW Beta (95% CI) | p-value |
| **Exposure: DHA** | |  |  |  |  |  |
| CRP | 10 | p<0.005 | 0.315 | | 0.19 (0.03, 0.36) | 0.027 |
| IL-6 | 0 | 0.301 |  | |  |  |
| **Exposure: LA** | |  |  |  |  |  |
| CRP | 13 | p<0.001 | 0.812 | | 0.12 (-0.03, 0.27) | 0.121 |
| IL-6 | 1 | 0.018 | 0.897 |  | -0.20 (-0.07, 0.03) | 0.444 |
| **Exposure: Total n-3** | |  |  |  |  |  |
| CRP | 8 | p<0.001 | 0.038 | | 0.29 (0.18, 0.41) | p<0.001 |
| IL-6 | 0 | 0.110 |  |  |  |  |
| **Exposure: Total n-6** | |  |  |  |  |  |
| CRP | 4 | p<0.001 | p<0.001 | | 0.44 (0.32, 0.56) | 0.006 |
| IL-6 | 0 |  |  | |  |  |
| *MR, Mendelian randomization; IVW, Inverse-variance weighted; PUFAs, polyunsaturated fatty acids; 95% CI, 95% confidence intervals; DHA, Docosahexaenoic acid; LA, Linoleic acid; Total n-3, Total omega-3; Total n-6, Total omega-6; IL-6, Interleukin-6; CRP, C-reactive protein.* | | | | | | |

**Supplementary Figure S1: Schematic representation of polyunsaturated fatty acid biosynthesis in mammals adapted from Videla 2022 (1)**
n-3 PUFAs, Total omega-3 polyunsaturated fatty acids; n-6, Total omega-6 polyunsaturared fatty acids.

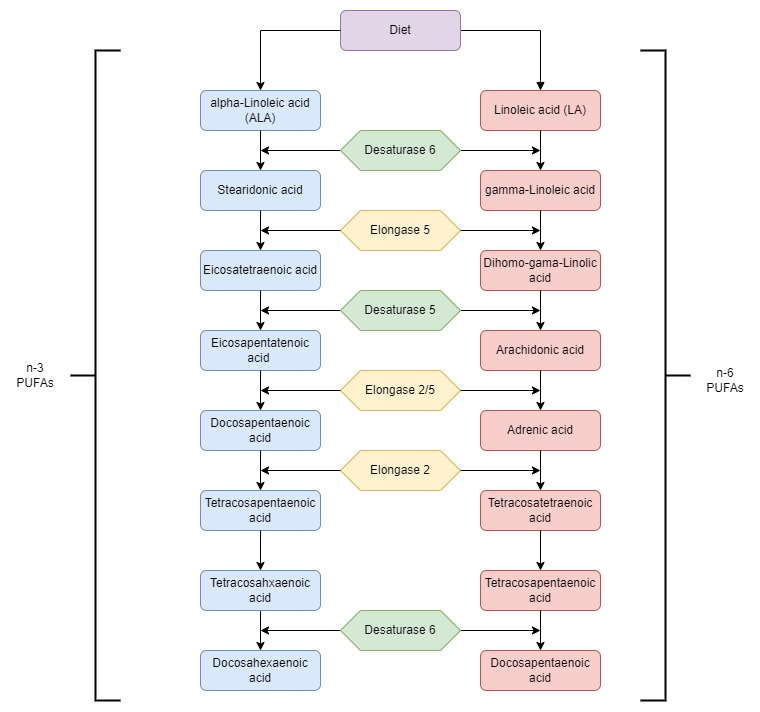

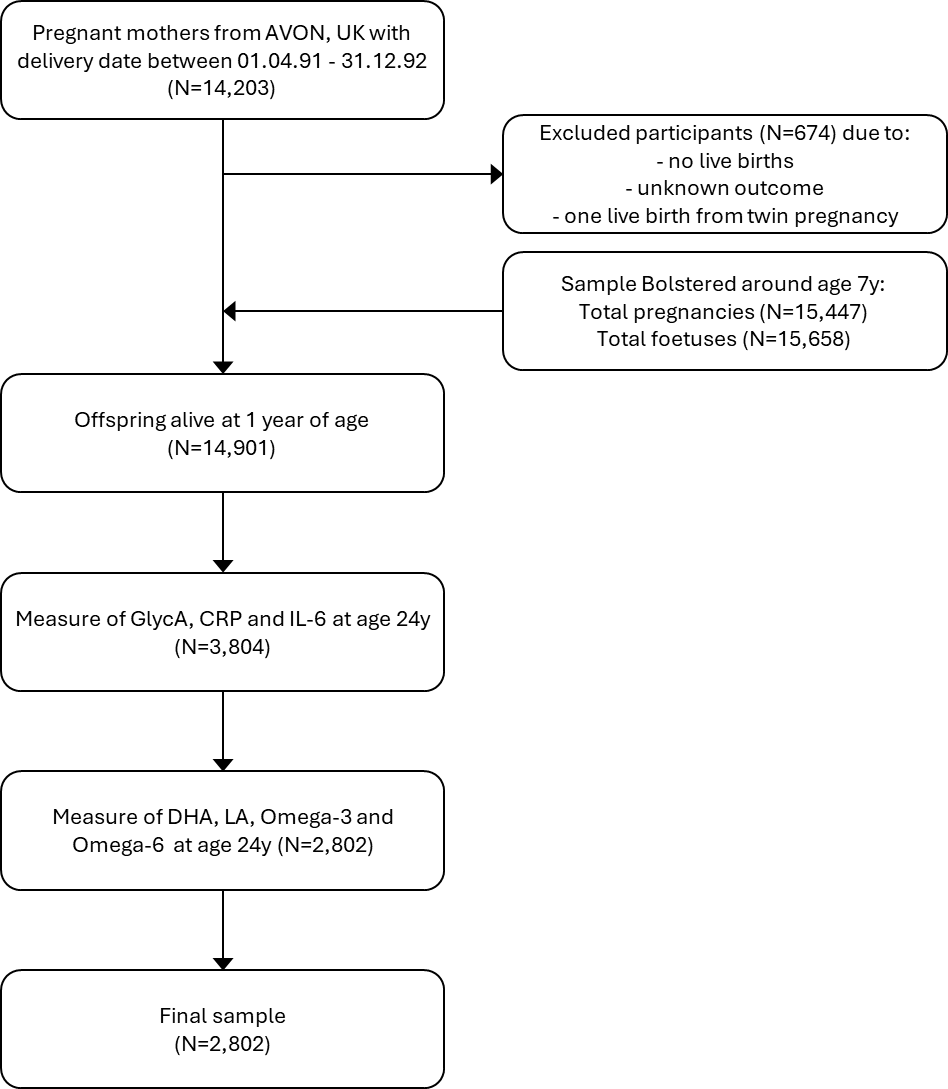

**Supplementary Figure S2: Flowchart of inclusion into the study using Avon Longitudinal Study of Parents and Children (ALSPAC) data**

*DHA, Docosahexaenoic acid; LA, Linoleic acid; Total n-3, Total omega-3; Total n-6, Total omega-6; IL-6, Interleukin-6; CRP, C-reactive protein.*

**Supplementary Figure S3: Variance inflation factors (VIFs) for each covariate included in the model 4 analysis of the association between polyunsaturated fatty acids and inflammatory biomarkers. Shades of blue and red indicate smaller and larger VIFs, respectively**

*DHA, Docosahexaenoic acid; LA, Linoleic acid.*

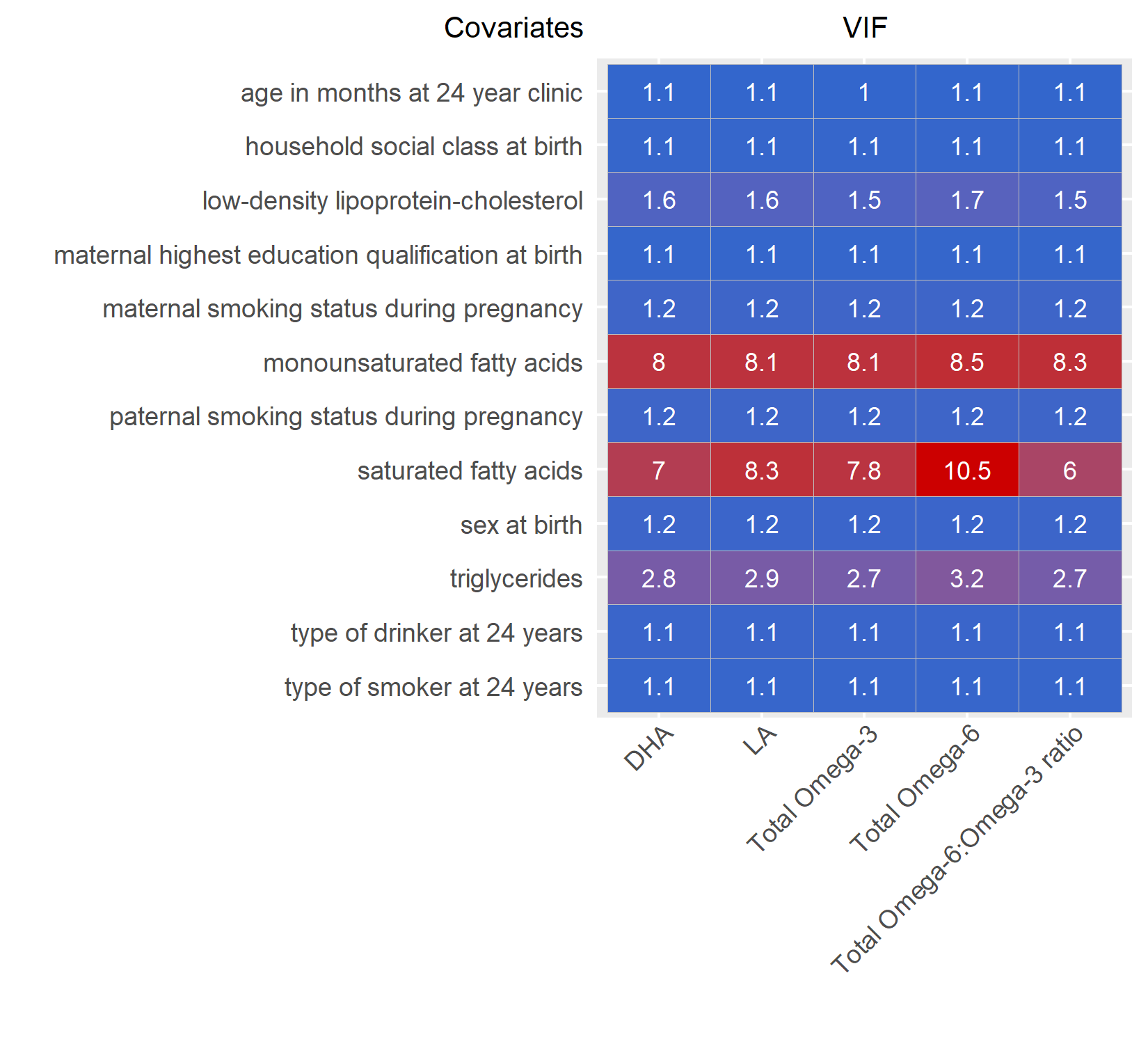

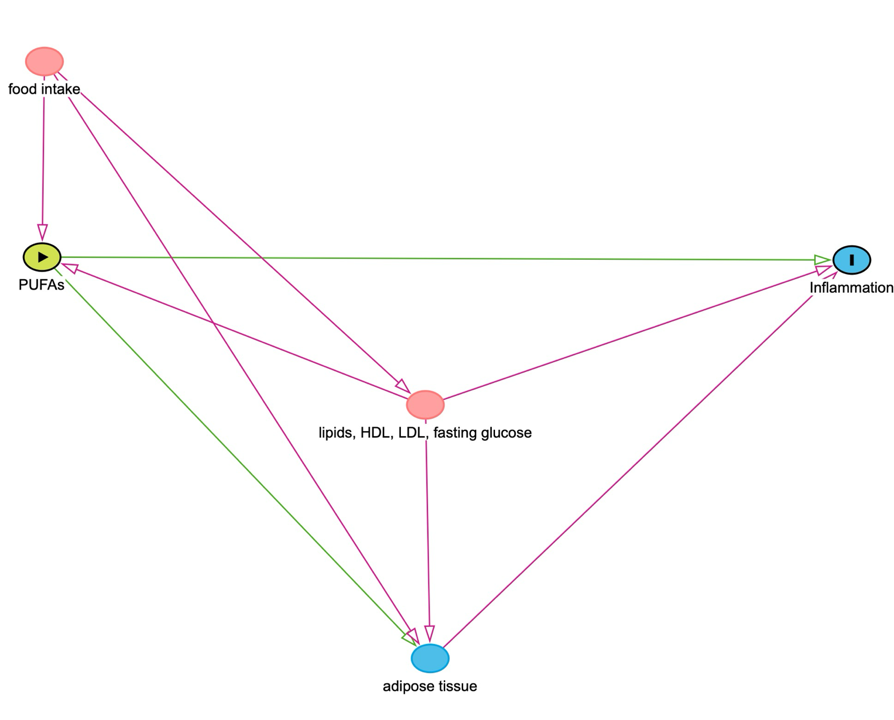

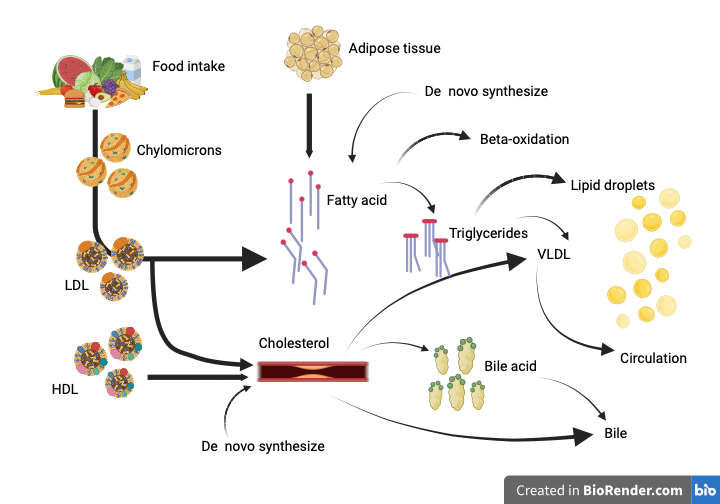

**Supplementary Figure S4: Pictorial representation of the inter-related metabolomic pathway between fatty acids, LDL-cholesterol and triglycerides and their influence on inflammation.**

*LDL, Low-density lipoprotein; HDL, High-density lipoprotein; VLDL, Very-low-density lipoprotein;PUFAs, Polyunsaturated fatty acids.*
*Nb: Pictorial representation created with BioRender.com.*

**Supplementary Figure S5: Univariable Mendelian randomization analysis investigating the causal effect of Polyunsaturated fatty acids (DHA, LA, omega-3 and omega-6) on levels of three biomarkers of inflammation (CRP, IL-6 and GlycA).**
Plot A reflects the effect of PUFAs on the biomarker CRP, Plot B reflects the effect of PUFAs on the biomarker IL-6 and Plot C reflects the effect of PUFAs on the biomarker GlycA.

*DHA, Docosahexaenoic acid; LA, Linoleic acid; Total n-3, Total omega-3; Total n-6, Total omega-6; IL-6, Interleukin-6; CRP, C-reactive protein; GlycA, Glycoprotein acetyls, IVW, Inverse-weighted variance.*

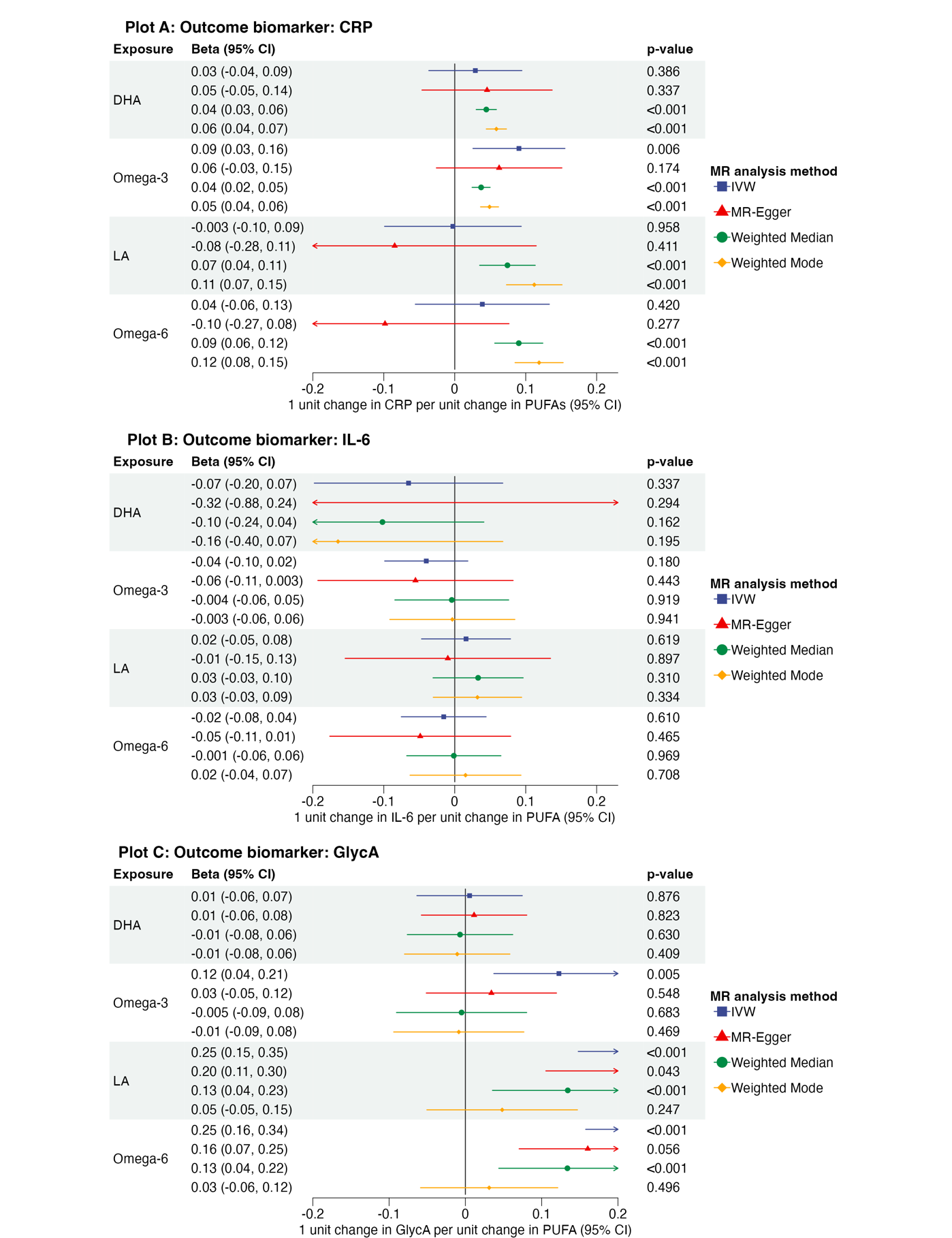

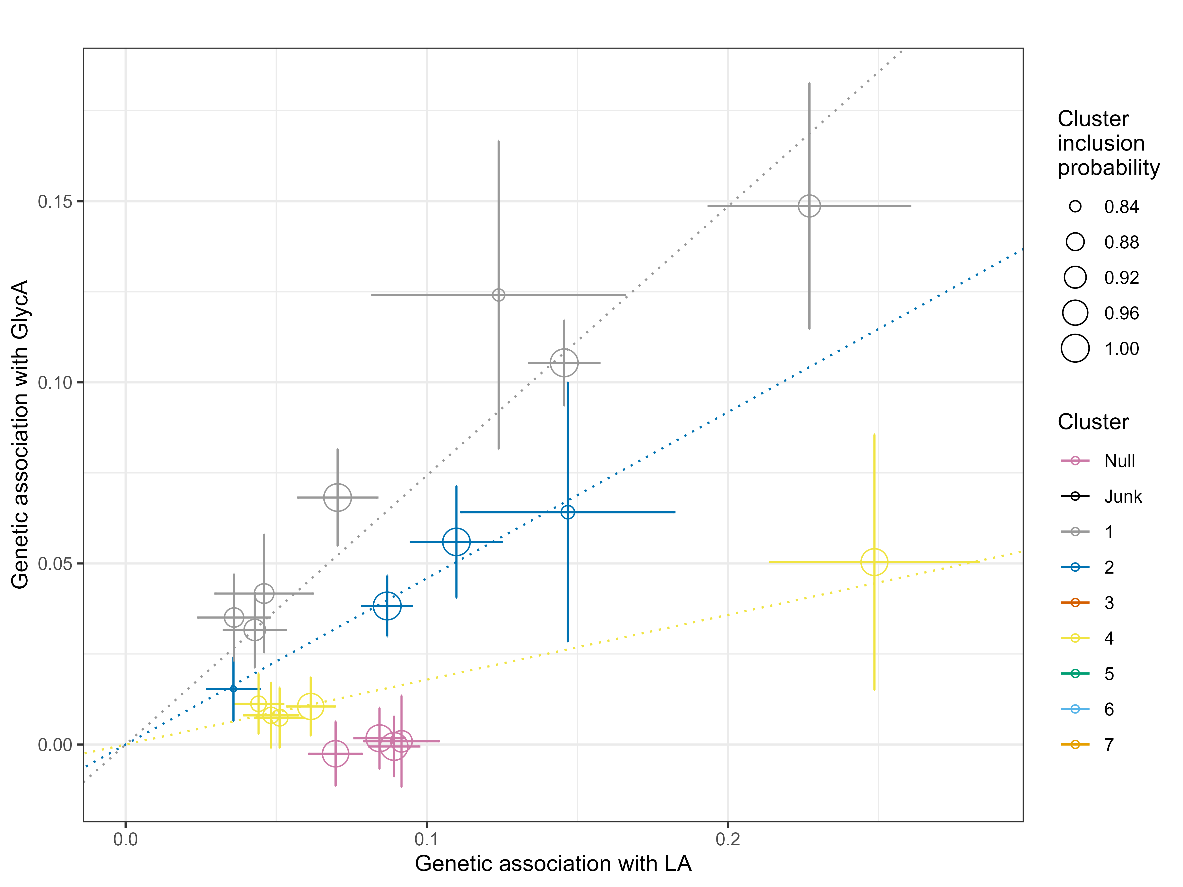

**Supplementary Figure S6: Clusters identified using MR-Clust for the effect of LA on GlycA**

*LA, Linolic Acid; GlycA, Glycoprotein Acetyl; MR, Mendelian randomization.
N.B. Key provides all clusters identified, but only those which fulfil threshold criteria are presented in the plot.*

**Supplementary Figure S7: Clusters identified using MR-Clust for the effect of total omega-3 on GlycA**

*N-3 PUFAs, Total omega-3 polyunsatruated fatty acids; GlycA, Glycoprotein Acetyl; MR, Mendelian randomization.
N.B. Key provides all clusters identified, but only those which fulfil threshold criteria are presented in the plot.*

*N.B. Key provides all clusters identified, but only those which fulfil threshold criteria are presented in the plot*

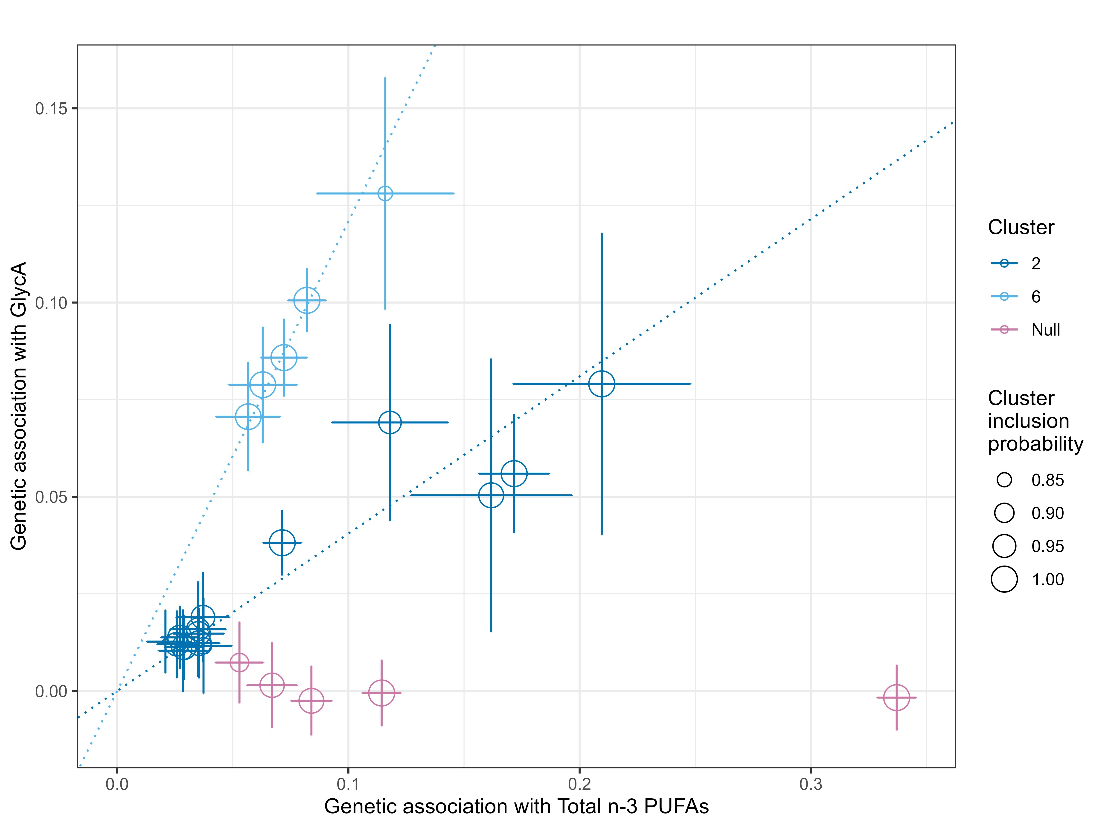

**Supplementary Figure S8: Clusters identified using MR-Clust for the effect of total omega-6 on GlycA**

*N-6 PUFAs, Total omega-6 polyunsatruated fatty acids; GlycA, Glycoprotein Acetyl; MR, Mendelian randomization.*
*N.B. Key provides all clusters identified, but only those which fulfil threshold criteria are presented in the plot.*

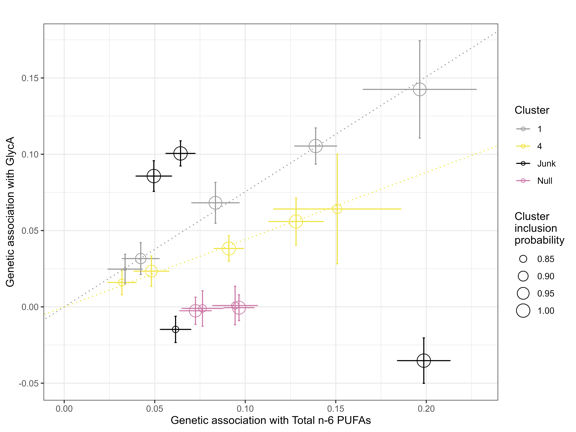

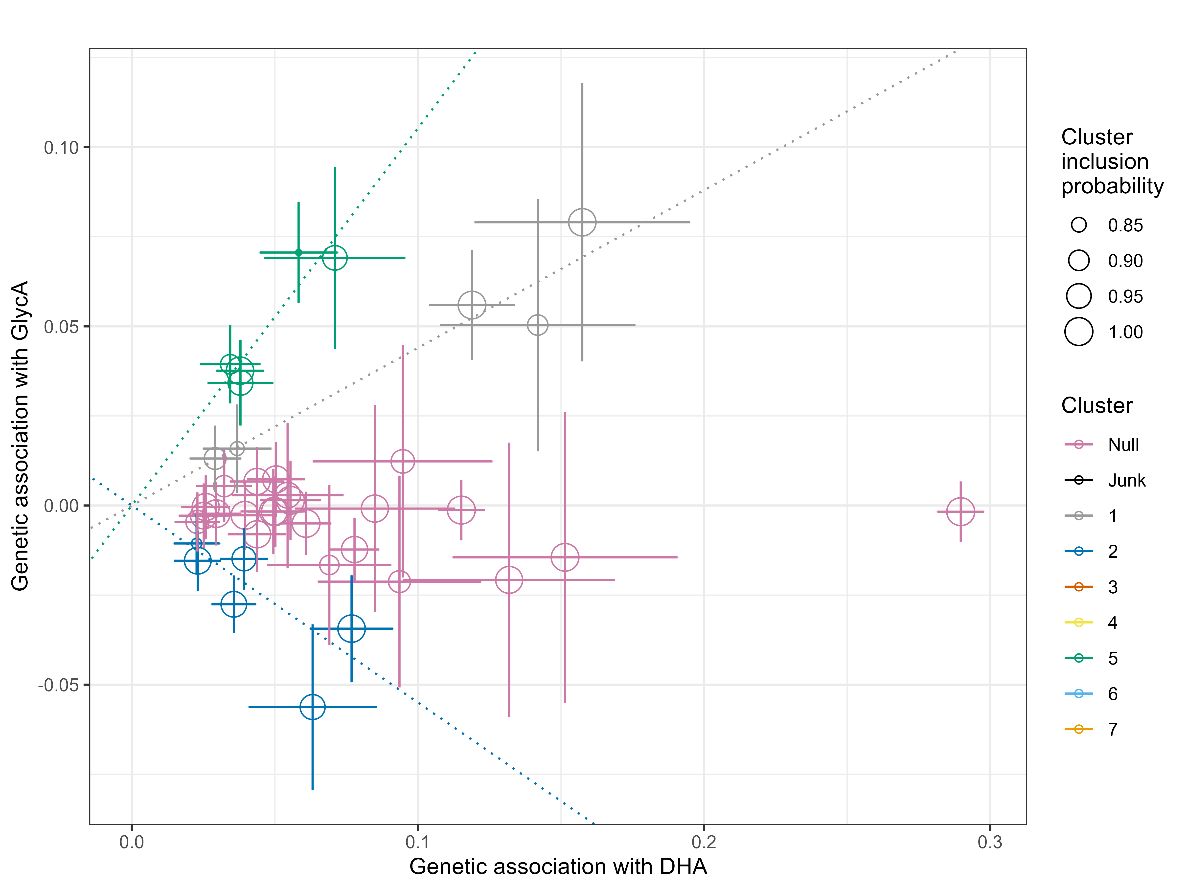

**Supplementary Figure S9: Clusters identified using MR-Clust for the effect of DHA on GlycA**

*DHA, Docosahexaenoic acid; GlycA, Glycoprotein Acetyl; MR, Mendelian randomization.*
*N.B. Key provides all clusters identified, but only those which fulfil threshold criteria are presented in the plot.*

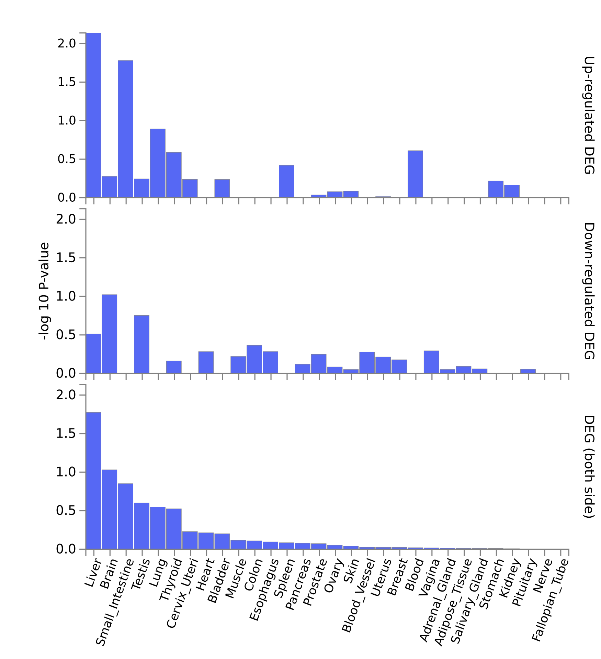

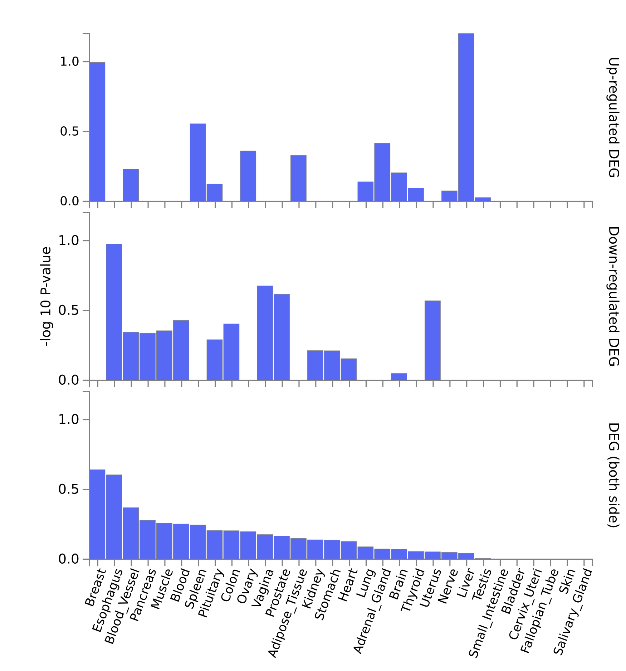

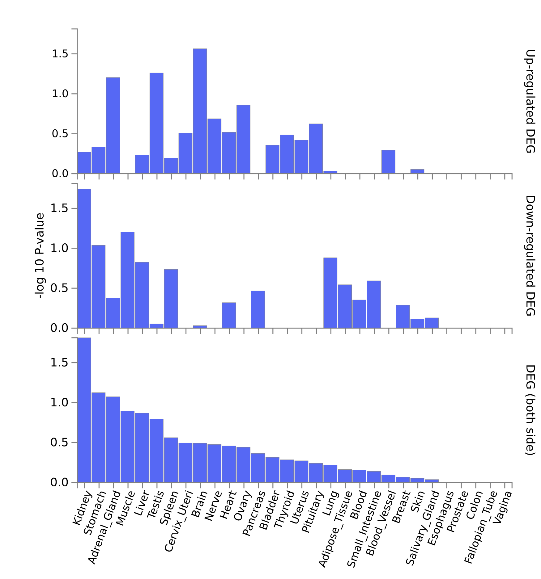

**Supplementary Figure S10: Tissue specific differentially expressed genes (DEG) results based on MR-Clust SNP cluster membership for the effect of DHA on the outcome GlycA: Cluster 1 (left), Cluster 2 (middle) and Cluster 5 (right).**

*MR, Mendelian randomization; SNP, Single nucleotide polymorphism; DHA, Docosahexaenoic acid; GlycA, Glycoprotein Acetyl.*

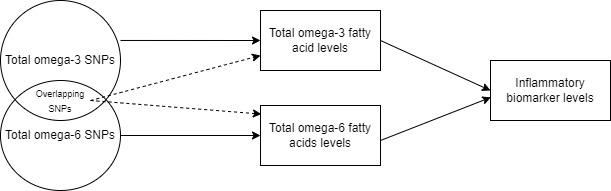

**Supplementary Figure S11: Directed Acyclic Graph demonstrating the direct (solid arrows) and indirect (dashed arrows) effects in the Multivariable Mendelian Randomisation analysis**

*SNP, Single nucleotide polymorphism.*
